# Assessing MMR Vaccination Strategies in Texas: A Scenario-Based Modeling Study of Measles Outbreaks

**DOI:** 10.1101/2025.03.25.25324613

**Authors:** Kaiming Bi, Thuy Nguyen, Boya Peng, Trudy Krause, Cecilia Ganduglia Cazaban, Janelle Rios, Cici Bauer, Catherine Troisi, Eric Boerwinkle, Aanand D. Naik

## Abstract

Measles, once common in the U.S., declined sharply after the MMR vaccine’s adoption in 1971 and was declared eliminated in the U.S. in 2000. Measles has reemerged due to waning vaccination coverage and case importation and more than 200 cases reported in the past few months in West Texas. We used a calibrated mathematical model and stochastic simulations to assess the impact of vaccination on the expected number of cases. Raising vaccination coverage by 5% could prevent 133 cases (95% CI: 97–169), while a 5% reduction may result in 190 additional cases (95% CI: 133–273) in Gaines County. These findings highlight the critical need for robust vaccination strategies to mitigate measles outbreak risks.

## Introduction

Measles was a ubiquitous childhood disease in the United States until widespread adoption of the measles-mumps-rubella (MMR) vaccine in 1971. This led to a dramatic decline in cases and declaration of measles elimination in 2000 [1]. Periodic outbreaks have since reemerged, driven by lapses in vaccination and the importation of cases from abroad [2]. On January 29, 2025, local transmission was identified among four school-aged children in Gaines County, Texas [3]. By March 11, 2025, Texas reported 223 measles cases with 156 occurring in Gaines County alone.

These events underscore the challenges in maintaining community immunity in the setting of vaccine hesitancy [4] and disruption of routine vaccination programs, including drops in MMR coverage [5]. In Texas, the Kindergarten MMR vaccination rates fell from 98.51% in 2013-14 to 94.25% in 2024-25, and in Gaines County, it declined even more, dropping from 92.55% to 81.97% over the same periods (**Figure 1.A**) [6].

**Figure 1.**
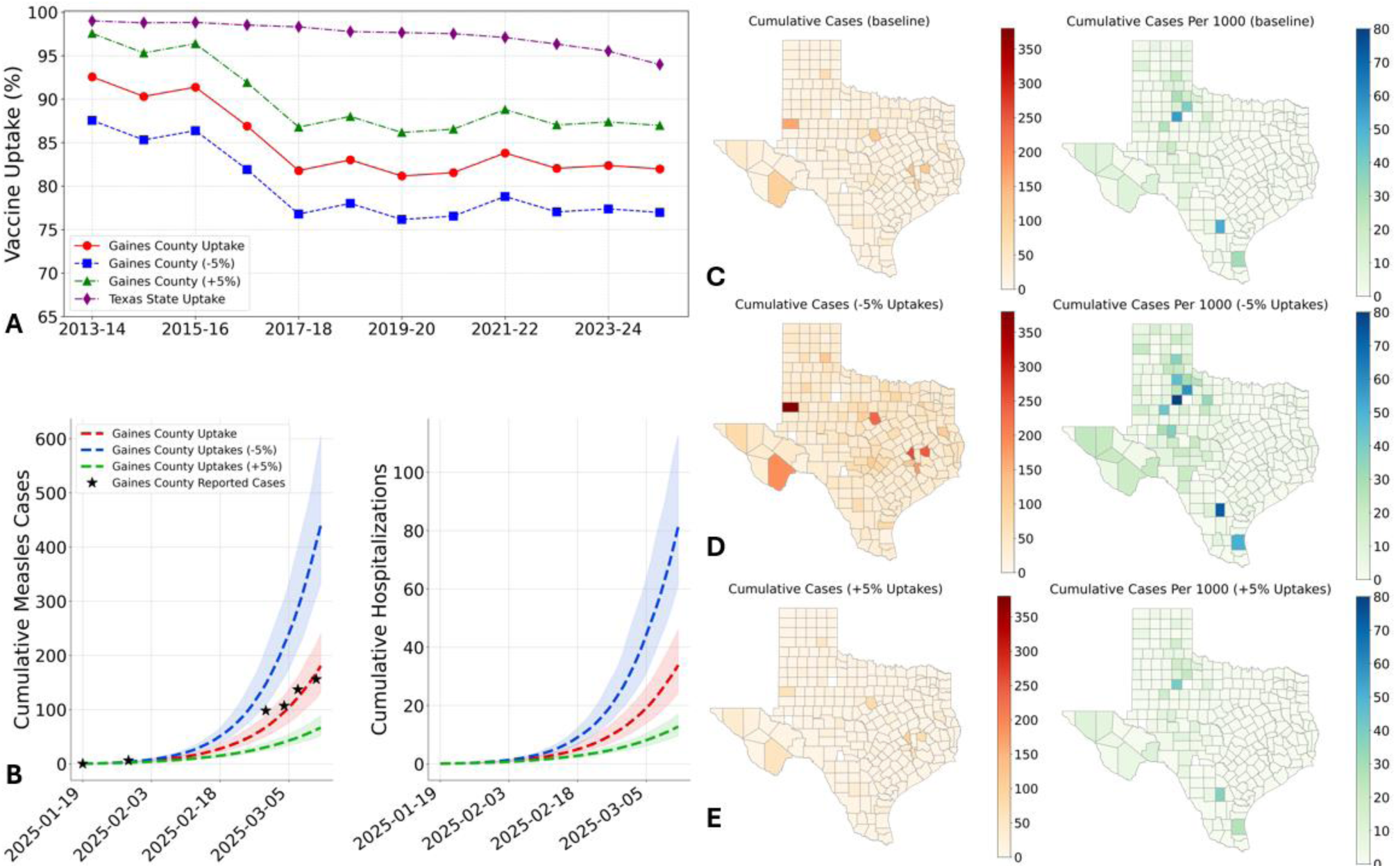
Vaccination rates and counterfactual scenario modeling projections for the 2025 measles outbreak in Gaines County and Texas. (A) Vaccination rates for kindergarten students from 2013 to 2025 in Gaines County and Texas. (B) Counterfactual scenario projection results for Gaines County. The estimated reported measles cases and hospitalizations are shown for the baseline scenario (red), an elevated vaccination scenario assuming vaccine coverage increased by 5% from the reported levels (green), and a depressed vaccination scenario with coverage reduced by 5% from the reported levels (blue). Dashed lines and shaded ribbons represent the median and 95% estimation intervals across 200 stochastic simulations, respectively. Black stars indicate reported measles cases. (C), (D), and (E) show heatmaps of the mean cumulative reported cases and cases per 1000 population from the simulations under the baseline, 5% reduced, and 5% improved vaccination coverage scenarios. The simulations were run from January 20 to March 11, 2025, with mean results generated from 200 stochastic simulations.

### Method

We developed a mathematical model (**Figure S.1**), first calibrating it using reported case data from Gaines County and local vaccination rates [4,6]. Expanding our analysis, we simulated a hypothetical measles case importation on January 20, 2025—coinciding with the introduction of patient zero in Gaines County—to estimate potential outbreak sizes across various Texas counties under three scenarios: (a) reported vaccination rates for each county, referred to as baseline, (b) a 5% increase in reported vaccination coverage, and (c) a 5% reduction in reported vaccination coverage for each county.

### Result

Under the baseline scenario across all Texas counties, Gaines county (**Figures 1.B**) along with Brazos (128 cases, 95% CI: 101–154), Walker (117 cases, 95% CI: 91–150), Erath (114 cases, 95% CI: 93–143), and Brewster (100 cases, 95% CI: 82–118) counties were each projected to report over 100 cases. Moreover, Kent (56 cases per 1,000 population, 95% CI: 51–61) and McMullen (52 cases per 1,000 population, 95% CI: 47–56) counties were estimated to experience outbreaks exceeding 50 cases per 1,000 population (**Figures 1.C**).

Under the 5% reduction scenario, outbreak magnitudes increased substantially. Gaines (375 cases, 95% CI 294-502), Brazos (260 cases, 95% CI: 195–344), Walker (247 cases, 95% CI: 197–310), and Erath (235 cases, 95% CI: 189-301) counties were each projected to exceed 200 reported cases. Additionally, Kent (79 cases per 1,000 population, 95% CI: 72–86), King (59 cases per 1,000 population, 95% CI: 55–64), and McMullen (73 cases per 1,000 population, 95% CI: 67–78) counties were projected to experience more than 50 cases per 1,000 population (**Figures 1.D**). In contrast, if all counties increased their current vaccination rates by 5%, no county was expected to exceed 100 reported cases in total or 50 cases per 1,000 population (**Figures 1.E**).

## Discussion

Our findings highlight the critical role of improving MMR vaccination coverage to prevent large-scale measles outbreaks, particularly in regions with declining immunization rates. Projecting potential measles outbreaks that have not yet emerged requires making assumptions about case importation, transmission dynamics, undocumented vaccination rates in certain age groups, and other factors that remain incompletely understood—particularly the complex behaviors of unvaccinated populations during an outbreak. Nonetheless, even if some assumptions are later disproven, simulating a range of carefully constructed scenarios can contribute significantly towards pandemic preparedness. Our modeling framework is readily adaptable to other locations with sufficient epidemiological and vaccination data beyond Texas.

## Data Availability

All data produced are available online at github

https://github.com/bikaiming93/measles_model

## Acknowledgements

This work is supported by the Texas Epidemic Public Health Institute (TEPHI).

## Appendix

### Data

#### Census Data

Age-specific population data for Texas counties were obtained from the 2023 Annual County and Puerto Rico Municipio Resident Population Estimates by Single Year of Age and Sex, as reported by the United States Census Bureau [7].

#### Measles Case Data

The case data were obtained from the Texas Department of State Health Services (DSHS) [4]. Beginning February 28, 2025, DSHS issued biweekly measles outbreak briefs every Tuesday and Friday.

#### Measles Case and Hospitalization age distributions

The CDC reports the age breakdowns for measles cases and hospitalizations, updating the data on a weekly basis [8].

#### Measles vaccination rates

DSHS provided annual MMR vaccination rates for kindergarten students across all Texas counties from the 2013–2014 to 2024–2025 school years [9][6].

### Model Description

We employed an age-structured SEIR (Susceptible-Exposed-Infectious-Recovered) model to simulate measles transmission dynamics, incorporating health outcomes such as hospitalizations and deaths (**Figure S.1**). Vaccination efficacy was represented using a “leaky” model, where vaccinated individuals have reduced susceptibility rather than complete immunity [10]. The population was segmented into three age groups—0-4, 5-19, and 20+ years—to align with the age categories reported by the CDC [8]. For age group *i*, the age-specific transitions among disease compartments as given by:

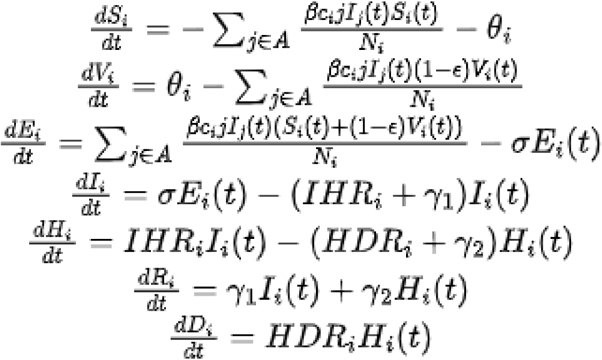

where *S_i_, V_i_, E_i_, I_i_, R_i_, H_i_, Di* are age-specific numbers of people who are in the susceptible, vaccinated, exposed, infectious, recovered, hospitalized and death compartments, respectively. β represents the time-dependent transmission rate; *c_i,j_* represents the number daily contacts between age group *i* to *j*; *c* represents the vaccine efficacy, θ_i_ represents the daily vaccinated population in the age group *i*; σ represents the exposed to infectious rate; *IHR_i_* represents the infectious to hospitalization rate in age group *i*; *HDR_i_* represents the hospitalization to death rate in age group *i*; γ_1_ and γ_2_ represent the recovery rates from the infectious and hospitalizations. *A* is the set of the age groups, which include 0-4, 5-19, and 20+ years.

We calibrate the transmission rate β of the baseline model to the reported case using the least squares fitting. The fitting period spans from January 20, 2025, to March 10, 2025. For each scenario projection, we run 200 stochastic simulations. We compute the projected measles cases, hospitalizations and deaths and summarize their evolving distributions using the 0.025, 0.50, and 0.975 quantiles for each day.

**Figure S.1.**
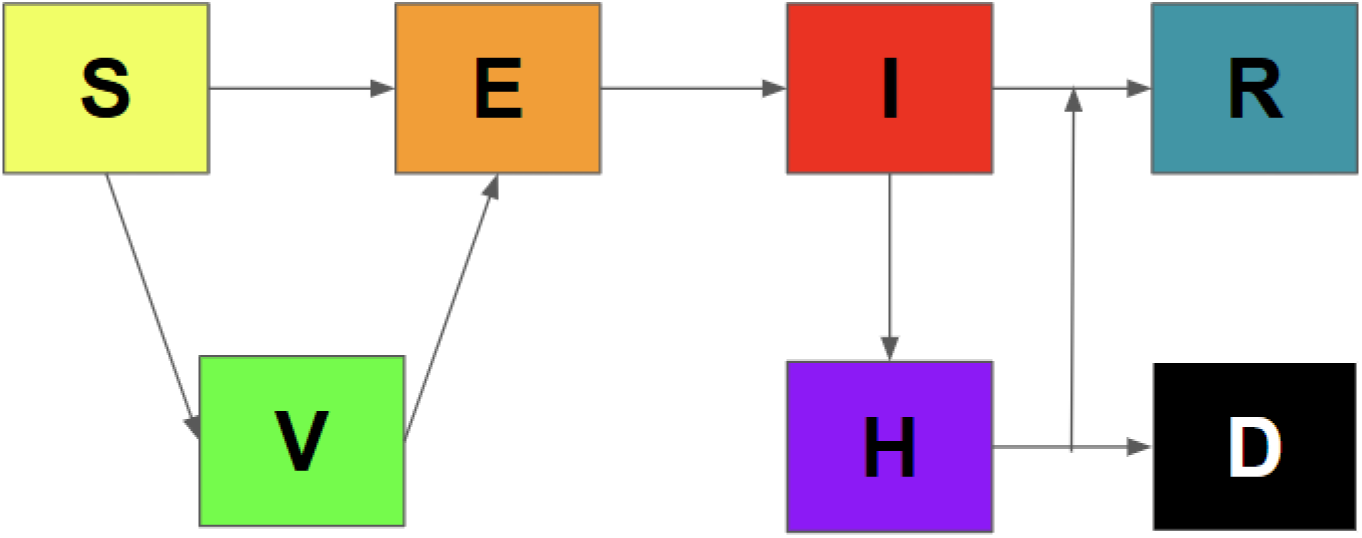
Schematic representation of measles transmission model.

### Model Assumption

To implement the counterfactual projections, we made the following assumptions:

#### Vaccine efficacy (VE)

Vaccinated individuals experience reduced infection rates on a per-exposure basis, determined by the corresponding VE for one-dose or two-dose MMR vaccination[8].

#### Vaccine uptakes

Kindergarten MMR vaccination rates from the 2013–2014 to 2024–2025 school years[6] were used to estimate uptake for children aged 5 to 16 (for example., the 2024–2025 rate represents current 5-year-olds, 2023–2024 for current 6-year-olds, and so on.). For other age groups, uptake is assumed to equal the county-specific average kindergarten rate over this period.

#### Newly vaccinated population during simulation

Children aged 12–15 months are assumed to receive their first MMR dose[11], uniformly distributed throughout the year. Kindergarten-aged children received their second dose during the back-to-school period in August 2024 [12], so no second doses occur during the simulation period (January 20 to March 10, 2025).

#### Total Populations

Using the 2023 census data as the most recent source [7], we assumed the age distribution in 2025 is identical to that of 2023 for each county. The total population is considered stable during the simulation period, with natural births equaling natural deaths, and age progression is ignored given the period is less than one year.

#### Contact between age groups

Contacts between age groups follow the MOBS contact matrix[13], with both vaccinated and unvaccinated individuals within each age group having equal probability of contacting an infectious case.

Other assumptions: Asymptomatic and unreported measles cases are excluded due to limited data. All hospitalizations are assumed to occur among infectious individuals, and all deaths are assumed to arise from these hospitalizations.

### Model Parameters

We present the parameter values and age-specific contact matrix in **Tables S.1** and **S.2**, respectively. The 3×3 contact matrix in **Table S.2** was aggregated from an 85×85 matrix provided by the MOBS Lab by consolidating ages into three groups: 0–4, 5–19, and 20+[13].

**Table S.1.** Parameter values for measles model.

| Parameter | Notation | Value | Reference |
| --- | --- | --- | --- |
| Transmission rate | $\beta$ | Calibrated to case data | [4] |
| Contact between age group $i$ and $j$ | $c_{i,j}$ | Contact matrix in **Table S.2** | [13] |
| Newly vaccinated population | $\theta_i$ | Derive from vaccine uptake | [9] |
| Vaccine efficacy | $\epsilon$ | 93% for 0-4 in first dose<br>97% for 5+ in second dose | [8,11] |
| Exposed to infectious rate | $\sigma$ | 1/7-1/14 daily rates | [14] |
| Recovery rate from infectious | $\gamma_1$ | 1/4-1/7 daily rates | [15] |
| Recovery rate from hospitalizations | $\gamma_2$ | 1/5-1/6 daily rates | [16] |
| Infectious to Hospitalization rate in age group $i$ | $IHR_i$ | Derive from 29%, 13%, 17% hospitalized in age groups | [8] |
| Hospitalization death rate | $HDR_i$ | Derive from 0.1-0.3% | [14] |

**Table S.2.**
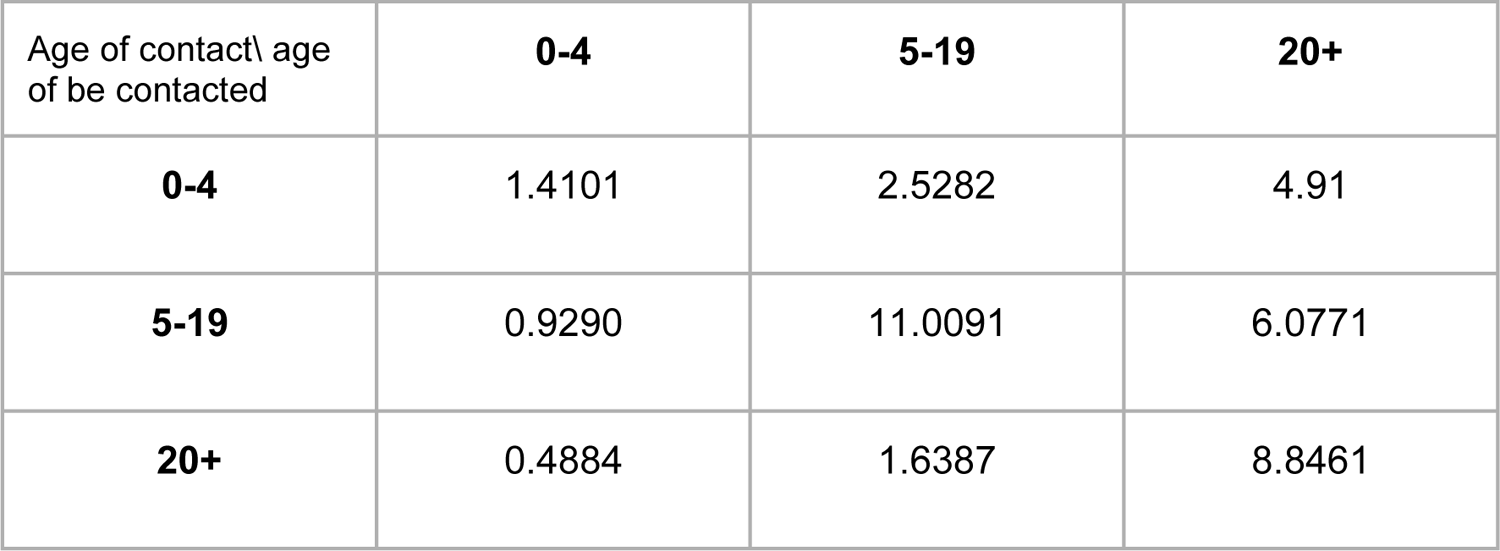
Age-specific contact matrix in Texas.

### Scenarios

#### Baseline scenario

For each county, we use the reported MMR vaccination rates and introduce one infectious case (patient zero) on January 20, 2025, and apply the transmission rate calibrated from Gaines County.

#### Reduced vaccination uptake scenario

For each county, we assume MMR vaccination rates are 5% lower than the reported rates across all three age groups, and introduce one infectious case on January 20, 2025, and apply the transmission rate calibrated from Gaines County.

#### Texas average vaccination uptake scenario

For each county, we apply the Texas average MMR vaccination rate for all three age groups, unless the county’s reported rate is higher—in which case the reported rate is retained—and introduce one infectious case on January 20, 2025, and apply the transmission rate calibrated from Gaines County.

### Results in hospitalizations

**Figure S.2.**
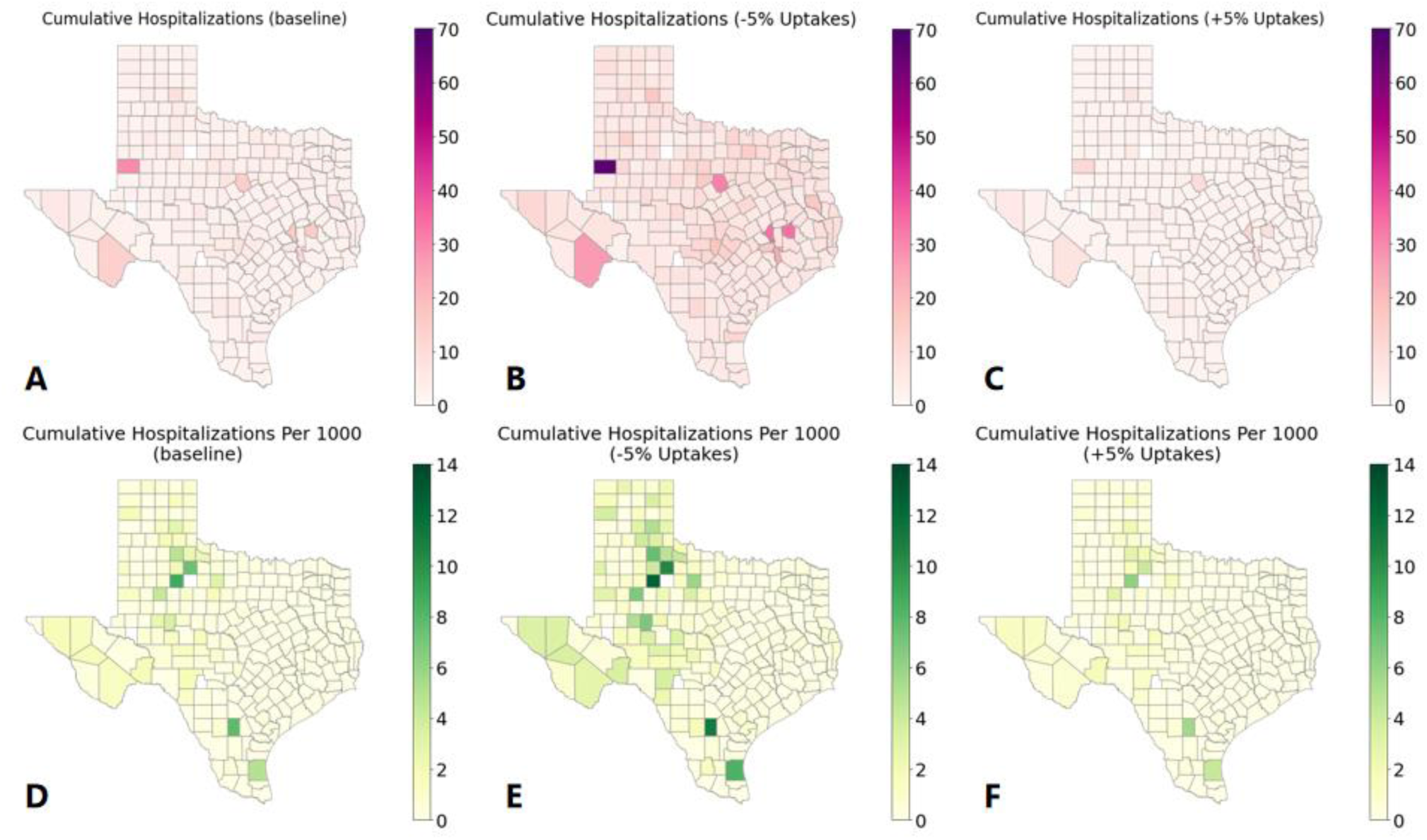
Counterfactual scenario modeling projections on hospitalizations for the 2025 measles outbreak in Texas. **(A)** and **(D)** display heatmaps of the mean cumulative reported hospitalizations and hospitalizations per 1,000 population under the reported coverage scenario. **(B)** and **(E)** present heatmaps of the same outcomes under the reduced vaccination uptake scenario. **(C)** and **(F)** illustrate heatmaps of the outcomes under the increased vaccination uptake scenario. The simulations were run from January 20 to March 11, 2025, with mean results generated from 200 stochastic simulations.

### Results in age-specific Cases

**Figure S.3.**
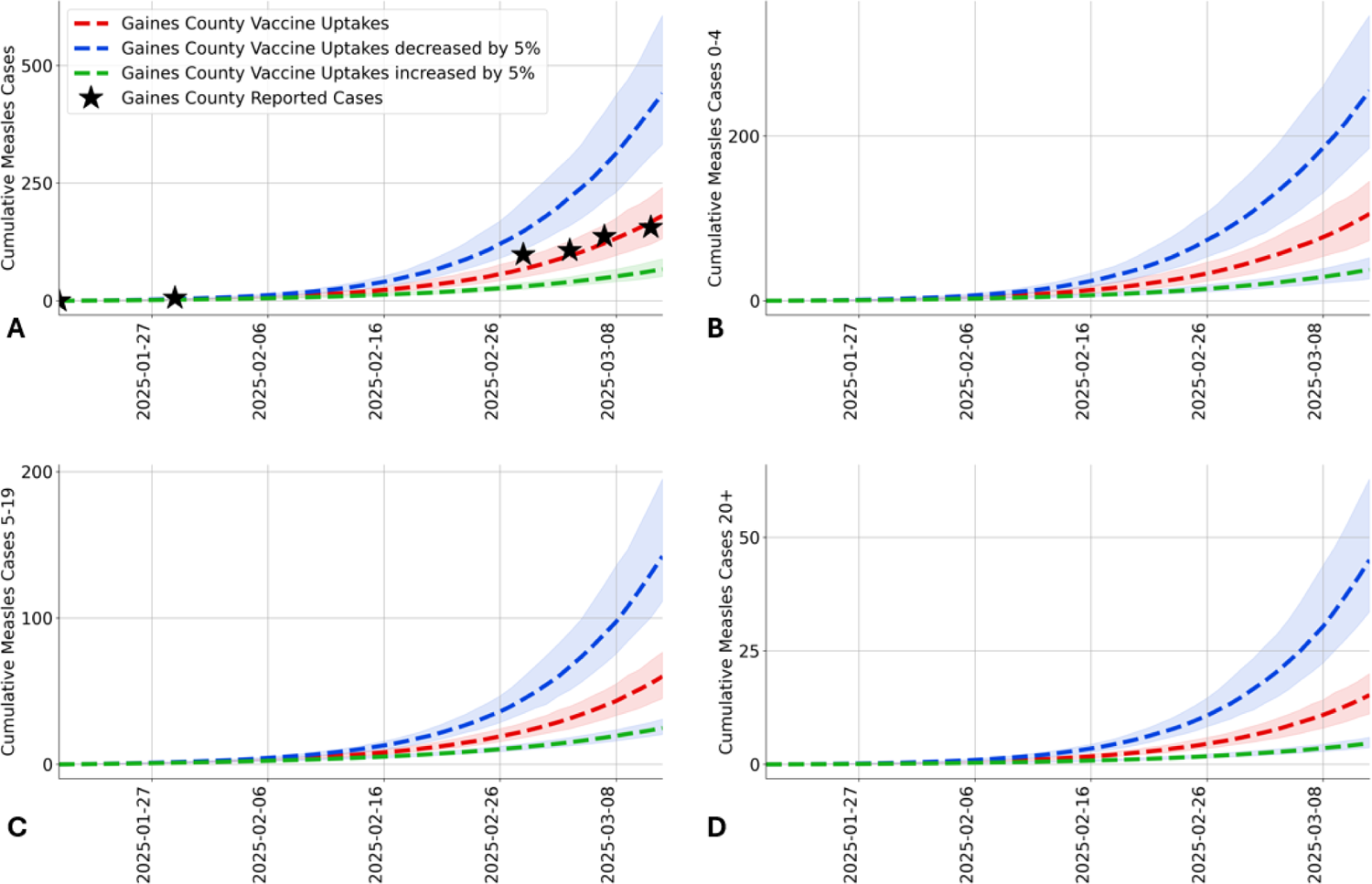
Counterfactual scenario projection results for Gaines County. The estimated reported measles cases in total **(A)**, 0-4 age group **(B)**, 5-19 age group **(C)**, and 20+ age group **(D)** are shown for the baseline scenario (red), an increased vaccination scenario with coverage boosted by 5% from the reported levels (green), and a depressed vaccination scenario with coverage reduced by 5% from the reported levels (blue). Dashed lines and shaded ribbons represent the median and 95% estimation intervals across 200 stochastic simulations, respectively. Black stars indicate reported measles cases.

**Figure S.4.**
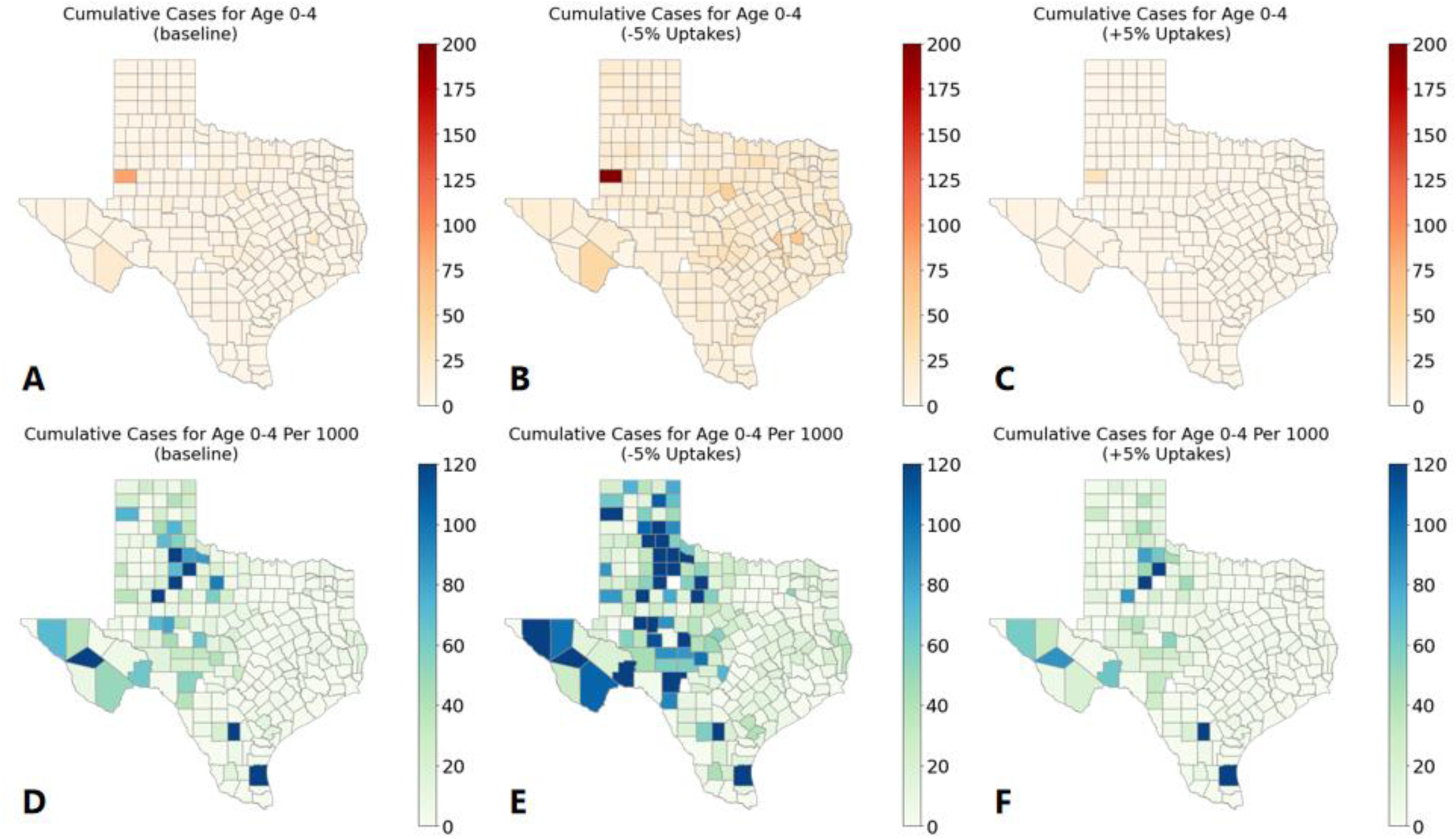
Counterfactual scenario modeling projections on reported cases from 0 to 4 age group for the 2025 measles outbreak in Texas. **(A)** and **(D)** display heatmaps of the mean cumulative reported hospitalizations and hospitalizations per 1,000 population under the reported coverage scenario. **(B)** and **(E)** present heatmaps of the same outcomes under the reduced vaccination uptake scenario. **(C)** and **(F)** illustrate heatmaps of the outcomes under the increased vaccination uptake scenario. The simulations were run from January 20 to March 11, 2025, with mean results generated from 200 stochastic simulations.

**Figure S.5.**
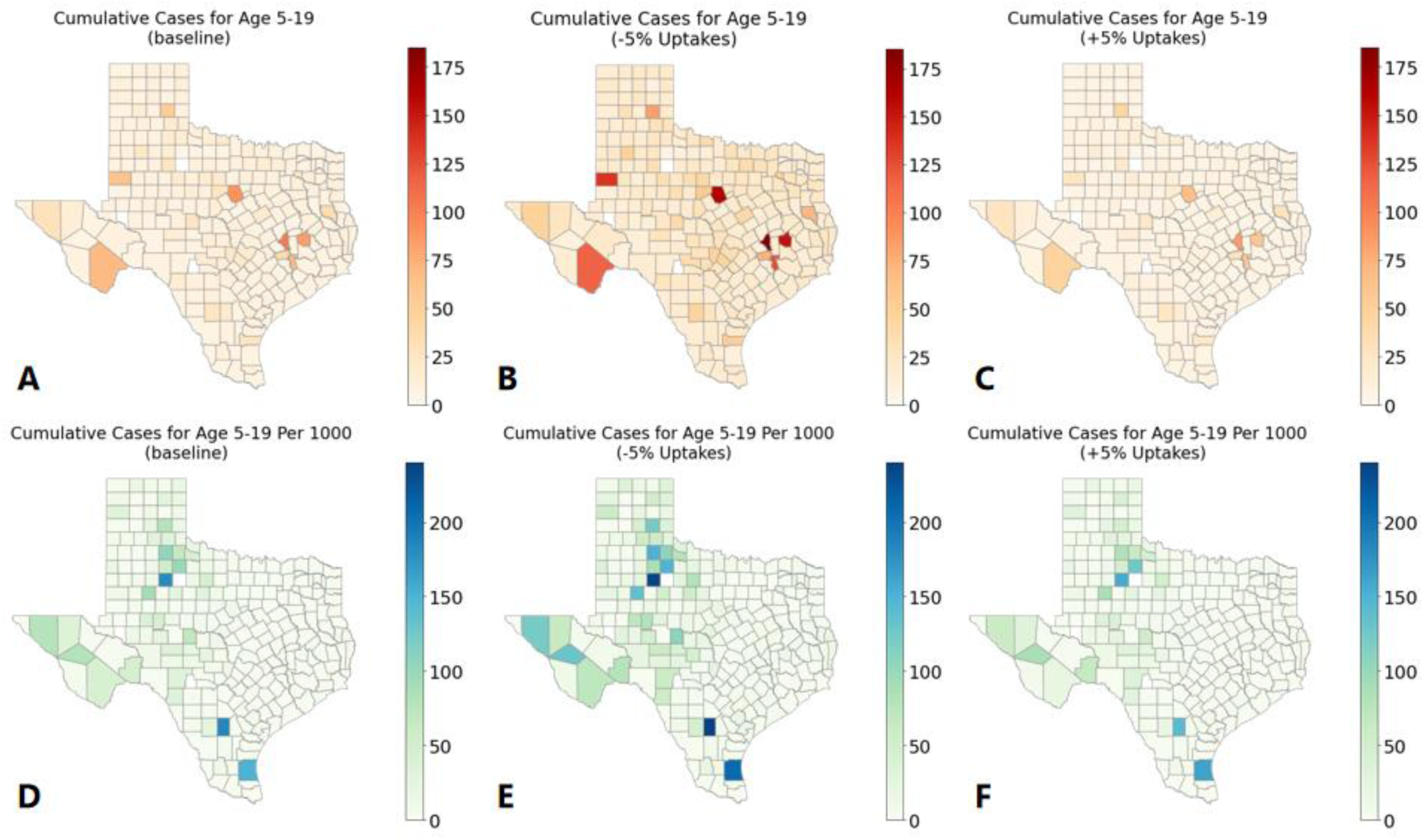
Counterfactual scenario modeling projections on reported cases from 0 to 4 age group for the 2025 measles outbreak in Texas. **(A)** and **(D)** display heatmaps of the mean cumulative reported hospitalizations and hospitalizations per 1,000 population under the reported coverage scenario. **(B)** and **(E)** present heatmaps of the same outcomes under the reduced vaccination uptake scenario. **(C)** and **(F)** illustrate heatmaps of the outcomes under the increased vaccination uptake scenario. The simulations were run from January 20 to March 11, 2025, with mean results generated from 200 stochastic simulations.

**Figure S.6.**
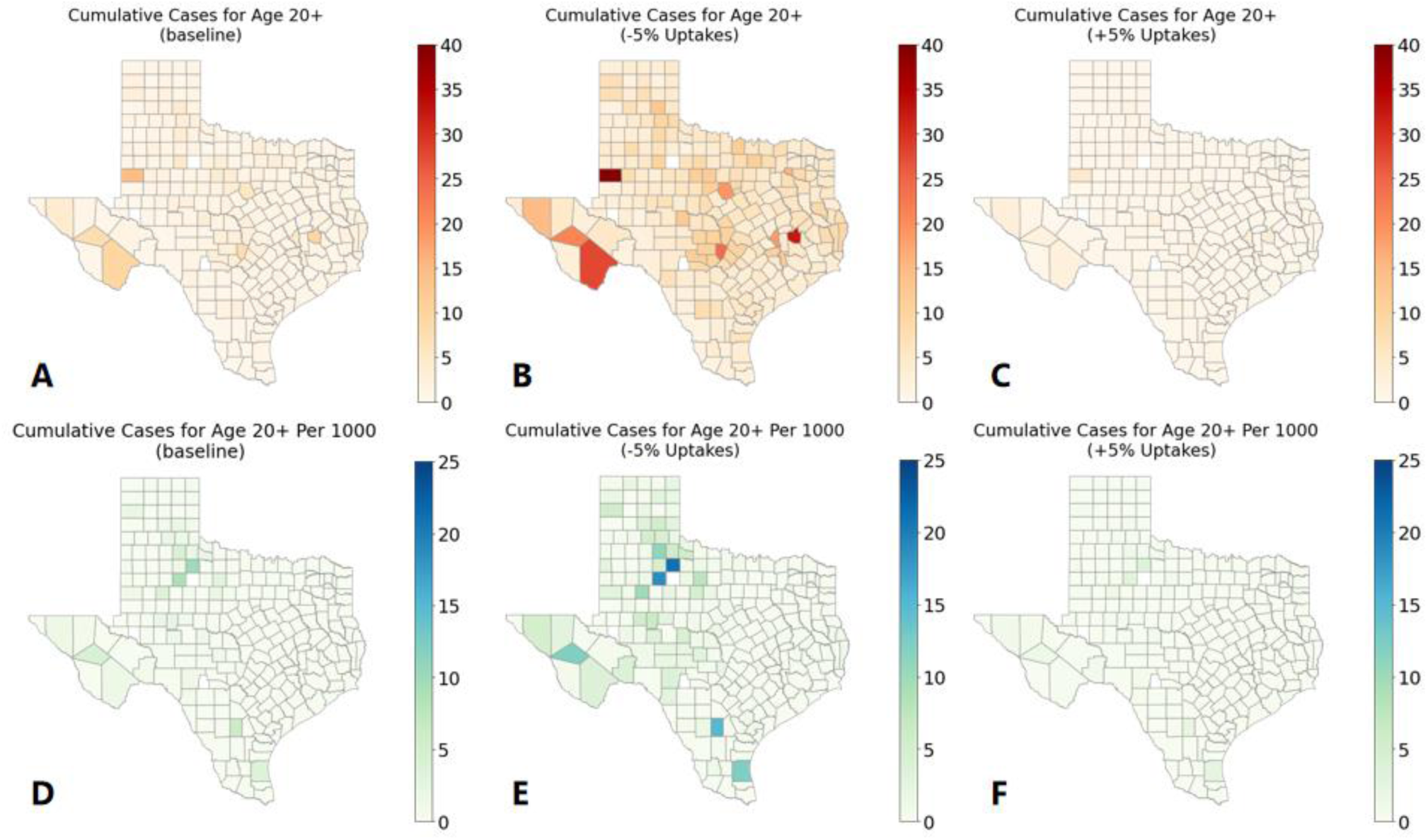
Counterfactual scenario modeling projections on reported cases from 20+ age group for the 2025 measles outbreak in Texas. **(A)** and **(D)** display heatmaps of the mean cumulative reported hospitalizations and hospitalizations per 1,000 population under the reported coverage scenario. **(B)** and **(E)** present heatmaps of the same outcomes under the reduced vaccination uptake scenario. **(C)** and **(F)** illustrate heatmaps of the outcomes under the increased vaccination uptake scenario. The simulations were run from January 20 to March 11, 2025, with mean results generated from 200 stochastic simulations.

### Results in age-specific Hospitalizations

**Figure S.7.**
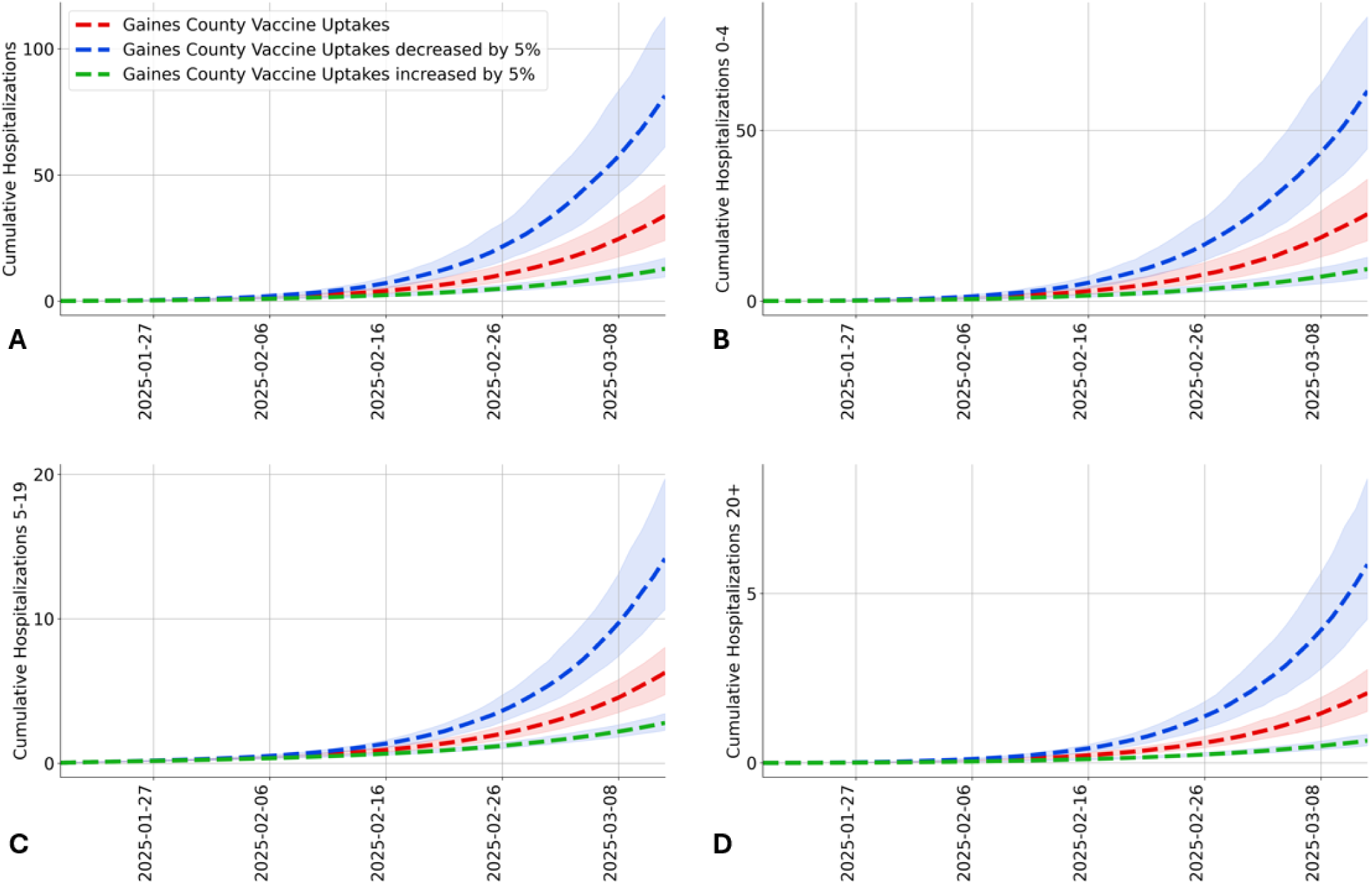
Counterfactual scenario projection results for Gaines County. The estimated reported measles hospitalizations in total **(A)**, 0-4 age group **(B)**, 5-19 age group **(C)**, and 20+ age group **(D)** are shown for the baseline scenario (red), an increased vaccination scenario with coverage boosted by 5% from the reported levels (green), and a depressed vaccination scenario with coverage reduced by 15% from the reported levels (blue). Dashed lines and shaded ribbons represent the median and 95% estimation intervals across 200 stochastic simulations, respectively.

**Figure S.8.**
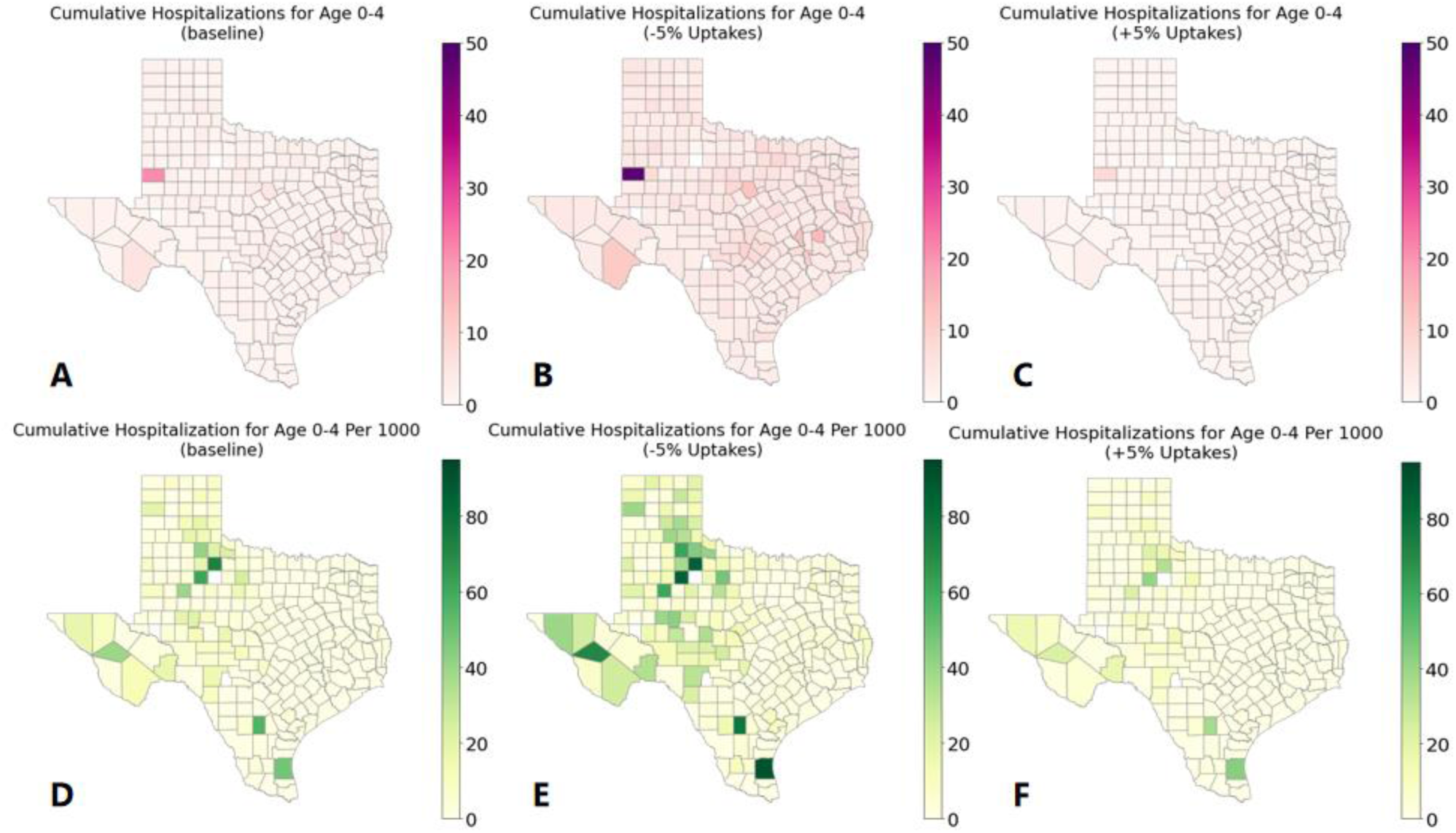
Counterfactual scenario modeling projections on reported hospitalizations from 0 to 4 age group for the 2025 measles outbreak in Texas. **(A)** and **(D)** display heatmaps of the mean cumulative reported hospitalizations and hospitalizations per 1,000 population under the reported coverage scenario. **(B)** and **(E)** present heatmaps of the same outcomes under the reduced vaccination uptake scenario. **(C)** and **(F)** illustrate heatmaps of the outcomes under the increased vaccination uptake scenario. The simulations were run from January 20 to March 11, 2025, with mean results generated from 200 stochastic simulations.

**Figure S.9.**
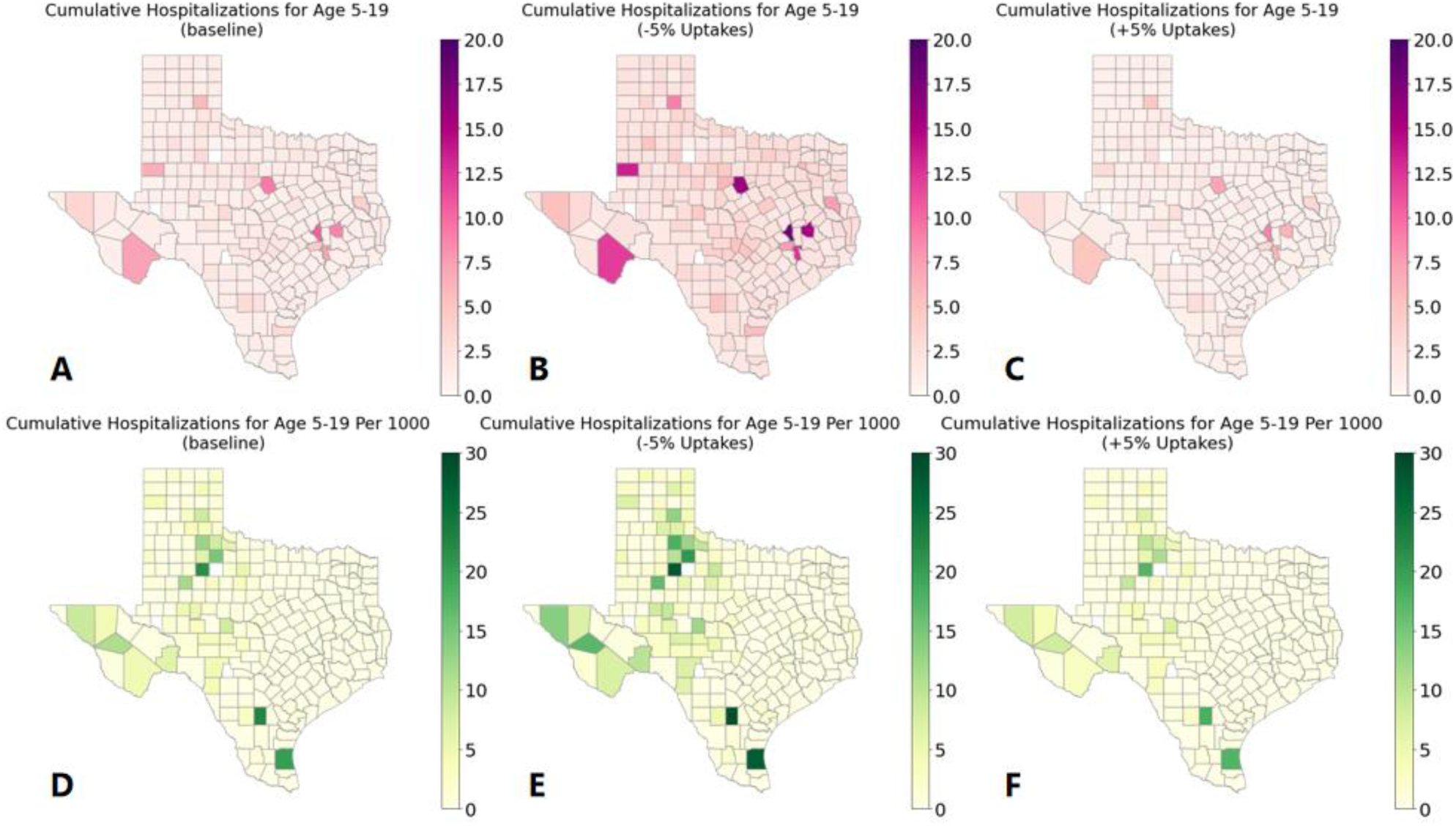
Counterfactual scenario modeling projections on reported hospitalizations from 5 to 19 age group for the 2025 measles outbreak in Texas. **(A)** and **(D)** display heatmaps of the mean cumulative reported hospitalizations and hospitalizations per 1,000 population under the reported coverage scenario. **(B)** and **(E)** present heatmaps of the same outcomes under the reduced vaccination uptake scenario. **(C)** and **(F)** illustrate heatmaps of the outcomes under the increased vaccination uptake scenario. The simulations were run from January 20 to March 11, 2025, with mean results generated from 200 stochastic simulations.

**Figure S.10.**
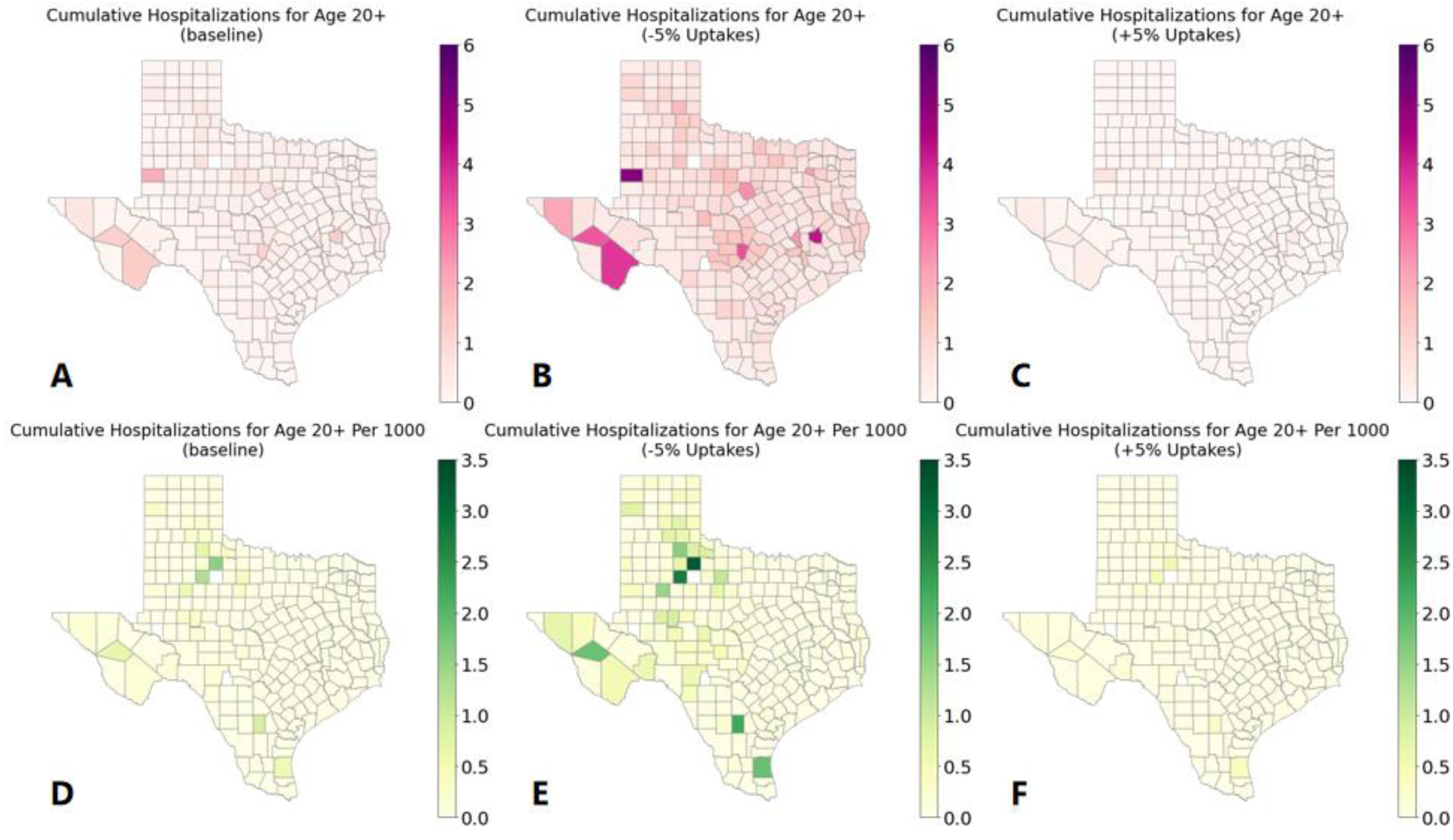
Counterfactual scenario modeling projections on reported hospitalizations from 20+ age group for the 2025 measles outbreak in Texas. **(A)** and **(D)** display heatmaps of the mean cumulative reported hospitalizations and hospitalizations per 1,000 population under the reported coverage scenario. **(B)** and **(E)** present heatmaps of the same outcomes under the reduced vaccination uptake scenario. **(C)** and **(F)** illustrate heatmaps of the outcomes under the increased vaccination uptake scenario. The simulations were run from January 20 to March 11, 2025, with mean results generated from 200 stochastic simulations.

### Tables summarize all the results

**Table S.3.** Age-specific cumulative case counts and hospitalizations derived from baseline scenario projections.

| County Name | Total Cases | 0-4 Cases | 5-19 Cases | 20+ Cases | Total Hosp | 0-4 Hosp | 5-19 Hosp | 20+ Hosp |
| --- | --- | --- | --- | --- | --- | --- | --- | --- |
| Anderson | 14.79<br>[12.67, 17.83] | 4.79<br>[3.95, 5.95] | 8.67<br>[7.52, 10.22] | 1.27<br>[1.08, 1.51] | 2.53<br>[2.15, 3.06] | 1.25<br>[1.01, 1.58] | 1.09<br>[0.95, 1.28] | 0.19<br>[0.16, 0.23] |
| Andrews | 9.71<br>[8.54, 11.34] | 3.25<br>[2.74, 3.9] | 6.07<br>[5.41, 6.93] | 0.42<br>[0.37, 0.49] | 1.73<br>[1.52, 2.02] | 0.86<br>[0.71, 1.02] | 0.82<br>[0.72, 0.94] | 0.06<br>[0.05, 0.08] |
| Angelina | 15.79<br>[13.27, 18.87] | 5.2<br>[4.29, 6.3] | 9.53<br>[8.08, 11.27] | 1<br>[0.84, 1.21] | 2.68<br>[2.24, 3.24] | 1.34<br>[1.1, 1.66] | 1.19<br>[1, 1.43] | 0.15<br>[0.12, 0.18] |
| Aransas | 12.43<br>[10.57, 14.57] | 3.81<br>[3.04, 4.55] | 7.54<br>[6.53, 8.71] | 1.12<br>[0.95, 1.33] | 2.14<br>[1.78, 2.54] | 1<br>[0.79, 1.21] | 0.97<br>[0.83, 1.15] | 0.17<br>[0.14, 0.21] |
| Archer | 10.89<br>[9.43, 12.57] | 3.35<br>[2.81, 3.91] | 6.68<br>[5.94, 7.7] | 0.84<br>[0.73, 0.97] | 1.9<br>[1.62, 2.22] | 0.88<br>[0.73, 1.04] | 0.88<br>[0.78, 1.03] | 0.13<br>[0.11, 0.15] |
| Armstrong | 13.61<br>[11.6, 15.61] | 4.95<br>[4.09, 5.9] | 7.33<br>[6.34, 8.33] | 1.31<br>[1.12, 1.52] | 2.46<br>[2.04, 2.91] | 1.31<br>[1.08, 1.6] | 0.96<br>[0.82, 1.09] | 0.2<br>[0.16, 0.24] |
| Atascosa | 10.98<br>[9.51, 13.28] | 3.25<br>[2.69, 4.16] | 7.23<br>[6.27, 8.48] | 0.53<br>[0.45, 0.64] | 1.87<br>[1.59, 2.27] | 0.85<br>[0.7, 1.09] | 0.94<br>[0.8, 1.11] | 0.08<br>[0.07, 0.1] |
| Austin | 13.92<br>[11.67, 16.57] | 4.66<br>[3.75, 5.76] | 8.25<br>[6.96, 9.71] | 1.08<br>[0.89, 1.28] | 2.41<br>[2.01, 2.9] | 1.2<br>[0.97, 1.5] | 1.04<br>[0.9, 1.26] | 0.16<br>[0.13, 0.2] |
| Bailey | 14.1<br>[11.93, 16.4] | 4.36<br>[3.59, 5.22] | 9.09<br>[7.79, 10.48] | 0.56<br>[0.47, 0.65] | 2.38<br>[2.02, 2.82] | 1.14<br>[0.95, 1.38] | 1.16<br>[0.99, 1.35] | 0.08<br>[0.07, 0.1] |
| Bandera | 17.09<br>[14.46, 20.62] | 5.5<br>[4.48, 6.89] | 9.4<br>[8.08, 11.14] | 2.14<br>[1.8, 2.58] | 2.92<br>[2.42, 3.51] | 1.42<br>[1.14, 1.83] | 1.18<br>[1, 1.39] | 0.31<br>[0.27, 0.38] |
| Bastrop | 16.09<br>[13.51, 18.69] | 5.3<br>[4.3, 6.38] | 9.7<br>[8.21, 11.14] | 1.05<br>[0.88, 1.23] | 2.73<br>[2.32, 3.21] | 1.37<br>[1.09, 1.64] | 1.21<br>[1.03, 1.4] | 0.16<br>[0.13, 0.18] |
| Baylor | 11.82<br>[10.08, 13.41] | 3.86<br>[3.16, 4.55] | 7.04<br>[6.02, 7.99] | 0.88<br>[0.74, 1.01] | 2.08<br>[1.76, 2.42] | 1.01<br>[0.83, 1.22] | 0.92<br>[0.8, 1.06] | 0.13<br>[0.11, 0.15] |
| Bee | 17.81<br>[15.04, 21.23] | 4.42<br>[3.6, 5.4] | 12.29<br>[10.34, 14.56] | 1.04<br>[0.86, 1.25] | 2.8<br>[2.31, 3.33] | 1.14<br>[0.92, 1.43] | 1.5<br>[1.25, 1.73] | 0.15<br>[0.13, 0.19] |
| Bell | 14.18<br>[11.98, 16.8] | 5.14<br>[4.23, 6.36] | 8.27<br>[7.07, 9.7] | 0.8<br>[0.67, 0.95] | 2.49<br>[2.07, 3.01] | 1.33<br>[1.07, 1.66] | 1.05<br>[0.91, 1.24] | 0.12<br>[0.1, 0.14] |
| Bexar | 16.33<br>[13.66, 18.91] | 4.96<br>[4.17, 6.04] | 10.37<br>[8.86, 12.01] | 0.97<br>[0.81, 1.13] | 2.73<br>[2.26, 3.2] | 1.29<br>[1.06, 1.57] | 1.29<br>[1.08, 1.52] | 0.14<br>[0.12, 0.17] |
| Blanco | 45.43<br>[38.12, 53.94] | 14.81<br>[11.98, 17.97] | 22.99<br>[19.32, 27.28] | 7.82<br>[6.56, 9.4] | 7.41<br>[6.14, 8.82] | 3.71<br>[3.01, 4.48] | 2.58<br>[2.17, 3.11] | 1.1<br>[0.92, 1.36] |
| Borden | 14.93<br>[13.49, 16.62] | 3.49<br>[3.07, 3.99] | 9.85<br>[8.93, 10.98] | 1.61<br>[1.41, 1.83] | 2.44<br>[2.15, 2.7] | 0.93<br>[0.81, 1.07] | 1.26<br>[1.13, 1.42] | 0.24<br>[0.21, 0.28] |
| Bosque | 14.92<br>[12.76, 17.36] | 4.86<br>[4.11, 5.97] | 8.6<br>[7.46, 9.9] | 1.44<br>[1.24, 1.68] | 2.59<br>[2.13, 3.06] | 1.27<br>[1.04, 1.57] | 1.09<br>[0.94, 1.26] | 0.22<br>[0.18, 0.26] |
| Bowie | 15.51<br>[13.2, 18.45] | 5.2<br>[4.3, 6.32] | 9.19<br>[7.94, 10.85] | 1.18<br>[1, 1.41] | 2.67<br>[2.25, 3.22] | 1.34<br>[1.1, 1.67] | 1.15<br>[0.99, 1.37] | 0.18<br>[0.15, 0.21] |
| Brazoria | 12.37<br>[10.91, 14.65] | 4.01<br>[3.28, 4.88] | 7.62<br>[6.81, 8.95] | 0.78<br>[0.69, 0.93] | 2.15<br>[1.86, 2.55] | 1.05<br>[0.85, 1.29] | 0.98<br>[0.87, 1.14] | 0.12<br>[0.1, 0.14] |
| Brazos | 125.84<br>[98.85, 154.42] | 19.35<br>[15.62, 23.97] | 101.85<br>[79.98, 126.03] | 4.69<br>[3.66, 5.8] | 15.51<br>[12.41, 19.12] | 4.6<br>[3.63, 5.93] | 10.32<br>[8.31, 12.83] | 0.63<br>[0.5, 0.8] |
| Brewster | 100.88<br>[83.81, 120.33] | 21.65<br>[18.18, 26.05] | 69.4<br>[57.87, 82.81] | 9.59<br>[7.93, 11.66] | 13.77<br>[11.58, 17.04] | 5.28<br>[4.32, 6.59] | 7.25<br>[6.04, 8.94] | 1.29<br>[1.06, 1.63] |
| Briscoe | 15.18<br>[12.98, 17.87] | 5.4<br>[4.48, 6.4] | 7.92<br>[6.86, 9.18] | 1.85<br>[1.55, 2.19] | 2.72<br>[2.29, 3.26] | 1.43<br>[1.18, 1.72] | 1.02<br>[0.88, 1.21] | 0.27<br>[0.23, 0.34] |
| Brooks | 10.6<br>[9.36, 12.06] | 3.11<br>[2.67, 3.7] | 7.02<br>[6.19, 7.99] | 0.5<br>[0.44, 0.57] | 1.81<br>[1.56, 2.12] | 0.82<br>[0.69, 0.98] | 0.92<br>[0.79, 1.08] | 0.08<br>[0.07, 0.09] |
| Brown | 23.42<br>[19.96, 28.19] | 7.12<br>[6, 8.77] | 14.4<br>[12.32, 17.09] | 1.85<br>[1.55, 2.22] | 3.77<br>[3.17, 4.62] | 1.81<br>[1.5, 2.23] | 1.71<br>[1.44, 2.04] | 0.27<br>[0.22, 0.33] |
| Burleson | 8.66<br>[7.41, 9.99] | 2.79<br>[2.31, 3.33] | 5.35<br>[4.64, 6.15] | 0.54<br>[0.46, 0.62] | 1.56<br>[1.3, 1.84] | 0.74<br>[0.6, 0.89] | 0.73<br>[0.63, 0.86] | 0.08<br>[0.07, 0.1] |
| Burnet | 24.52<br>[20.59, 29.76] | 8.72<br>[6.95, 11.13] | 13.08<br>[11.02, 15.67] | 2.78<br>[2.32, 3.34] | 4.19<br>[3.45, 5.08] | 2.22<br>[1.79, 2.77] | 1.57<br>[1.31, 1.93] | 0.41<br>[0.34, 0.5] |
| Caldwell | 20.91<br>[17.16, 23.76] | 6.35<br>[5.17, 7.45] | 13.23<br>[11.05, 15.11] | 1.34<br>[1.1, 1.54] | 3.43<br>[2.84, 3.93] | 1.62<br>[1.31, 1.92] | 1.59<br>[1.34, 1.84] | 0.2<br>[0.16, 0.23] |
| Calhoun | 12.37<br>[10.75, 14.34] | 3.66<br>[3.02, 4.36] | 8.06<br>[7.06, 9.36] | 0.68<br>[0.59, 0.78] | 2.08<br>[1.77, 2.54] | 0.96<br>[0.78, 1.18] | 1.03<br>[0.9, 1.22] | 0.1<br>[0.09, 0.12] |
| Callahan | 15.31<br>[13.41, 18.35] | 5.1<br>[4.36, 6.2] | 8.74<br>[7.65, 10.51] | 1.48<br>[1.28, 1.8] | 2.66<br>[2.29, 3.2] | 1.32<br>[1.12, 1.65] | 1.1<br>[0.96, 1.31] | 0.22<br>[0.19, 0.27] |
| Cameron | 11.83<br>[10.15, 14.02] | 3.18<br>[2.63, 3.84] | 8.15<br>[7.1, 9.64] | 0.46<br>[0.39, 0.54] | 1.96<br>[1.67, 2.32] | 0.84<br>[0.69, 1.02] | 1.04<br>[0.9, 1.22] | 0.07<br>[0.06, 0.08] |
| Camp | 11.54<br>[10.02, 13.43] | 3.9<br>[3.28, 4.63] | 7.09<br>[6.17, 8.16] | 0.57<br>[0.49, 0.67] | 2.04<br>[1.73, 2.41] | 1.02<br>[0.85, 1.24] | 0.93<br>[0.8, 1.07] | 0.09<br>[0.07, 0.1] |
| Carson | 19.69<br>[16.73, 23.53] | 6.17<br>[5.15, 7.65] | 11.62<br>[9.85, 13.6] | 1.99<br>[1.67, 2.38] | 3.3<br>[2.77, 3.96] | 1.59<br>[1.29, 1.98] | 1.42<br>[1.21, 1.63] | 0.29<br>[0.25, 0.35] |
| Cass | 14.42<br>[12.01, 17.18] | 5.02<br>[4.05, 6.22] | 8.24<br>[6.93, 9.67] | 1.17<br>[0.97, 1.4] | 2.54<br>[2.08, 3.08] | 1.31<br>[1.04, 1.65] | 1.06<br>[0.88, 1.23] | 0.18<br>[0.14, 0.22] |
| Castro | 11.47<br>[9.86, 13.25] | 3.61<br>[2.96, 4.34] | 7.37<br>[6.43, 8.49] | 0.49<br>[0.42, 0.57] | 2<br>[1.66, 2.35] | 0.95<br>[0.76, 1.15] | 0.96<br>[0.83, 1.12] | 0.07<br>[0.06, 0.09] |
| Chambers | 14.1<br>[11.91, 17.07] | 4.84<br>[4, 6.05] | 8.37<br>[7.28, 9.76] | 0.93<br>[0.79, 1.11] | 2.45<br>[2.06, 2.98] | 1.26<br>[1.04, 1.56] | 1.07<br>[0.92, 1.25] | 0.14<br>[0.12, 0.17] |
| Cherokee | 10.3<br>[8.86, 12.24] | 3.33<br>[2.78, 4.09] | 6.44<br>[5.61, 7.55] | 0.54<br>[0.47, 0.65] | 1.81<br>[1.53, 2.18] | 0.87<br>[0.72, 1.09] | 0.85<br>[0.74, 1.01] | 0.08<br>[0.07, 0.1] |
| Childress | 22.71<br>[19.52, 26.78] | 6.6<br>[5.56, 7.97] | 13.83<br>[11.88, 16.3] | 2.25<br>[1.9, 2.68] | 3.68<br>[3.1, 4.38] | 1.7<br>[1.38, 2.07] | 1.65<br>[1.41, 1.94] | 0.33<br>[0.27, 0.4] |
| Clay | 13.79<br>[11.84, 16.76] | 4.3<br>[3.59, 5.42] | 8.28<br>[7.24, 9.87] | 1.21<br>[1.04, 1.47] | 2.37<br>[2.02, 2.87] | 1.12<br>[0.92, 1.43] | 1.06<br>[0.91, 1.27] | 0.18<br>[0.15, 0.22] |
| Cochran | 18.72<br>[15.78, 21.51] | 7.62<br>[6.47, 8.97] | 9.81<br>[8.3, 11.13] | 1.29<br>[1.07, 1.49] | 3.42<br>[2.87, 4] | 1.99<br>[1.68, 2.41] | 1.23<br>[1.02, 1.41] | 0.19<br>[0.16, 0.23] |
| Coke | 15.39<br>[13.27, 18] | 6.24<br>[5.26, 7.23] | 7.2<br>[6.2, 8.34] | 2<br>[1.68, 2.34] | 2.87<br>[2.42, 3.34] | 1.64<br>[1.37, 1.98] | 0.94<br>[0.81, 1.08] | 0.3<br>[0.24, 0.36] |
| Coleman | 16.13<br>[13.47, 18.88] | 5.58<br>[4.54, 6.81] | 9.04<br>[7.6, 10.55] | 1.5<br>[1.26, 1.76] | 2.82<br>[2.34, 3.34] | 1.46<br>[1.17, 1.75] | 1.14<br>[0.96, 1.34] | 0.22<br>[0.19, 0.27] |
| Collin | 23.52<br>[19.48, 28.14] | 7.69<br>[6.09, 9.54] | 13.85<br>[11.73, 16.52] | 1.97<br>[1.64, 2.38] | 3.89<br>[3.14, 4.76] | 1.95<br>[1.52, 2.45] | 1.64<br>[1.39, 1.99] | 0.29<br>[0.23, 0.35] |
| Collingsworth | 17.51<br>[15.33, 20.19] | 6.09<br>[5.15, 7.2] | 10<br>[8.73, 11.34] | 1.46<br>[1.25, 1.69] | 3.05<br>[2.6, 3.66] | 1.59<br>[1.31, 1.92] | 1.24<br>[1.08, 1.45] | 0.22<br>[0.18, 0.26] |
| Colorado | 9.77<br>[8.37, 11.66] | 3.5<br>[2.94, 4.26] | 5.67<br>[4.92, 6.6] | 0.63<br>[0.54, 0.75] | 1.79<br>[1.5, 2.16] | 0.92<br>[0.75, 1.14] | 0.77<br>[0.67, 0.92] | 0.1<br>[0.08, 0.12] |
| Comal | 19.55<br>[16.44, 23.63] | 6.73<br>[5.5, 8.38] | 10.92<br>[9.35, 12.96] | 1.92<br>[1.61, 2.32] | 3.35<br>[2.79, 4.12] | 1.73<br>[1.38, 2.2] | 1.34<br>[1.15, 1.6] | 0.28<br>[0.23, 0.35] |
| Comanche | 14.25<br>[12.15, 17.1] | 4.52<br>[3.71, 5.64] | 8.77<br>[7.55, 10.48] | 0.96<br>[0.82, 1.16] | 2.46<br>[2.02, 2.97] | 1.17<br>[0.95, 1.52] | 1.11<br>[0.92, 1.35] | 0.14<br>[0.12, 0.18] |
| Concho | 36.43<br>[31.7, 42.02] | 7.75<br>[6.48, 9.07] | 25.6<br>[22.36, 29.48] | 3.11<br>[2.68, 3.69] | 5.37<br>[4.64, 6.2] | 1.96<br>[1.64, 2.34] | 2.94<br>[2.59, 3.38] | 0.45<br>[0.38, 0.53] |
| Cooke | 17.98<br>[15.46, 21.21] | 6.15<br>[5.17, 7.55] | 10.39<br>[8.98, 12.26] | 1.4<br>[1.21, 1.67] | 3.08<br>[2.63, 3.7] | 1.57<br>[1.32, 1.99] | 1.28<br>[1.1, 1.55] | 0.21<br>[0.18, 0.25] |
| Coryell | 16.4<br>[13.88, 19.75] | 6.03<br>[4.87, 7.51] | 9.04<br>[7.78, 10.65] | 1.3<br>[1.1, 1.56] | 2.88<br>[2.43, 3.55] | 1.56<br>[1.29, 1.98] | 1.14<br>[0.97, 1.34] | 0.19<br>[0.16, 0.24] |
| Cottle | 20.97<br>[18.32, 24.37] | 4.22<br>[3.58, 5] | 15.04<br>[13.15, 17.25] | 1.71<br>[1.46, 2.02] | 3.18<br>[2.76, 3.7] | 1.1<br>[0.92, 1.33] | 1.83<br>[1.59, 2.11] | 0.25<br>[0.21, 0.3] |
| Crockett | 9.54<br>[8.3, 11.06] | 2.94<br>[2.45, 3.58] | 6.13<br>[5.35, 7.03] | 0.45<br>[0.39, 0.52] | 1.67<br>[1.43, 1.97] | 0.78<br>[0.65, 0.95] | 0.82<br>[0.72, 0.95] | 0.07<br>[0.06, 0.08] |
| Crosby | 15.36<br>[13.23, 18.17] | 4.09<br>[3.43, 5] | 10.46<br>[9.07, 12.31] | 0.81<br>[0.69, 0.98] | 2.48<br>[2.09, 2.96] | 1.06<br>[0.86, 1.3] | 1.3<br>[1.11, 1.54] | 0.12<br>[0.1, 0.15] |
| Culberson | 21.39<br>[18.59, 24.78] | 5.5<br>[4.67, 6.53] | 14.84<br>[12.87, 17.11] | 1.06<br>[0.91, 1.24] | 3.34<br>[2.85, 3.96] | 1.41<br>[1.19, 1.72] | 1.78<br>[1.52, 2.08] | 0.16<br>[0.13, 0.18] |
| Dallam | 13.89<br>[11.74, 16.01] | 6.22<br>[5.08, 7.39] | 7<br>[5.98, 8.05] | 0.63<br>[0.53, 0.74] | 2.64<br>[2.2, 3.1] | 1.63<br>[1.3, 1.99] | 0.91<br>[0.78, 1.04] | 0.1<br>[0.08, 0.11] |
| Dallas | 14.88<br>[12.89, 17.38] | 4.87<br>[4.13, 5.88] | 9.11<br>[8, 10.6] | 0.9<br>[0.79, 1.07] | 2.54<br>[2.18, 3.03] | 1.26<br>[1.07, 1.55] | 1.14<br>[0.99, 1.33] | 0.13<br>[0.12, 0.16] |
| Dawson | 24.37<br>[20.08, 28.13] | 9.91<br>[7.98, 11.77] | 12.39<br>[10.39, 14.41] | 2.05<br>[1.68, 2.39] | 4.31<br>[3.59, 5.09] | 2.52<br>[2.05, 3.06] | 1.49<br>[1.27, 1.76] | 0.3<br>[0.25, 0.36] |
| DeWitt | 10.74<br>[8.92, 12.35] | 3.36<br>[2.71, 3.97] | 6.63<br>[5.7, 7.63] | 0.71<br>[0.59, 0.82] | 1.88<br>[1.54, 2.19] | 0.89<br>[0.72, 1.06] | 0.88<br>[0.74, 1] | 0.11<br>[0.09, 0.13] |
| Deaf Smith | 10.19<br>[8.81, 12.04] | 3.25<br>[2.75, 3.95] | 6.57<br>[5.76, 7.7] | 0.33<br>[0.29, 0.39] | 1.79<br>[1.5, 2.09] | 0.86<br>[0.71, 1.05] | 0.87<br>[0.75, 1.01] | 0.05<br>[0.04, 0.06] |
| Delta | 9.52<br>[8.34, 11.22] | 3.54<br>[3.01, 4.39] | 5.29<br>[4.67, 6.07] | 0.67<br>[0.59, 0.79] | 1.77<br>[1.54, 2.06] | 0.94<br>[0.79, 1.14] | 0.73<br>[0.63, 0.83] | 0.1<br>[0.09, 0.12] |
| Denton | 31.32<br>[25.64, 37.04] | 11.23<br>[8.7, 13.7] | 16.94<br>[14.17, 19.73] | 3.11<br>[2.56, 3.65] | 5.22<br>[4.23, 6.27] | 2.85<br>[2.17, 3.47] | 1.96<br>[1.66, 2.32] | 0.44<br>[0.36, 0.53] |
| Dickens | 22.94<br>[19.77, 26.2] | 6.31<br>[5.26, 7.33] | 14.73<br>[12.83, 16.8] | 1.84<br>[1.57, 2.13] | 3.7<br>[3.15, 4.34] | 1.64<br>[1.37, 1.96] | 1.78<br>[1.52, 2.09] | 0.27<br>[0.23, 0.32] |
| Dimmit | 12.23<br>[10.58, 14.03] | 3.22<br>[2.73, 3.89] | 8.61<br>[7.51, 9.84] | 0.41<br>[0.35, 0.47] | 2<br>[1.69, 2.34] | 0.84<br>[0.7, 1.02] | 1.09<br>[0.93, 1.27] | 0.06<br>[0.05, 0.07] |
| Donley | 70.79<br>[59.11, 82.27] | 11.8<br>[9.69, 13.98] | 54.8<br>[46.21, 63.86] | 4.08<br>[3.32, 4.79] | 9.5<br>[7.78, 11.28] | 2.94<br>[2.35, 3.57] | 5.96<br>[4.97, 7.12] | 0.57<br>[0.46, 0.69] |
| Duval | 13.82<br>[11.79, 15.79] | 3.98<br>[3.29, 4.76] | 9.2<br>[7.9, 10.37] | 0.63<br>[0.53, 0.71] | 2.27<br>[1.93, 2.69] | 1.03<br>[0.84, 1.25] | 1.15<br>[0.99, 1.34] | 0.09<br>[0.08, 0.11] |
| Eastland | 42.79<br>[36.59, 50.93] | 11.84<br>[9.91, 14.29] | 28.23<br>[23.8, 33.43] | 2.98<br>[2.54, 3.54] | 6.51<br>[5.48, 7.89] | 2.97<br>[2.43, 3.62] | 3.15<br>[2.67, 3.78] | 0.42<br>[0.35, 0.51] |
| Ector | 12.7<br>[11.08, 14.65] | 4.67<br>[3.94, 5.62] | 7.42<br>[6.46, 8.54] | 0.61<br>[0.53, 0.71] | 2.27<br>[1.96, 2.69] | 1.22<br>[1.01, 1.47] | 0.96<br>[0.84, 1.12] | 0.09<br>[0.08, 0.11] |
| Edwards | 13.55<br>[11.91, 15.34] | 4.63<br>[3.91, 5.36] | 7.83<br>[7.01, 8.84] | 1.11<br>[0.98, 1.28] | 2.42<br>[2.07, 2.81] | 1.22<br>[1.03, 1.45] | 1.02<br>[0.87, 1.16] | 0.17<br>[0.14, 0.2] |
| El Paso | 13.86<br>[11.47, 16.03] | 4.02<br>[3.3, 4.83] | 9.11<br>[7.61, 10.68] | 0.73<br>[0.6, 0.86] | 2.31<br>[1.88, 2.71] | 1.04<br>[0.83, 1.26] | 1.16<br>[0.95, 1.36] | 0.11<br>[0.09, 0.13] |
| Ellis | 17.63<br>[14.5, 21.18] | 5.97<br>[4.59, 7.53] | 10.43<br>[8.73, 12.35] | 1.23<br>[1.02, 1.47] | 3.02<br>[2.45, 3.65] | 1.53<br>[1.19, 1.94] | 1.29<br>[1.08, 1.51] | 0.18<br>[0.15, 0.22] |
| Erath | 112<br>[92.83, 148.24] | 18.93<br>[15.12, 25.73] | 87.96<br>[72.41, 116.05] | 5.19<br>[4.25, 6.93] | 14.23<br>[11.61, 19.14] | 4.51<br>[3.59, 6.08] | 9.05<br>[7.39, 11.85] | 0.7<br>[0.57, 0.96] |
| Falls | 11.73<br>[10.3, 13.65] | 3.97<br>[3.31, 4.75] | 6.88<br>[6.08, 8.01] | 0.89<br>[0.78, 1.06] | 2.09<br>[1.78, 2.5] | 1.04<br>[0.87, 1.26] | 0.9<br>[0.79, 1.07] | 0.14<br>[0.12, 0.16] |
| Fannin | 20.1<br>[17.29, 24.33] | 6.41<br>[5.17, 7.97] | 11.91<br>[10.22, 14.09] | 1.85<br>[1.59, 2.24] | 3.33<br>[2.79, 4.03] | 1.62<br>[1.31, 2.07] | 1.44<br>[1.24, 1.74] | 0.27<br>[0.22, 0.33] |
| Fayette | 11.93<br>[10.1, 13.96] | 3.56<br>[2.9, 4.33] | 7.46<br>[6.41, 8.77] | 0.86<br>[0.73, 1] | 2.04<br>[1.71, 2.4] | 0.94<br>[0.77, 1.14] | 0.97<br>[0.84, 1.15] | 0.13<br>[0.11, 0.15] |
| Fisher | 25.56<br>[21.56, 30] | 4.75<br>[3.86, 5.69] | 19.33<br>[16.38, 22.62] | 1.47<br>[1.23, 1.75] | 3.68<br>[3.03, 4.35] | 1.2<br>[0.96, 1.46] | 2.26<br>[1.89, 2.72] | 0.22<br>[0.17, 0.25] |
| Floyd | 12.19<br>[10.25, 14.27] | 3.4<br>[2.76, 4.05] | 8.14<br>[7.04, 9.43] | 0.68<br>[0.58, 0.8] | 2.03<br>[1.71, 2.4] | 0.89<br>[0.72, 1.07] | 1.05<br>[0.88, 1.22] | 0.1<br>[0.09, 0.12] |
| Foard | 14.71<br>[13.09, 16.97] | 4.14<br>[3.61, 4.83] | 9.07<br>[8.21, 10.48] | 1.48<br>[1.31, 1.75] | 2.47<br>[2.15, 2.87] | 1.09<br>[0.92, 1.29] | 1.16<br>[1.02, 1.37] | 0.22<br>[0.19, 0.27] |
| Fort Bend | 10.22<br>[8.83, 11.98] | 2.9<br>[2.45, 3.56] | 6.73<br>[5.95, 7.8] | 0.56<br>[0.49, 0.66] | 1.74<br>[1.49, 2.05] | 0.76<br>[0.63, 0.92] | 0.89<br>[0.78, 1.03] | 0.09<br>[0.07, 0.1] |
| Franklin | 14.85<br>[12.41, 17.2] | 4.46<br>[3.56, 5.28] | 9.32<br>[7.97, 10.67] | 1.06<br>[0.89, 1.23] | 2.47<br>[2.04, 2.9] | 1.15<br>[0.91, 1.38] | 1.17<br>[0.99, 1.36] | 0.16<br>[0.13, 0.19] |
| Freestone | 16.28<br>[13.5, 19.32] | 5.73<br>[4.63, 7.18] | 9.06<br>[7.63, 10.46] | 1.47<br>[1.22, 1.73] | 2.82<br>[2.29, 3.38] | 1.48<br>[1.18, 1.83] | 1.13<br>[0.94, 1.33] | 0.22<br>[0.18, 0.26] |
| Frio | 13.58<br>[11.8, 16.23] | 3.52<br>[2.93, 4.34] | 9.54<br>[8.29, 11.27] | 0.57<br>[0.49, 0.68] | 2.19<br>[1.9, 2.63] | 0.91<br>[0.77, 1.14] | 1.19<br>[1.04, 1.42] | 0.09<br>[0.07, 0.1] |
| Gaines | 163.92<br>[126.07, 212.62] | 87.22<br>[65.81, 116.74] | 62.37<br>[49.91, 78.12] | 14.57<br>[11.49, 18.57] | 29.47<br>[22.62, 38.47] | 20.93<br>[15.73, 28.04] | 6.41<br>[5.08, 8.34] | 1.93<br>[1.51, 2.56] |
| Galveston | 15.82<br>[13.46, 18.84] | 5<br>[4.09, 6.13] | 9.61<br>[8.21, 11.42] | 1.17<br>[0.99, 1.41] | 2.68<br>[2.23, 3.23] | 1.3<br>[1.05, 1.62] | 1.21<br>[1.03, 1.44] | 0.17<br>[0.14, 0.21] |
| Garza | 15.11<br>[13, 17.38] | 4.32<br>[3.57, 5.06] | 9.58<br>[8.22, 10.98] | 1.23<br>[1.05, 1.42] | 2.51<br>[2.13, 2.94] | 1.13<br>[0.93, 1.34] | 1.2<br>[1.05, 1.4] | 0.18<br>[0.16, 0.21] |
| Gillespie | 25.2<br>[21.12, 29.89] | 8.71<br>[7.09, 10.7] | 13.32<br>[11.43, 15.82] | 3.01<br>[2.54, 3.59] | 4.28<br>[3.44, 5.17] | 2.2<br>[1.71, 2.77] | 1.59<br>[1.35, 1.91] | 0.44<br>[0.36, 0.54] |
| Glasscock | 16.18<br>[14.34, 18.61] | 5.88<br>[5.02, 6.78] | 8.96<br>[8.05, 10.27] | 1.39<br>[1.22, 1.62] | 2.9<br>[2.52, 3.37] | 1.55<br>[1.31, 1.81] | 1.15<br>[1.01, 1.33] | 0.21<br>[0.18, 0.25] |
| Goliad | 13.48<br>[11.69, 15.79] | 4.05<br>[3.43, 4.92] | 8.15<br>[7.13, 9.39] | 1.27<br>[1.09, 1.47] | 2.31<br>[1.96, 2.69] | 1.06<br>[0.87, 1.29] | 1.04<br>[0.91, 1.19] | 0.19<br>[0.16, 0.22] |
| Gonzales | 13.34<br>[11.48, 15.92] | 4.27<br>[3.59, 5.25] | 8.38<br>[7.29, 9.84] | 0.67<br>[0.58, 0.8] | 2.28<br>[1.92, 2.75] | 1.12<br>[0.92, 1.37] | 1.06<br>[0.91, 1.26] | 0.1<br>[0.08, 0.12] |
| Gray | 13.41<br>[11.56, 15.71] | 4.18<br>[3.53, 5.14] | 8.34<br>[7.15, 9.75] | 0.9<br>[0.76, 1.06] | 2.29<br>[1.94, 2.69] | 1.09<br>[0.92, 1.35] | 1.06<br>[0.89, 1.25] | 0.13<br>[0.11, 0.16] |
| Grayson | 18.26<br>[15.57, 21.86] | 6.28<br>[5.18, 7.93] | 10.48<br>[8.92, 12.29] | 1.4<br>[1.17, 1.67] | 3.12<br>[2.66, 3.78] | 1.62<br>[1.33, 2.08] | 1.3<br>[1.08, 1.51] | 0.21<br>[0.17, 0.25] |
| Gregg | 16.73<br>[14.45, 20] | 5.44<br>[4.54, 6.58] | 10.28<br>[9.05, 12.3] | 1.02<br>[0.89, 1.23] | 2.84<br>[2.37, 3.37] | 1.41<br>[1.14, 1.75] | 1.27<br>[1.11, 1.5] | 0.15<br>[0.13, 0.18] |
| Grimes | 15.21<br>[12.95, 17.66] | 5.34<br>[4.38, 6.37] | 8.72<br>[7.46, 10] | 1.17<br>[0.99, 1.35] | 2.65<br>[2.22, 3.16] | 1.38<br>[1.13, 1.68] | 1.11<br>[0.91, 1.29] | 0.17<br>[0.14, 0.21] |
| Guadalupe | 14.05<br>[11.97, 16.58] | 4.09<br>[3.43, 5.12] | 9.04<br>[7.72, 10.52] | 0.93<br>[0.79, 1.09] | 2.34<br>[2.01, 2.84] | 1.06<br>[0.88, 1.33] | 1.13<br>[0.98, 1.32] | 0.14<br>[0.12, 0.17] |
| Hale | 14.35<br>[12.32, 16.94] | 3.79<br>[3.16, 4.68] | 9.91<br>[8.49, 11.78] | 0.65<br>[0.55, 0.77] | 2.32<br>[1.98, 2.79] | 0.99<br>[0.82, 1.23] | 1.24<br>[1.06, 1.46] | 0.1<br>[0.08, 0.12] |
| Hall | 27.41<br>[23.39, 31.78] | 7.83<br>[6.52, 9.28] | 16.61<br>[14.16, 19.12] | 3.01<br>[2.55, 3.51] | 4.41<br>[3.77, 5.14] | 2.01<br>[1.69, 2.43] | 1.95<br>[1.68, 2.26] | 0.44<br>[0.37, 0.52] |
| Hamilton | 10.42<br>[9.23, 12.1] | 3.26<br>[2.8, 3.96] | 6.27<br>[5.54, 7.09] | 0.86<br>[0.76, 1] | 1.84<br>[1.58, 2.18] | 0.86<br>[0.71, 1.07] | 0.84<br>[0.74, 0.98] | 0.13<br>[0.11, 0.15] |
| Hansford | 15.66<br>[13.38, 18.02] | 4.49<br>[3.74, 5.32] | 10.45<br>[9.02, 11.98] | 0.76<br>[0.65, 0.88] | 2.58<br>[2.17, 2.99] | 1.18<br>[0.94, 1.41] | 1.29<br>[1.12, 1.49] | 0.11<br>[0.09, 0.13] |
| Hardeman | 16.43<br>[14.1, 18.95] | 4.73<br>[3.9, 5.63] | 10.72<br>[9.16, 12.31] | 1.04<br>[0.88, 1.19] | 2.71<br>[2.27, 3.17] | 1.22<br>[1, 1.48] | 1.33<br>[1.13, 1.52] | 0.15<br>[0.13, 0.18] |
| Hardin | 14.98<br>[12.38, 17.59] | 5.42<br>[4.3, 6.48] | 8.3<br>[7.11, 9.66] | 1.21<br>[1, 1.43] | 2.65<br>[2.16, 3.15] | 1.4<br>[1.12, 1.69] | 1.06<br>[0.89, 1.25] | 0.18<br>[0.15, 0.21] |
| Harris | 13.88<br>[11.54, 16.38] | 4.51<br>[3.59, 5.48] | 8.56<br>[7.21, 9.97] | 0.81<br>[0.67, 0.96] | 2.37<br>[1.95, 2.87] | 1.17<br>[0.92, 1.46] | 1.08<br>[0.91, 1.26] | 0.12<br>[0.1, 0.15] |
| Harrison | 18.21<br>[15.44, 21.25] | 5.06<br>[4.09, 6.11] | 12.05<br>[10.28, 13.87] | 1.13<br>[0.95, 1.3] | 2.92<br>[2.44, 3.46] | 1.3<br>[1.05, 1.58] | 1.45<br>[1.26, 1.73] | 0.16<br>[0.14, 0.2] |
| Hartley | 20.08<br>[17.34, 23.16] | 7.92<br>[6.68, 9.38] | 10.23<br>[8.9, 11.51] | 1.94<br>[1.65, 2.22] | 3.62<br>[3.07, 4.19] | 2.07<br>[1.71, 2.45] | 1.28<br>[1.07, 1.46] | 0.29<br>[0.24, 0.34] |
| Haskell | 17.04<br>[14.7, 20.34] | 5.23<br>[4.36, 6.42] | 10.1<br>[8.77, 11.85] | 1.73<br>[1.47, 2.07] | 2.88<br>[2.46, 3.52] | 1.34<br>[1.12, 1.7] | 1.26<br>[1.1, 1.47] | 0.25<br>[0.21, 0.31] |
| Hays | 34.14<br>[28.37, 42.66] | 9.55<br>[8.04, 12.69] | 22.14<br>[18.21, 27.35] | 2.45<br>[2.01, 3.1] | 5.26<br>[4.38, 6.72] | 2.41<br>[1.95, 3.13] | 2.51<br>[2.08, 3.16] | 0.35<br>[0.29, 0.45] |
| Hemphill | 18.12<br>[15.08, 20.94] | 4.59<br>[3.75, 5.53] | 12.39<br>[10.4, 14.26] | 1.12<br>[0.93, 1.3] | 2.88<br>[2.35, 3.35] | 1.2<br>[0.97, 1.45] | 1.51<br>[1.25, 1.76] | 0.17<br>[0.13, 0.19] |
| Henderson | 18.18<br>[15.2, 21.62] | 6.27<br>[5.05, 7.74] | 10.36<br>[8.85, 12.14] | 1.58<br>[1.33, 1.86] | 3.13<br>[2.61, 3.79] | 1.61<br>[1.27, 2] | 1.28<br>[1.08, 1.53] | 0.23<br>[0.19, 0.28] |
| Hidalgo | 11.38<br>[9.71, 12.93] | 3.28<br>[2.65, 3.82] | 7.69<br>[6.64, 8.71] | 0.43<br>[0.37, 0.49] | 1.93<br>[1.62, 2.21] | 0.86<br>[0.7, 1] | 0.99<br>[0.85, 1.13] | 0.06<br>[0.05, 0.07] |
| Hill | 16.47<br>[14, 19.41] | 5.5<br>[4.45, 6.75] | 9.67<br>[8.26, 11.29] | 1.31<br>[1.11, 1.57] | 2.82<br>[2.35, 3.36] | 1.43<br>[1.13, 1.76] | 1.21<br>[1.03, 1.41] | 0.2<br>[0.16, 0.24] |
| Hockley | 16.92<br>[14.65, 20.53] | 5.16<br>[4.25, 6.51] | 10.8<br>[9.42, 12.75] | 0.93<br>[0.8, 1.13] | 2.8<br>[2.39, 3.52] | 1.33<br>[1.08, 1.7] | 1.33<br>[1.15, 1.61] | 0.14<br>[0.11, 0.17] |
| Hood | 21<br>[18.06, 25.76] | 7.48<br>[6.25, 9.5] | 11.18<br>[9.53, 13.2] | 2.37<br>[2.01, 2.86] | 3.6<br>[3, 4.5] | 1.89<br>[1.56, 2.45] | 1.37<br>[1.17, 1.62] | 0.35<br>[0.29, 0.42] |
| Hopkins | 13.43<br>[11.29, 15.57] | 4.44<br>[3.64, 5.28] | 8.07<br>[6.87, 9.32] | 0.9<br>[0.75, 1.04] | 2.33<br>[1.9, 2.77] | 1.16<br>[0.92, 1.41] | 1.04<br>[0.87, 1.23] | 0.13<br>[0.11, 0.16] |
| Houston | 16.6<br>[13.84, 19.76] | 5.96<br>[4.73, 7.3] | 9.05<br>[7.77, 10.58] | 1.59<br>[1.33, 1.89] | 2.92<br>[2.41, 3.49] | 1.55<br>[1.23, 1.91] | 1.14<br>[0.98, 1.34] | 0.24<br>[0.2, 0.28] |
| Howard | 15.76<br>[13.6, 18.34] | 4.6<br>[3.85, 5.44] | 10.16<br>[8.81, 11.81] | 0.98<br>[0.85, 1.15] | 2.62<br>[2.21, 3.06] | 1.2<br>[1, 1.41] | 1.26<br>[1.09, 1.47] | 0.15<br>[0.13, 0.17] |
| Hudspeth | 42.67<br>[37.03, 51.6] | 7.71<br>[6.48, 9.27] | 30.96<br>[26.85, 36.87] | 4.12<br>[3.52, 5.07] | 6.07<br>[5.16, 7.33] | 1.96<br>[1.65, 2.4] | 3.52<br>[3.03, 4.21] | 0.59<br>[0.49, 0.74] |
| Hunt | 18.26<br>[15.31, 21.61] | 5.74<br>[4.69, 7.01] | 11.33<br>[9.64, 13.24] | 1.16<br>[0.97, 1.36] | 3.03<br>[2.55, 3.57] | 1.47<br>[1.2, 1.79] | 1.39<br>[1.17, 1.62] | 0.17<br>[0.14, 0.2] |
| Hutchinson | 17.08<br>[14.63, 19.58] | 5.19<br>[4.33, 6.11] | 10.69<br>[9.25, 12.25] | 1.23<br>[1.05, 1.4] | 2.83<br>[2.42, 3.24] | 1.35<br>[1.11, 1.59] | 1.31<br>[1.13, 1.49] | 0.18<br>[0.15, 0.21] |
| Irion | 12.07<br>[10.33, 13.87] | 4.31<br>[3.6, 5.1] | 6.62<br>[5.78, 7.56] | 1.13<br>[0.96, 1.32] | 2.2<br>[1.86, 2.56] | 1.15<br>[0.94, 1.38] | 0.88<br>[0.77, 1] | 0.17<br>[0.14, 0.2] |
| Jack | 14.02<br>[11.91, 16.31] | 4.2<br>[3.45, 5.09] | 8.66<br>[7.45, 10.1] | 1.14<br>[0.96, 1.35] | 2.35<br>[1.97, 2.83] | 1.09<br>[0.89, 1.33] | 1.1<br>[0.93, 1.29] | 0.17<br>[0.14, 0.21] |
| Jackson | 12.1<br>[10.65, 14.24] | 4.12<br>[3.55, 5] | 7.35<br>[6.56, 8.53] | 0.62<br>[0.55, 0.74] | 2.12<br>[1.84, 2.52] | 1.08<br>[0.91, 1.3] | 0.96<br>[0.84, 1.14] | 0.09<br>[0.08, 0.11] |
| Jasper | 22.24<br>[18.76, 26.54] | 8.15<br>[6.57, 9.94] | 12.05<br>[10.17, 14.01] | 2.12<br>[1.77, 2.49] | 3.87<br>[3.21, 4.57] | 2.11<br>[1.65, 2.57] | 1.46<br>[1.23, 1.7] | 0.31<br>[0.25, 0.36] |
| Jeff Davis | 17.65<br>[16, 19.6] | 4.53<br>[3.98, 5.08] | 5.49<br>[5.01, 6.01] | 7.64<br>[6.82, 8.58] | 3.17<br>[2.76, 3.6] | 1.23<br>[1.06, 1.4] | 0.77<br>[0.68, 0.86] | 1.17<br>[1, 1.37] |
| Jefferson | 12.25<br>[10.34, 14.2] | 4.1<br>[3.38, 4.9] | 7.36<br>[6.3, 8.44] | 0.78<br>[0.66, 0.91] | 2.14<br>[1.76, 2.52] | 1.08<br>[0.86, 1.31] | 0.96<br>[0.8, 1.09] | 0.12<br>[0.1, 0.14] |
| Jim Hogg | 16.24<br>[14.04, 18.46] | 4.47<br>[3.69, 5.29] | 10.93<br>[9.67, 12.27] | 0.86<br>[0.75, 0.97] | 2.63<br>[2.24, 3.07] | 1.15<br>[0.92, 1.41] | 1.35<br>[1.17, 1.55] | 0.13<br>[0.11, 0.15] |
| Jim Wells | 11.23<br>[9.61, 12.89] | 3.32<br>[2.75, 3.87] | 7.32<br>[6.31, 8.47] | 0.56<br>[0.48, 0.65] | 1.91<br>[1.63, 2.21] | 0.87<br>[0.73, 1.03] | 0.95<br>[0.83, 1.12] | 0.09<br>[0.07, 0.1] |
| Johnson | 18.69<br>[15.98, 22.27] | 6.6<br>[5.48, 8.19] | 10.71<br>[9.25, 12.56] | 1.45<br>[1.23, 1.71] | 3.22<br>[2.71, 3.82] | 1.69<br>[1.4, 2.11] | 1.32<br>[1.12, 1.57] | 0.21<br>[0.18, 0.26] |
| Jones | 15.16<br>[12.95, 17.9] | 3.82<br>[3.11, 4.75] | 10.05<br>[8.76, 11.89] | 1.31<br>[1.11, 1.56] | 2.45<br>[2.05, 2.97] | 1<br>[0.79, 1.25] | 1.25<br>[1.08, 1.51] | 0.19<br>[0.16, 0.24] |
| Karnes | 14.65<br>[12.57, 17.01] | 3.97<br>[3.27, 4.83] | 9.73<br>[8.39, 11.06] | 0.98<br>[0.84, 1.13] | 2.39<br>[2.02, 2.83] | 1.03<br>[0.84, 1.27] | 1.21<br>[1.03, 1.42] | 0.15<br>[0.12, 0.17] |
| Kaufman | 13.46<br>[11.21, 15.84] | 4.83<br>[3.93, 5.79] | 7.85<br>[6.6, 9.15] | 0.8<br>[0.67, 0.95] | 2.4<br>[1.98, 2.83] | 1.26<br>[1.01, 1.52] | 1.02<br>[0.86, 1.18] | 0.12<br>[0.1, 0.15] |
| Kendall | 25.67<br>[21.49, 30.81] | 8.09<br>[6.51, 10.15] | 14.78<br>[12.77, 17.71] | 2.73<br>[2.3, 3.33] | 4.2<br>[3.47, 5.1] | 2.06<br>[1.65, 2.58] | 1.75<br>[1.46, 2.1] | 0.4<br>[0.33, 0.49] |
| Kenedy | 10.74<br>[9.74, 11.76] | 1.82<br>[1.58, 2.09] | 7.87<br>[7.24, 8.51] | 1.03<br>[0.91, 1.18] | 1.7<br>[1.52, 1.91] | 0.49<br>[0.41, 0.57] | 1.05<br>[0.95, 1.17] | 0.16<br>[0.14, 0.19] |
| Kent | 41.62<br>[37.42, 45.63] | 10.17<br>[8.99, 11.43] | 26.62<br>[24.36, 28.9] | 4.87<br>[4.28, 5.54] | 6.61<br>[5.83, 7.41] | 2.7<br>[2.35, 3.04] | 3.18<br>[2.83, 3.58] | 0.72<br>[0.6, 0.84] |
| Kerr | 28.4<br>[23.42, 34.33] | 8.6<br>[6.91, 10.47] | 16.77<br>[13.98, 20.26] | 2.99<br>[2.48, 3.61] | 4.57<br>[3.68, 5.53] | 2.16<br>[1.7, 2.63] | 1.96<br>[1.63, 2.35] | 0.43<br>[0.35, 0.53] |
| Kimble | 13.84<br>[11.92, 16.66] | 4.49<br>[3.78, 5.55] | 8.15<br>[7.08, 9.7] | 1.23<br>[1.06, 1.5] | 2.4<br>[2.08, 2.92] | 1.18<br>[0.99, 1.48] | 1.04<br>[0.91, 1.25] | 0.18<br>[0.16, 0.22] |
| King | 8.52<br>[7.87, 9.28] | 2.63<br>[2.37, 2.89] | 4.22<br>[3.94, 4.56] | 1.66<br>[1.5, 1.85] | 1.61<br>[1.44, 1.8] | 0.73<br>[0.65, 0.82] | 0.62<br>[0.56, 0.69] | 0.26<br>[0.22, 0.3] |
| Kinney | 23.5<br>[20.41, 27.16] | 5.22<br>[4.46, 6.19] | 16.48<br>[14.3, 18.96] | 1.77<br>[1.52, 2.07] | 3.56<br>[3.07, 4.23] | 1.36<br>[1.12, 1.66] | 1.95<br>[1.68, 2.24] | 0.26<br>[0.21, 0.31] |
| Kleberg | 36.2<br>[29.54, 43.37] | 7.75<br>[6.3, 9.65] | 26.87<br>[21.99, 32.08] | 1.45<br>[1.17, 1.74] | 5.16<br>[4.21, 6.31] | 1.93<br>[1.55, 2.47] | 3.02<br>[2.44, 3.66] | 0.21<br>[0.17, 0.25] |
| Knox | 8.04<br>[6.83, 9.18] | 2.54<br>[2.13, 2.95] | 5.04<br>[4.32, 5.67] | 0.44<br>[0.37, 0.5] | 1.45<br>[1.21, 1.69] | 0.68<br>[0.56, 0.81] | 0.7<br>[0.59, 0.8] | 0.07<br>[0.06, 0.08] |
| La Salle | 34.13<br>[29.46, 40.92] | 6.83<br>[5.73, 8.4] | 25.74<br>[22.07, 30.46] | 1.62<br>[1.39, 1.95] | 4.87<br>[4.13, 5.88] | 1.72<br>[1.41, 2.14] | 2.93<br>[2.48, 3.5] | 0.23<br>[0.19, 0.28] |
| Lamar | 12.98<br>[11.01, 15.77] | 4.48<br>[3.66, 5.45] | 7.67<br>[6.57, 9.03] | 0.83<br>[0.7, 1.01] | 2.29<br>[1.85, 2.76] | 1.17<br>[0.94, 1.48] | 0.98<br>[0.82, 1.18] | 0.12<br>[0.1, 0.16] |
| Lamb | 11.05<br>[9.19, 12.79] | 3.05<br>[2.49, 3.63] | 7.48<br>[6.31, 8.58] | 0.52<br>[0.43, 0.6] | 1.85<br>[1.52, 2.17] | 0.81<br>[0.64, 0.98] | 0.96<br>[0.82, 1.13] | 0.08<br>[0.06, 0.09] |
| Lampasas | 22.03<br>[18.61, 25.98] | 7.35<br>[5.91, 8.67] | 12.41<br>[10.62, 14.53] | 2.36<br>[2, 2.8] | 3.71<br>[3.1, 4.46] | 1.86<br>[1.48, 2.28] | 1.49<br>[1.29, 1.77] | 0.34<br>[0.29, 0.42] |
| Lavaca | 16.11<br>[13.63, 18.85] | 5.34<br>[4.34, 6.4] | 9.61<br>[8.24, 11.07] | 1.17<br>[0.99, 1.36] | 2.76<br>[2.36, 3.29] | 1.39<br>[1.14, 1.69] | 1.21<br>[1.04, 1.41] | 0.17<br>[0.15, 0.21] |
| Lee | 19.7<br>[16.56, 23.67] | 5.85<br>[4.67, 7.23] | 12.46<br>[10.55, 14.73] | 1.37<br>[1.15, 1.65] | 3.24<br>[2.65, 3.9] | 1.51<br>[1.15, 1.86] | 1.51<br>[1.29, 1.83] | 0.2<br>[0.17, 0.24] |
| Leon | 12.01<br>[10.52, 13.85] | 3.95<br>[3.39, 4.61] | 7.17<br>[6.3, 8.12] | 0.88<br>[0.77, 1.01] | 2.11<br>[1.8, 2.43] | 1.04<br>[0.89, 1.25] | 0.93<br>[0.8, 1.07] | 0.13<br>[0.11, 0.15] |
| Liberty | 9.83<br>[8.38, 11.4] | 3.26<br>[2.69, 3.84] | 6.12<br>[5.27, 6.92] | 0.47<br>[0.39, 0.54] | 1.76<br>[1.48, 2.08] | 0.87<br>[0.71, 1.03] | 0.82<br>[0.7, 0.93] | 0.07<br>[0.06, 0.08] |
| Limestone | 11.78<br>[10.17, 13.65] | 3.69<br>[3.09, 4.38] | 7.3<br>[6.37, 8.44] | 0.79<br>[0.68, 0.91] | 2.03<br>[1.74, 2.41] | 0.97<br>[0.8, 1.18] | 0.95<br>[0.82, 1.12] | 0.12<br>[0.1, 0.14] |
| Lipscomb | 17.14<br>[14.82, 20] | 4.99<br>[4.12, 6.06] | 10.93<br>[9.58, 12.79] | 1.21<br>[1.04, 1.43] | 2.82<br>[2.37, 3.39] | 1.29<br>[1.05, 1.59] | 1.35<br>[1.18, 1.62] | 0.18<br>[0.15, 0.21] |
| Live Oak | 10.8<br>[9.03, 12.2] | 3.29<br>[2.72, 3.89] | 6.59<br>[5.57, 7.46] | 0.9<br>[0.75, 1.02] | 1.89<br>[1.59, 2.16] | 0.87<br>[0.72, 1.04] | 0.87<br>[0.74, 1] | 0.14<br>[0.11, 0.16] |
| Llano | 20.62<br>[17.55, 24.54] | 7.34<br>[6.13, 9.06] | 10.58<br>[9.04, 12.49] | 2.7<br>[2.29, 3.22] | 3.58<br>[3.04, 4.38] | 1.88<br>[1.54, 2.36] | 1.29<br>[1.13, 1.59] | 0.4<br>[0.34, 0.49] |
| Lubbock | 34.68<br>[28.72, 42.21] | 9.01<br>[7.2, 11.11] | 23.66<br>[19.76, 28.64] | 1.97<br>[1.63, 2.42] | 5.2<br>[4.29, 6.44] | 2.25<br>[1.8, 2.85] | 2.68<br>[2.22, 3.24] | 0.28<br>[0.23, 0.35] |
| Lynn | 13.8<br>[11.82, 16.25] | 4.89<br>[4.04, 5.98] | 7.88<br>[6.83, 9.11] | 1.03<br>[0.88, 1.2] | 2.46<br>[2.06, 2.93] | 1.28<br>[1.06, 1.59] | 1.01<br>[0.88, 1.18] | 0.15<br>[0.13, 0.18] |
| Madison | 14.87<br>[12.53, 16.94] | 4.73<br>[4, 5.59] | 9.28<br>[7.88, 10.49] | 0.81<br>[0.67, 0.92] | 2.51<br>[2.12, 2.93] | 1.23<br>[1.02, 1.46] | 1.17<br>[0.99, 1.34] | 0.12<br>[0.1, 0.14] |
| Marion | 11.51<br>[9.88, 13.44] | 3.14<br>[2.57, 3.92] | 7.43<br>[6.42, 8.45] | 0.94<br>[0.8, 1.09] | 1.92<br>[1.65, 2.32] | 0.82<br>[0.67, 1.05] | 0.96<br>[0.83, 1.12] | 0.14<br>[0.12, 0.17] |
| Martin | 13.19<br>[11.01, 15.77] | 4.33<br>[3.44, 5.3] | 8<br>[6.84, 9.34] | 0.85<br>[0.71, 1.01] | 2.27<br>[1.86, 2.73] | 1.13<br>[0.89, 1.39] | 1.02<br>[0.86, 1.22] | 0.13<br>[0.1, 0.16] |
| Mason | 24.56<br>[21.43, 28.66] | 6.65<br>[5.63, 8.02] | 14.93<br>[13.13, 17.39] | 2.91<br>[2.51, 3.45] | 3.93<br>[3.33, 4.68] | 1.71<br>[1.43, 2.1] | 1.78<br>[1.54, 2.11] | 0.43<br>[0.37, 0.52] |
| Matagorda | 11.82<br>[9.89, 13.77] | 4.2<br>[3.44, 5.02] | 7.01<br>[5.86, 8.13] | 0.67<br>[0.56, 0.78] | 2.12<br>[1.76, 2.49] | 1.1<br>[0.89, 1.33] | 0.92<br>[0.76, 1.06] | 0.1<br>[0.08, 0.12] |
| Maverick | 10.99<br>[9.44, 12.84] | 3.4<br>[2.81, 4.17] | 7.2<br>[6.27, 8.26] | 0.38<br>[0.33, 0.44] | 1.9<br>[1.59, 2.25] | 0.89<br>[0.72, 1.1] | 0.95<br>[0.81, 1.1] | 0.06<br>[0.05, 0.07] |
| McCulloch | 15.33<br>[13.06, 18.05] | 4.83<br>[3.93, 5.92] | 9.07<br>[7.68, 10.44] | 1.48<br>[1.24, 1.73] | 2.61<br>[2.16, 3.13] | 1.25<br>[1.02, 1.54] | 1.14<br>[0.96, 1.34] | 0.22<br>[0.18, 0.26] |
| McLennan | 28.67<br>[24, 34.8] | 7.5<br>[6.07, 9.23] | 19.74<br>[16.76, 23.93] | 1.48<br>[1.25, 1.82] | 4.38<br>[3.58, 5.42] | 1.88<br>[1.51, 2.34] | 2.26<br>[1.9, 2.75] | 0.22<br>[0.17, 0.27] |
| McMullen | 29.11<br>[26.74, 32.02] | 5.64<br>[5, 6.33] | 20.92<br>[19.41, 22.82] | 2.58<br>[2.31, 2.96] | 4.4<br>[3.97, 5.04] | 1.49<br>[1.3, 1.72] | 2.53<br>[2.31, 2.86] | 0.38<br>[0.33, 0.45] |
| Medina | 15.89<br>[13.57, 18.21] | 4.45<br>[3.62, 5.3] | 10.51<br>[8.93, 11.9] | 1<br>[0.85, 1.15] | 2.58<br>[2.2, 3] | 1.14<br>[0.95, 1.38] | 1.3<br>[1.13, 1.49] | 0.15<br>[0.12, 0.17] |
| Menard | 10.64<br>[9.25, 12.2] | 2.81<br>[2.37, 3.37] | 6.99<br>[6.15, 8.01] | 0.84<br>[0.73, 0.98] | 1.8<br>[1.52, 2.09] | 0.74<br>[0.6, 0.9] | 0.92<br>[0.8, 1.06] | 0.13<br>[0.11, 0.15] |
| Midland | 11.15<br>[9.68, 13.35] | 4.38<br>[3.6, 5.29] | 6.14<br>[5.39, 7.24] | 0.61<br>[0.53, 0.73] | 2.05<br>[1.74, 2.48] | 1.15<br>[0.93, 1.43] | 0.82<br>[0.71, 0.97] | 0.09<br>[0.08, 0.11] |
| Milam | 14.95<br>[12.76, 17.83] | 4.61<br>[3.79, 5.63] | 9.27<br>[8.02, 10.92] | 1.05<br>[0.9, 1.27] | 2.51<br>[2.09, 3.01] | 1.2<br>[0.97, 1.47] | 1.16<br>[1, 1.39] | 0.16<br>[0.13, 0.19] |
| Mills | 13.49<br>[11.45, 15.83] | 3.81<br>[3.13, 4.53] | 8.43<br>[7.19, 9.78] | 1.28<br>[1.08, 1.5] | 2.25<br>[1.89, 2.64] | 0.99<br>[0.81, 1.2] | 1.08<br>[0.92, 1.24] | 0.19<br>[0.16, 0.23] |
| Mitchell | 13.4<br>[11.27, 15.7] | 3.41<br>[2.77, 4.18] | 8.95<br>[7.62, 10.41] | 0.99<br>[0.83, 1.18] | 2.18<br>[1.81, 2.6] | 0.88<br>[0.71, 1.13] | 1.14<br>[0.96, 1.34] | 0.15<br>[0.12, 0.18] |
| Montague | 27.81<br>[23.16, 32.77] | 10.43<br>[8.51, 12.54] | 13.97<br>[11.77, 16.39] | 3.26<br>[2.71, 3.87] | 4.81<br>[3.97, 5.8] | 2.64<br>[2.11, 3.29] | 1.66<br>[1.38, 1.96] | 0.47<br>[0.39, 0.57] |
| Montgomery | 16.49<br>[14.25, 19.89] | 5.77<br>[4.77, 7.16] | 9.56<br>[8.26, 11.09] | 1.25<br>[1.08, 1.49] | 2.88<br>[2.42, 3.57] | 1.5<br>[1.2, 1.94] | 1.2<br>[1.02, 1.42] | 0.19<br>[0.16, 0.23] |
| Moore | 10.84<br>[9.28, 12.62] | 3.87<br>[3.21, 4.66] | 6.58<br>[5.69, 7.63] | 0.42<br>[0.36, 0.48] | 1.95<br>[1.66, 2.34] | 1.02<br>[0.85, 1.25] | 0.87<br>[0.75, 1.01] | 0.06<br>[0.05, 0.08] |
| Morris | 10.52<br>[9.05, 12.49] | 2.96<br>[2.54, 3.59] | 6.86<br>[5.98, 7.95] | 0.68<br>[0.59, 0.81] | 1.78<br>[1.52, 2.15] | 0.78<br>[0.65, 0.94] | 0.9<br>[0.78, 1.05] | 0.1<br>[0.09, 0.12] |
| Motley | 29.46<br>[26.5, 33.29] | 8.39<br>[7.54, 9.52] | 17.6<br>[15.83, 19.63] | 3.49<br>[3.06, 4.01] | 4.83<br>[4.27, 5.49] | 2.21<br>[1.97, 2.56] | 2.11<br>[1.85, 2.4] | 0.51<br>[0.44, 0.61] |
| Nacogdoches | 49.43<br>[40.46, 60.49] | 10.55<br>[8.58, 13.29] | 36.75<br>[29.66, 44.67] | 2.13<br>[1.72, 2.61] | 6.87<br>[5.47, 8.43] | 2.59<br>[2.07, 3.29] | 4<br>[3.16, 4.85] | 0.3<br>[0.23, 0.38] |
| Navarro | 14.25<br>[12.55, 16.88] | 4.17<br>[3.5, 5.13] | 9.42<br>[8.21, 10.9] | 0.72<br>[0.62, 0.85] | 2.35<br>[2.06, 2.84] | 1.07<br>[0.9, 1.33] | 1.17<br>[1.03, 1.39] | 0.11<br>[0.09, 0.13] |
| Newton | 21<br>[17.47, 24.48] | 7.35<br>[5.68, 8.66] | 11.15<br>[9.43, 12.88] | 2.46<br>[2.03, 2.87] | 3.59<br>[2.98, 4.26] | 1.88<br>[1.48, 2.26] | 1.37<br>[1.16, 1.59] | 0.36<br>[0.29, 0.43] |
| Nolan | 15.06<br>[12.96, 17.7] | 4.32<br>[3.62, 5.18] | 9.9<br>[8.6, 11.53] | 0.86<br>[0.73, 1] | 2.48<br>[2.12, 2.94] | 1.12<br>[0.93, 1.36] | 1.23<br>[1.06, 1.45] | 0.13<br>[0.11, 0.15] |
| Nueces | 15.24<br>[13.09, 18.03] | 4.5<br>[3.72, 5.39] | 9.85<br>[8.61, 11.67] | 0.88<br>[0.76, 1.05] | 2.53<br>[2.14, 2.98] | 1.16<br>[0.97, 1.42] | 1.23<br>[1.07, 1.45] | 0.13<br>[0.11, 0.16] |
| Ochiltree | 15.81<br>[13.61, 18.25] | 4.7<br>[3.95, 5.71] | 10.33<br>[8.91, 11.87] | 0.71<br>[0.61, 0.83] | 2.62<br>[2.23, 3.07] | 1.23<br>[1.02, 1.49] | 1.29<br>[1.11, 1.51] | 0.11<br>[0.09, 0.13] |
| Oldham | 19.03<br>[16.79, 22.16] | 6.02<br>[5.22, 7.07] | 10.44<br>[9.26, 12.17] | 2.53<br>[2.2, 3.02] | 3.26<br>[2.8, 3.86] | 1.58<br>[1.34, 1.88] | 1.29<br>[1.12, 1.55] | 0.37<br>[0.32, 0.45] |
| Orange | 17.54<br>[15.02, 21.18] | 6.73<br>[5.43, 8.56] | 9.3<br>[8.18, 11.04] | 1.48<br>[1.28, 1.78] | 3.14<br>[2.61, 3.87] | 1.75<br>[1.4, 2.24] | 1.16<br>[1, 1.39] | 0.22<br>[0.19, 0.27] |
| Palo Pinto | 16.25<br>[13.46, 19.14] | 5.72<br>[4.61, 6.85] | 9.15<br>[7.76, 10.69] | 1.32<br>[1.09, 1.56] | 2.83<br>[2.33, 3.42] | 1.49<br>[1.16, 1.81] | 1.16<br>[0.97, 1.35] | 0.2<br>[0.16, 0.24] |
| Panola | 18.26<br>[15.68, 21.46] | 5.66<br>[4.78, 6.87] | 11.19<br>[9.58, 13.09] | 1.31<br>[1.11, 1.55] | 3.03<br>[2.58, 3.59] | 1.46<br>[1.22, 1.77] | 1.37<br>[1.16, 1.59] | 0.19<br>[0.16, 0.23] |
| Parker | 19.79<br>[16.82, 23.09] | 7.31<br>[6.15, 8.76] | 10.64<br>[9.29, 12.26] | 1.84<br>[1.58, 2.15] | 3.45<br>[2.83, 4.04] | 1.86<br>[1.52, 2.23] | 1.3<br>[1.13, 1.51] | 0.27<br>[0.23, 0.32] |
| Parmer | 13.58<br>[11.72, 15.87] | 4.33<br>[3.64, 5.3] | 8.56<br>[7.43, 9.94] | 0.71<br>[0.62, 0.84] | 2.31<br>[1.95, 2.8] | 1.13<br>[0.93, 1.4] | 1.09<br>[0.95, 1.28] | 0.11<br>[0.09, 0.13] |
| Pecos | 16.31<br>[14.02, 19.98] | 5.5<br>[4.6, 6.92] | 9.59<br>[8.29, 11.56] | 1.22<br>[1.04, 1.49] | 2.77<br>[2.38, 3.49] | 1.41<br>[1.18, 1.84] | 1.19<br>[1.03, 1.45] | 0.18<br>[0.15, 0.22] |
| Polk | 10.94<br>[9.56, 12.61] | 3.48<br>[2.95, 4.24] | 6.56<br>[5.77, 7.49] | 0.89<br>[0.77, 1.02] | 1.92<br>[1.67, 2.26] | 0.92<br>[0.77, 1.1] | 0.87<br>[0.76, 1.01] | 0.13<br>[0.12, 0.16] |
| Potter | 13.02<br>[11.06, 15.13] | 4.34<br>[3.53, 5.14] | 7.89<br>[6.76, 9.11] | 0.76<br>[0.64, 0.89] | 2.28<br>[1.91, 2.64] | 1.13<br>[0.9, 1.38] | 1.01<br>[0.86, 1.17] | 0.11<br>[0.1, 0.13] |
| Presidio | 11.66<br>[10.16, 13.9] | 3.52<br>[3.01, 4.3] | 7.5<br>[6.61, 8.86] | 0.59<br>[0.51, 0.7] | 1.98<br>[1.71, 2.42] | 0.92<br>[0.78, 1.14] | 0.97<br>[0.84, 1.17] | 0.09<br>[0.08, 0.11] |
| Rains | 42.62<br>[36.12, 51.85] | 12.6<br>[10.01, 15.66] | 25.34<br>[21.62, 30.54] | 4.59<br>[3.86, 5.64] | 6.64<br>[5.43, 8.08] | 3.15<br>[2.45, 3.84] | 2.84<br>[2.37, 3.44] | 0.65<br>[0.52, 0.81] |
| Randall | 23.98<br>[20.5, 29.01] | 7.5<br>[6.29, 9.45] | 14.65<br>[12.59, 17.62] | 1.78<br>[1.52, 2.16] | 3.93<br>[3.3, 4.74] | 1.92<br>[1.57, 2.42] | 1.75<br>[1.49, 2.06] | 0.26<br>[0.22, 0.32] |
| Reagan | 9.35<br>[8.21, 10.86] | 2.8<br>[2.37, 3.33] | 6.18<br>[5.46, 7.16] | 0.35<br>[0.31, 0.41] | 1.63<br>[1.41, 1.93] | 0.75<br>[0.62, 0.91] | 0.83<br>[0.73, 0.97] | 0.05<br>[0.05, 0.06] |
| Red River | 11.64<br>[10.09, 13.73] | 4.2<br>[3.52, 5.15] | 6.4<br>[5.58, 7.39] | 1.08<br>[0.93, 1.28] | 2.12<br>[1.82, 2.49] | 1.11<br>[0.92, 1.36] | 0.85<br>[0.75, 0.98] | 0.16<br>[0.14, 0.19] |
| Reeves | 10.98<br>[9.57, 12.72] | 3.65<br>[2.96, 4.25] | 6.79<br>[6, 7.83] | 0.55<br>[0.48, 0.64] | 1.94<br>[1.61, 2.23] | 0.96<br>[0.76, 1.12] | 0.89<br>[0.78, 1.03] | 0.08<br>[0.07, 0.1] |
| Refugio | 15.76<br>[13.51, 18.7] | 3.92<br>[3.26, 4.91] | 10.52<br>[9.02, 12.27] | 1.32<br>[1.11, 1.57] | 2.52<br>[2.11, 3] | 1.02<br>[0.82, 1.26] | 1.31<br>[1.11, 1.53] | 0.2<br>[0.16, 0.23] |
| Roberts | 7.08<br>[6.18, 8.17] | 2.18<br>[1.83, 2.64] | 4.58<br>[4.09, 5.18] | 0.33<br>[0.29, 0.38] | 1.29<br>[1.12, 1.49] | 0.59<br>[0.49, 0.69] | 0.65<br>[0.58, 0.74] | 0.05<br>[0.04, 0.06] |
| Robertson | 11.35<br>[9.52, 12.95] | 3.72<br>[3.02, 4.38] | 6.92<br>[5.92, 7.94] | 0.67<br>[0.56, 0.76] | 1.99<br>[1.66, 2.28] | 0.98<br>[0.81, 1.19] | 0.91<br>[0.78, 1.04] | 0.1<br>[0.08, 0.12] |
| Rockwall | 20.04<br>[16.83, 23.86] | 7.03<br>[5.77, 8.52] | 11.38<br>[9.62, 13.18] | 1.68<br>[1.4, 1.98] | 3.41<br>[2.83, 4.09] | 1.8<br>[1.44, 2.21] | 1.39<br>[1.15, 1.62] | 0.25<br>[0.2, 0.29] |
| Runnels | 12.83<br>[11.1, 14.61] | 4.16<br>[3.49, 4.84] | 7.77<br>[6.72, 8.89] | 0.88<br>[0.75, 1.01] | 2.21<br>[1.89, 2.54] | 1.09<br>[0.9, 1.27] | 1<br>[0.87, 1.16] | 0.13<br>[0.11, 0.15] |
| Rusk | 13.5<br>[11.71, 16.42] | 4.11<br>[3.44, 5.17] | 8.39<br>[7.32, 10.29] | 1.02<br>[0.88, 1.26] | 2.29<br>[1.96, 2.83] | 1.07<br>[0.89, 1.34] | 1.07<br>[0.91, 1.32] | 0.15<br>[0.13, 0.19] |
| Sabine | 18.12<br>[15.32, 21.31] | 6.5<br>[5.4, 8.01] | 9.59<br>[8.25, 11.36] | 1.92<br>[1.62, 2.27] | 3.15<br>[2.66, 3.83] | 1.67<br>[1.36, 2.12] | 1.2<br>[1.03, 1.44] | 0.28<br>[0.24, 0.34] |
| San Augustine | 13.92<br>[11.8, 16.13] | 4.97<br>[4.07, 6.03] | 7.51<br>[6.49, 8.5] | 1.4<br>[1.18, 1.62] | 2.48<br>[2.05, 2.94] | 1.3<br>[1.04, 1.6] | 0.97<br>[0.83, 1.11] | 0.21<br>[0.18, 0.25] |
| San Jacinto | 14.39<br>[12.23, 17.34] | 4.52<br>[3.61, 5.65] | 8.71<br>[7.46, 10.23] | 1.18<br>[0.99, 1.41] | 2.46<br>[2.07, 2.95] | 1.17<br>[0.95, 1.47] | 1.11<br>[0.94, 1.3] | 0.18<br>[0.15, 0.21] |
| San Patricio | 12.26<br>[10.55, 14.64] | 3.74<br>[3.14, 4.53] | 7.83<br>[6.74, 9.23] | 0.66<br>[0.56, 0.79] | 2.09<br>[1.77, 2.58] | 0.98<br>[0.81, 1.23] | 1.01<br>[0.87, 1.2] | 0.1<br>[0.08, 0.12] |
| San Saba | 14.85<br>[12.79, 17.23] | 4.23<br>[3.45, 4.97] | 9.28<br>[8.13, 10.72] | 1.38<br>[1.18, 1.6] | 2.46<br>[2.12, 2.93] | 1.1<br>[0.89, 1.32] | 1.17<br>[1.01, 1.38] | 0.21<br>[0.17, 0.24] |
| Schleicher | 21.76<br>[18.86, 25.74] | 4.81<br>[4.08, 5.93] | 16.16<br>[14.01, 18.96] | 0.78<br>[0.67, 0.94] | 3.27<br>[2.78, 3.95] | 1.23<br>[1.03, 1.57] | 1.91<br>[1.63, 2.29] | 0.11<br>[0.1, 0.14] |
| Scurry | 13.65<br>[11.73, 15.86] | 3.69<br>[3.01, 4.39] | 9.33<br>[8.05, 10.65] | 0.63<br>[0.54, 0.73] | 2.23<br>[1.88, 2.61] | 0.96<br>[0.78, 1.14] | 1.17<br>[1, 1.36] | 0.09<br>[0.08, 0.11] |
| Shackelford | 33.5<br>[28.49, 40.34] | 10.53<br>[8.76, 12.71] | 19.29<br>[16.38, 22.88] | 3.8<br>[3.18, 4.61] | 5.45<br>[4.59, 6.61] | 2.68<br>[2.23, 3.31] | 2.22<br>[1.88, 2.66] | 0.54<br>[0.45, 0.69] |
| Shelby | 13.23<br>[11.41, 15.42] | 4.57<br>[3.77, 5.52] | 7.86<br>[6.88, 8.95] | 0.78<br>[0.67, 0.91] | 2.33<br>[1.95, 2.73] | 1.2<br>[0.97, 1.46] | 1.01<br>[0.87, 1.15] | 0.12<br>[0.1, 0.14] |
| Sherman | 16.76<br>[14.35, 19.56] | 5.42<br>[4.45, 6.43] | 10.4<br>[8.99, 11.97] | 1<br>[0.86, 1.16] | 2.86<br>[2.38, 3.36] | 1.41<br>[1.14, 1.72] | 1.3<br>[1.13, 1.51] | 0.15<br>[0.12, 0.18] |
| Smith | 21.18<br>[17.84, 25.3] | 6.85<br>[5.73, 8.38] | 12.74<br>[10.85, 14.88] | 1.55<br>[1.32, 1.85] | 3.49<br>[2.91, 4.23] | 1.75<br>[1.43, 2.19] | 1.52<br>[1.29, 1.82] | 0.23<br>[0.18, 0.28] |
| Somervell | 20.39<br>[17.09, 23.87] | 6.94<br>[5.55, 8.34] | 11.28<br>[9.58, 12.93] | 2.16<br>[1.8, 2.52] | 3.48<br>[2.86, 4.11] | 1.78<br>[1.42, 2.14] | 1.38<br>[1.17, 1.6] | 0.32<br>[0.25, 0.38] |
| Starr | 9.31<br>[7.94, 10.92] | 2.81<br>[2.28, 3.42] | 6.23<br>[5.42, 7.24] | 0.3<br>[0.26, 0.35] | 1.63<br>[1.35, 1.92] | 0.74<br>[0.59, 0.9] | 0.83<br>[0.71, 0.98] | 0.05<br>[0.04, 0.05] |
| Stephens | 35.05<br>[29.15, 40.26] | 10.34<br>[8.49, 12.65] | 21.07<br>[17.57, 24.23] | 3.52<br>[2.94, 4.07] | 5.53<br>[4.54, 6.46] | 2.62<br>[2.12, 3.21] | 2.39<br>[1.96, 2.81] | 0.5<br>[0.41, 0.6] |
| Sterling | 27.34<br>[23.35, 31.76] | 11.11<br>[9.25, 12.95] | 14.23<br>[12.44, 16.3] | 2.01<br>[1.73, 2.36] | 4.89<br>[4.1, 5.71] | 2.87<br>[2.37, 3.4] | 1.72<br>[1.48, 2.03] | 0.3<br>[0.25, 0.35] |
| Sutton | 12.73<br>[10.85, 14.85] | 3.72<br>[3.03, 4.49] | 8.06<br>[7, 9.3] | 0.91<br>[0.78, 1.07] | 2.16<br>[1.79, 2.51] | 0.98<br>[0.8, 1.18] | 1.04<br>[0.88, 1.21] | 0.14<br>[0.11, 0.16] |
| Swisher | 15.79<br>[13.49, 18.6] | 4.4<br>[3.59, 5.38] | 10.36<br>[8.98, 12.02] | 0.99<br>[0.84, 1.17] | 2.56<br>[2.19, 3.07] | 1.14<br>[0.93, 1.39] | 1.28<br>[1.1, 1.52] | 0.15<br>[0.12, 0.17] |
| Tarrant | 19.39<br>[16.18, 23.52] | 6.49<br>[5.2, 7.89] | 11.48<br>[9.63, 13.83] | 1.39<br>[1.16, 1.68] | 3.27<br>[2.66, 4] | 1.66<br>[1.31, 2.04] | 1.39<br>[1.14, 1.67] | 0.2<br>[0.17, 0.25] |
| Taylor | 24.39<br>[20.86, 30.07] | 7<br>[5.63, 8.77] | 16.22<br>[13.84, 19.8] | 1.3<br>[1.09, 1.6] | 3.89<br>[3.26, 4.81] | 1.78<br>[1.44, 2.17] | 1.92<br>[1.61, 2.34] | 0.19<br>[0.16, 0.23] |
| Terrell | 7.68<br>[6.81, 8.49] | 1.65<br>[1.37, 1.94] | 5.58<br>[5, 6.09] | 0.47<br>[0.41, 0.52] | 1.28<br>[1.11, 1.44] | 0.44<br>[0.36, 0.52] | 0.77<br>[0.67, 0.86] | 0.07<br>[0.06, 0.08] |
| Terry | 14.55<br>[12.35, 17.25] | 5.17<br>[4.14, 6.14] | 8.56<br>[7.37, 10.17] | 0.81<br>[0.68, 0.96] | 2.56<br>[2.1, 3.04] | 1.35<br>[1.08, 1.62] | 1.09<br>[0.91, 1.28] | 0.12<br>[0.1, 0.14] |
| Throckmorton | 26.34<br>[23.13, 30.1] | 9.89<br>[8.52, 11.46] | 13.3<br>[11.7, 15.09] | 3.16<br>[2.75, 3.67] | 4.64<br>[4.04, 5.43] | 2.58<br>[2.18, 3.04] | 1.61<br>[1.39, 1.88] | 0.47<br>[0.4, 0.55] |
| Titus | 12.07<br>[10.48, 14.4] | 3.79<br>[3.21, 4.56] | 7.7<br>[6.67, 9.09] | 0.59<br>[0.51, 0.71] | 2.09<br>[1.78, 2.45] | 1<br>[0.82, 1.2] | 0.99<br>[0.86, 1.19] | 0.09<br>[0.08, 0.11] |
| Tom Green | 17.26<br>[14.46, 20.05] | 4.94<br>[4.04, 6.06] | 11.32<br>[9.57, 13.04] | 0.98<br>[0.81, 1.14] | 2.8<br>[2.34, 3.33] | 1.27<br>[1.05, 1.57] | 1.38<br>[1.19, 1.62] | 0.14<br>[0.12, 0.17] |
| Travis | 28.12<br>[23.53, 33.71] | 9.49<br>[7.69, 11.58] | 16.07<br>[13.49, 18.99] | 2.62<br>[2.16, 3.14] | 4.65<br>[3.82, 5.6] | 2.4<br>[1.96, 2.96] | 1.88<br>[1.54, 2.24] | 0.38<br>[0.31, 0.46] |
| Trinity | 16.67<br>[14.33, 19.14] | 5.23<br>[4.36, 6.21] | 9.87<br>[8.52, 11.28] | 1.55<br>[1.33, 1.82] | 2.82<br>[2.41, 3.29] | 1.37<br>[1.12, 1.61] | 1.23<br>[1.05, 1.4] | 0.23<br>[0.19, 0.27] |
| Tyler | 20.23<br>[17.04, 24.71] | 6.66<br>[5.42, 8.54] | 11.45<br>[9.65, 13.66] | 2.12<br>[1.78, 2.59] | 3.43<br>[2.85, 4.28] | 1.72<br>[1.4, 2.22] | 1.4<br>[1.19, 1.65] | 0.31<br>[0.25, 0.39] |
| Upshur | 17.8<br>[15.03, 21.12] | 6.18<br>[5.01, 7.55] | 10.13<br>[8.64, 11.88] | 1.5<br>[1.25, 1.78] | 3.08<br>[2.58, 3.74] | 1.6<br>[1.28, 1.97] | 1.24<br>[1.06, 1.5] | 0.22<br>[0.18, 0.27] |
| Upton | 13.96<br>[12.08, 16.55] | 3.23<br>[2.64, 3.9] | 9.97<br>[8.65, 11.83] | 0.75<br>[0.64, 0.9] | 2.2<br>[1.87, 2.59] | 0.84<br>[0.7, 1.02] | 1.25<br>[1.09, 1.49] | 0.11<br>[0.09, 0.13] |
| Uvalde | 17.72<br>[15.37, 20.97] | 5.66<br>[4.86, 6.94] | 11.18<br>[9.75, 13.06] | 0.86<br>[0.74, 1.01] | 2.95<br>[2.53, 3.56] | 1.46<br>[1.23, 1.76] | 1.37<br>[1.17, 1.66] | 0.13<br>[0.11, 0.15] |
| Val Verde | 12.36<br>[10.32, 14.11] | 4.01<br>[3.2, 4.64] | 7.83<br>[6.62, 8.98] | 0.53<br>[0.44, 0.61] | 2.15<br>[1.74, 2.46] | 1.05<br>[0.83, 1.23] | 1.01<br>[0.84, 1.16] | 0.08<br>[0.06, 0.09] |
| Van Zandt | 20.21<br>[16.57, 23.51] | 7<br>[5.64, 8.3] | 11.3<br>[9.47, 13.07] | 1.87<br>[1.54, 2.16] | 3.42<br>[2.81, 4.08] | 1.79<br>[1.43, 2.2] | 1.38<br>[1.16, 1.61] | 0.28<br>[0.22, 0.32] |
| Victoria | 12.56<br>[10.68, 14.78] | 3.97<br>[3.26, 4.81] | 7.89<br>[6.82, 9.15] | 0.7<br>[0.59, 0.82] | 2.17<br>[1.81, 2.59] | 1.04<br>[0.85, 1.29] | 1.02<br>[0.86, 1.19] | 0.1<br>[0.09, 0.13] |
| Walker | 116.38<br>[93.79, 141.53] | 22.65<br>[18.1, 28.41] | 83.67<br>[67.25, 101.88] | 9.6<br>[7.63, 11.79] | 15.29<br>[12.17, 19.03] | 5.4<br>[4.31, 6.84] | 8.63<br>[6.83, 10.62] | 1.29<br>[1.01, 1.62] |
| Waller | 86.76<br>[70.74, 110.39] | 14.33<br>[11.94, 18.77] | 69.24<br>[56.29, 87.59] | 2.97<br>[2.4, 3.82] | 11.04<br>[9.16, 13.99] | 3.44<br>[2.83, 4.45] | 7.22<br>[5.85, 9.19] | 0.4<br>[0.32, 0.52] |
| Ward | 12.77<br>[10.77, 14.9] | 4.45<br>[3.62, 5.41] | 7.64<br>[6.58, 8.88] | 0.69<br>[0.59, 0.81] | 2.26<br>[1.88, 2.71] | 1.16<br>[0.94, 1.44] | 0.99<br>[0.84, 1.16] | 0.1<br>[0.09, 0.12] |
| Washington | 52.22<br>[42.24, 62.82] | 10.28<br>[7.93, 12.53] | 39.17<br>[31.57, 46.86] | 2.71<br>[2.16, 3.27] | 7.14<br>[5.71, 8.78] | 2.51<br>[1.96, 3.1] | 4.25<br>[3.43, 5.29] | 0.38<br>[0.3, 0.46] |
| Webb | 10.51<br>[9.13, 12.66] | 2.99<br>[2.47, 3.64] | 7.17<br>[6.28, 8.54] | 0.34<br>[0.3, 0.41] | 1.77<br>[1.52, 2.13] | 0.78<br>[0.64, 0.97] | 0.94<br>[0.81, 1.12] | 0.05<br>[0.04, 0.06] |
| Wharton | 13.1<br>[11.24, 15.52] | 3.99<br>[3.24, 4.76] | 8.47<br>[7.36, 10.02] | 0.64<br>[0.55, 0.77] | 2.22<br>[1.85, 2.66] | 1.04<br>[0.83, 1.28] | 1.08<br>[0.93, 1.3] | 0.1<br>[0.08, 0.12] |
| Wheeler | 14.45<br>[12.4, 17.19] | 4.54<br>[3.77, 5.72] | 8.78<br>[7.72, 10.43] | 1.11<br>[0.95, 1.32] | 2.46<br>[2.1, 2.99] | 1.17<br>[0.98, 1.5] | 1.11<br>[0.97, 1.32] | 0.17<br>[0.14, 0.2] |
| Wichita | 27.95<br>[23.36, 33.38] | 7.18<br>[5.85, 8.95] | 19.34<br>[16.3, 22.84] | 1.45<br>[1.2, 1.73] | 4.23<br>[3.46, 5.22] | 1.8<br>[1.44, 2.29] | 2.22<br>[1.83, 2.73] | 0.21<br>[0.17, 0.25] |
| Wilbarger | 19.38<br>[16.33, 23.17] | 5.27<br>[4.26, 6.25] | 13.06<br>[11.08, 15.64] | 1.05<br>[0.87, 1.27] | 3.1<br>[2.55, 3.69] | 1.35<br>[1.1, 1.63] | 1.58<br>[1.34, 1.93] | 0.15<br>[0.13, 0.19] |
| Willacy | 15.04<br>[12.91, 17.33] | 3.46<br>[2.9, 4.08] | 10.91<br>[9.44, 12.63] | 0.62<br>[0.54, 0.72] | 2.34<br>[2, 2.72] | 0.9<br>[0.76, 1.06] | 1.36<br>[1.16, 1.56] | 0.09<br>[0.08, 0.11] |
| Williamson | 19.81<br>[16.78, 23.04] | 7.31<br>[6.03, 8.84] | 10.7<br>[9.25, 12.44] | 1.79<br>[1.52, 2.09] | 3.45<br>[2.88, 4.05] | 1.88<br>[1.52, 2.32] | 1.31<br>[1.12, 1.55] | 0.26<br>[0.22, 0.32] |
| Wilson | 11.69<br>[10.3, 13.37] | 3.52<br>[3.03, 4.13] | 7.34<br>[6.48, 8.42] | 0.8<br>[0.7, 0.92] | 2.01<br>[1.73, 2.31] | 0.93<br>[0.78, 1.12] | 0.96<br>[0.82, 1.11] | 0.12<br>[0.1, 0.14] |
| Winkler | 11.64<br>[9.91, 13.45] | 3.63<br>[2.96, 4.3] | 7.56<br>[6.49, 8.65] | 0.49<br>[0.42, 0.57] | 2<br>[1.68, 2.33] | 0.95<br>[0.78, 1.13] | 0.98<br>[0.83, 1.13] | 0.07<br>[0.06, 0.09] |
| Wise | 21.93<br>[18.15, 26.26] | 8.17<br>[6.33, 10.23] | 11.78<br>[10.13, 13.87] | 2<br>[1.66, 2.38] | 3.8<br>[3.06, 4.58] | 2.08<br>[1.6, 2.61] | 1.43<br>[1.2, 1.7] | 0.29<br>[0.24, 0.35] |
| Wood | 21.24<br>[18.21, 24.86] | 6.02<br>[5.05, 7.4] | 13.09<br>[11.34, 15.29] | 2.05<br>[1.76, 2.44] | 3.45<br>[2.89, 4.11] | 1.56<br>[1.29, 1.92] | 1.58<br>[1.35, 1.88] | 0.3<br>[0.26, 0.36] |
| Yoakum | 16.24<br>[13.73, 18.64] | 5.59<br>[4.57, 6.66] | 9.92<br>[8.35, 11.2] | 0.74<br>[0.62, 0.84] | 2.8<br>[2.34, 3.24] | 1.45<br>[1.19, 1.73] | 1.24<br>[1.03, 1.43] | 0.11<br>[0.09, 0.13] |
| Young | 14.01<br>[12.09, 16.97] | 4.51<br>[3.76, 5.78] | 8.47<br>[7.36, 10.1] | 0.98<br>[0.85, 1.19] | 2.41<br>[2.06, 2.95] | 1.18<br>[0.97, 1.5] | 1.08<br>[0.93, 1.28] | 0.15<br>[0.12, 0.18] |
| Zapata | 9.21<br>[7.91, 10.49] | 2.35<br>[1.91, 2.77] | 6.54<br>[5.74, 7.43] | 0.3<br>[0.25, 0.34] | 1.54<br>[1.32, 1.78] | 0.62<br>[0.5, 0.73] | 0.87<br>[0.76, 1] | 0.05<br>[0.04, 0.05] |
| Zavala | 10.39<br>[8.87, 12.33] | 2.89<br>[2.42, 3.64] | 7.18<br>[6.18, 8.44] | 0.34<br>[0.29, 0.4] | 1.76<br>[1.49, 2.12] | 0.76<br>[0.62, 0.97] | 0.93<br>[0.81, 1.11] | 0.05<br>[0.04, 0.06] |

**Table S.4.** Age-specific cumulative case counts and hospitalizations derived from 5% reduced vaccination uptake scenario projections.

| County Name | Total Cases | 0-4 Cases | 5-19 Cases | 20+ Cases | Total Hosp | 0-4 Hosp | 5-19 Hosp | 20+ Hosp |
| --- | --- | --- | --- | --- | --- | --- | --- | --- |
| Anderson | 41.1<br>[34.8, 50.6] | 16.4<br>[13.5, 21] | 19.2<br>[16.4, 23.1] | 5.6<br>[4.7, 6.8] | 7.1<br>[6, 8.8] | 4.1<br>[3.4, 5.3] | 2.2<br>[1.9, 2.7] | 0.8<br>[0.7, 1] |
| Andrews | 26.6<br>[22, 31.8] | 11.1<br>[9.2, 13.7] | 13.3<br>[11.2, 15.7] | 2<br>[1.7, 2.4] | 4.7<br>[3.8, 5.8] | 2.8<br>[2.3, 3.6] | 1.6<br>[1.3, 1.9] | 0.3<br>[0.2, 0.4] |
| Angelina | 42.8<br>[34.8, 52.5] | 17.4<br>[14.4, 22.4] | 20.9<br>[16.9, 24.9] | 4.3<br>[3.5, 5.2] | 7.3<br>[6, 9] | 4.3<br>[3.5, 5.7] | 2.4<br>[1.9, 2.8] | 0.6<br>[0.5, 0.8] |
| Aransas | 33.5<br>[28.1, 41.9] | 12.4<br>[10, 15.6] | 16.2<br>[13.8, 19.9] | 5<br>[4.2, 6.3] | 5.7<br>[4.7, 7.2] | 3.1<br>[2.5, 4] | 1.9<br>[1.6, 2.3] | 0.7<br>[0.6, 0.9] |
| Archer | 28.7<br>[24.2, 34.2] | 10.5<br>[8.7, 12.8] | 14.6<br>[12.3, 17.1] | 3.7<br>[3, 4.3] | 4.9<br>[4.1, 6] | 2.7<br>[2.2, 3.3] | 1.7<br>[1.5, 2] | 0.5<br>[0.4, 0.6] |
| Armstrong | 30.3<br>[26.4, 35.4] | 11.5<br>[9.7, 13.3] | 14.3<br>[12.5, 16.8] | 4.5<br>[3.9, 5.4] | 5.4<br>[4.6, 6.3] | 3<br>[2.5, 3.5] | 1.7<br>[1.5, 2] | 0.7<br>[0.6, 0.8] |
| Atascosa | 29.6<br>[24.6, 36.2] | 11.4<br>[9.2, 14.4] | 15.7<br>[13.1, 19] | 2.6<br>[2.2, 3.2] | 5.1<br>[4.3, 6.3] | 2.9<br>[2.3, 3.6] | 1.8<br>[1.6, 2.3] | 0.4<br>[0.3, 0.5] |
| Austin | 38.9<br>[32.4, 47.6] | 15.6<br>[12.7, 19.4] | 18.7<br>[15.8, 22.7] | 4.6<br>[3.9, 5.7] | 6.7<br>[5.4, 8.2] | 3.9<br>[3.1, 4.9] | 2.1<br>[1.7, 2.7] | 0.7<br>[0.5, 0.8] |
| Bailey | 35.2<br>[29.4, 41.4] | 13.8<br>[10.9, 16.7] | 18.8<br>[16.1, 21.9] | 2.6<br>[2.2, 3.1] | 6<br>[4.9, 7.1] | 3.5<br>[2.8, 4.2] | 2.2<br>[1.8, 2.5] | 0.4<br>[0.3, 0.4] |
| Bandera | 46.7<br>[38.1, 57] | 16.9<br>[13.1, 21.4] | 21.1<br>[18, 25.8] | 8.5<br>[7.1, 10.5] | 7.9<br>[6.2, 9.8] | 4.2<br>[3.2, 5.3] | 2.4<br>[2, 2.9] | 1.2<br>[1, 1.5] |
| Bastrop | 44.2<br>[36.3, 54.2] | 17.9<br>[14, 22.6] | 21.7<br>[18.2, 26.1] | 4.5<br>[3.7, 5.5] | 7.6<br>[6.1, 9.2] | 4.4<br>[3.5, 5.6] | 2.5<br>[2.1, 3] | 0.6<br>[0.5, 0.8] |
| Baylor | 28.9<br>[24.9, 34.1] | 10.8<br>[9.2, 13.1] | 14.3<br>[12.3, 16.6] | 3.7<br>[3.1, 4.3] | 5<br>[4.3, 6.1] | 2.8<br>[2.4, 3.4] | 1.7<br>[1.4, 2] | 0.5<br>[0.4, 0.6] |
| Bee | 45.4<br>[37.2, 54.5] | 14.9<br>[11.8, 18.3] | 25.5<br>[21.3, 30.4] | 5<br>[4.1, 6] | 7.3<br>[5.8, 8.9] | 3.7<br>[2.9, 4.7] | 2.8<br>[2.4, 3.5] | 0.7<br>[0.6, 0.9] |
| Bell | 39.8<br>[33.4, 48] | 17.8<br>[14.4, 21.9] | 18.4<br>[15.8, 22.1] | 3.6<br>[3, 4.3] | 7<br>[5.8, 8.8] | 4.4<br>[3.5, 5.6] | 2.1<br>[1.8, 2.6] | 0.5<br>[0.4, 0.6] |
| Bexar | 43.9<br>[35.7, 52.3] | 16.9<br>[13.7, 21] | 22.6<br>[18.5, 26.4] | 4.3<br>[3.5, 5.2] | 7.3<br>[5.9, 8.9] | 4.2<br>[3.3, 5.2] | 2.5<br>[2.1, 3] | 0.6<br>[0.5, 0.8] |
| Blanco | 108.6<br>[91, 132.5] | 36.4<br>[29.7, 45.4] | 47.9<br>[40.3, 57.6] | 24.3<br>[20.1, 29.9] | 17.6<br>[14.3, 21.6] | 9<br>[7.3, 11.5] | 5.1<br>[4.2, 6.3] | 3.3<br>[2.7, 4.2] |
| Borden | 24.5<br>[22.3, 26.6] | 5.5<br>[5, 6] | 14.6<br>[13.4, 15.9] | 4.3<br>[3.9, 4.9] | 3.9<br>[3.5, 4.4] | 1.5<br>[1.3, 1.7] | 1.8<br>[1.6, 2] | 0.6<br>[0.6, 0.8] |
| Bosque | 40.7<br>[33.7, 49.2] | 15.7<br>[12.4, 19.5] | 19.1<br>[16.2, 22.6] | 5.8<br>[4.9, 7.1] | 7<br>[5.6, 8.3] | 3.9<br>[3, 4.7] | 2.2<br>[1.8, 2.6] | 0.8<br>[0.7, 1] |
| Bowie | 42.7<br>[34.3, 52.5] | 17.2<br>[13.1, 21.8] | 20.6<br>[16.9, 25] | 5<br>[4, 6.1] | 7.3<br>[5.8, 9.3] | 4.3<br>[3.2, 5.5] | 2.3<br>[1.9, 2.9] | 0.7<br>[0.5, 0.9] |
| Brazoria | 34.9<br>[28.9, 42.4] | 14<br>[11.2, 17.6] | 17.4<br>[14.6, 20.7] | 3.6<br>[3, 4.3] | 6<br>[4.9, 7.4] | 3.5<br>[2.8, 4.5] | 2<br>[1.7, 2.4] | 0.5<br>[0.4, 0.6] |
| Brazos | 260.3<br>[195.5, 344] | 56.6<br>[41.7, 74.6] | 184.4<br>[139.5, 244.6] | 18.4<br>[13.8, 24.3] | 33.5<br>[25.6, 45.3] | 13<br>[9.6, 18.3] | 18<br>[13.6, 23.7] | 2.4<br>[1.8, 3.1] |
| Brewster | 189.5<br>[160.6, 228] | 45.7<br>[38, 54.4] | 116.8<br>[97.9, 140.3] | 27.7<br>[23.1, 34.3] | 26.9<br>[22.6, 33] | 11.2<br>[9.3, 13.7] | 12.1<br>[10, 14.6] | 3.7<br>[3, 4.6] |
| Briscoe | 32.2<br>[28.2, 37.1] | 11.5<br>[9.8, 13.4] | 15<br>[13.4, 17.1] | 5.7<br>[4.9, 6.6] | 5.6<br>[4.9, 6.5] | 3<br>[2.6, 3.5] | 1.8<br>[1.5, 2.1] | 0.8<br>[0.7, 1] |
| Brooks | 27.2<br>[22.5, 32.7] | 10.1<br>[8, 12.4] | 14.6<br>[12.4, 17.4] | 2.5<br>[2, 3] | 4.7<br>[3.7, 5.7] | 2.6<br>[2, 3.2] | 1.7<br>[1.4, 2.1] | 0.4<br>[0.3, 0.4] |
| Brown | 60.6<br>[50.8, 73.4] | 22.6<br>[18, 27.7] | 30.6<br>[25.4, 36.7] | 7.5<br>[6.2, 9.1] | 9.9<br>[8.3, 12.2] | 5.5<br>[4.5, 7] | 3.4<br>[2.8, 4.1] | 1<br>[0.9, 1.3] |
| Burleson | 24.5<br>[21, 30.1] | 9.9<br>[8.1, 12.4] | 11.9<br>[10.4, 14.4] | 2.8<br>[2.4, 3.4] | 4.3<br>[3.6, 5.4] | 2.5<br>[2, 3.1] | 1.4<br>[1.2, 1.8] | 0.4<br>[0.3, 0.5] |
| Burnet | 67.1<br>[55.3, 81.7] | 27.2<br>[22, 34.1] | 29.7<br>[24.4, 35.5] | 10.4<br>[8.4, 12.5] | 11.4<br>[9.4, 13.9] | 6.7<br>[5.4, 8.5] | 3.2<br>[2.7, 4] | 1.4<br>[1.2, 1.8] |
| Caldwell | 54.1<br>[43.5, 67.9] | 20.7<br>[15.9, 26.5] | 27.8<br>[23.2, 34.3] | 5.6<br>[4.5, 7.1] | 9<br>[7.2, 11.2] | 5.1<br>[4, 6.5] | 3.1<br>[2.6, 3.8] | 0.8<br>[0.6, 1] |
| Calhoun | 33.3<br>[26.4, 39.8] | 12.7<br>[9.6, 15.5] | 17.3<br>[14.3, 20.4] | 3.4<br>[2.8, 4] | 5.7<br>[4.3, 6.9] | 3.2<br>[2.3, 4] | 2<br>[1.7, 2.4] | 0.5<br>[0.4, 0.6] |
| Callahan | 41.2<br>[34.5, 51.1] | 15.9<br>[13.3, 20.6] | 19.4<br>[16.5, 23.8] | 5.8<br>[4.9, 7.2] | 7<br>[5.9, 8.9] | 4<br>[3.3, 5.1] | 2.2<br>[1.9, 2.7] | 0.8<br>[0.7, 1] |
| Cameron | 31.6<br>[26.1, 38.6] | 11.3<br>[9, 14.3] | 17.7<br>[14.9, 21.5] | 2.4<br>[2, 2.9] | 5.2<br>[4.4, 6.4] | 2.8<br>[2.3, 3.6] | 2<br>[1.7, 2.5] | 0.3<br>[0.3, 0.4] |
| Camp | 31<br>[26, 38] | 12.8<br>[10.5, 16.1] | 15.5<br>[13.1, 18.7] | 2.8<br>[2.3, 3.4] | 5.5<br>[4.5, 6.8] | 3.2<br>[2.6, 4.1] | 1.8<br>[1.5, 2.2] | 0.4<br>[0.3, 0.5] |
| Carson | 48.8<br>[41, 59.9] | 17.1<br>[13.6, 20.9] | 24.8<br>[20.9, 30.2] | 7<br>[5.8, 8.8] | 8.1<br>[6.6, 9.9] | 4.3<br>[3.4, 5.3] | 2.8<br>[2.3, 3.4] | 1<br>[0.8, 1.2] |
| Cass | 39.4<br>[32.2, 48.7] | 16.2<br>[13, 20.9] | 18.2<br>[15.2, 22.1] | 4.8<br>[4, 5.9] | 6.8<br>[5.6, 8.7] | 4<br>[3.3, 5.2] | 2.1<br>[1.8, 2.6] | 0.7<br>[0.6, 0.9] |
| Castro | 29.9<br>[24.5, 35.6] | 11.8<br>[9.3, 14.6] | 15.7<br>[13.1, 18.6] | 2.4<br>[1.9, 2.8] | 5.2<br>[4.3, 6.2] | 3<br>[2.4, 3.8] | 1.8<br>[1.5, 2.2] | 0.3<br>[0.3, 0.4] |
| Chambers | 39<br>[32.4, 47.5] | 16.3<br>[13.1, 20.1] | 19<br>[15.9, 22.5] | 3.9<br>[3.2, 4.7] | 6.8<br>[5.5, 8.3] | 4.1<br>[3.2, 5.1] | 2.2<br>[1.8, 2.6] | 0.6<br>[0.5, 0.7] |
| Cherokee | 28.6<br>[24.4, 35] | 11.7<br>[9.8, 14.8] | 14.1<br>[12.2, 17.2] | 2.7<br>[2.3, 3.4] | 5<br>[4.2, 6.3] | 3<br>[2.5, 3.8] | 1.7<br>[1.4, 2.1] | 0.4<br>[0.3, 0.5] |
| Childress | 54.8<br>[45.7, 64.1] | 18.1<br>[14.6, 21.9] | 28.4<br>[23.9, 33.3] | 8.4<br>[7, 9.9] | 8.9<br>[7.2, 10.7] | 4.5<br>[3.6, 5.6] | 3.1<br>[2.7, 3.8] | 1.2<br>[1, 1.4] |
| Clay | 36.3<br>[29.4, 43] | 13.5<br>[10.6, 16.5] | 17.8<br>[15, 21.2] | 5<br>[4.1, 6.1] | 6.1<br>[5, 7.4] | 3.4<br>[2.7, 4.2] | 2.1<br>[1.7, 2.5] | 0.7<br>[0.6, 0.9] |
| Cochran | 42.3<br>[35.1, 48.3] | 18.4<br>[15, 21.5] | 19.3<br>[16.2, 22.3] | 4.5<br>[3.7, 5.3] | 7.6<br>[6.3, 8.9] | 4.8<br>[3.9, 5.6] | 2.2<br>[1.9, 2.6] | 0.6<br>[0.5, 0.8] |
| Coke | 37.8<br>[32.7, 44.7] | 15.6<br>[13.1, 18.8] | 15.5<br>[13.4, 18.3] | 6.7<br>[5.7, 8] | 6.8<br>[5.8, 8.2] | 4<br>[3.3, 4.9] | 1.8<br>[1.6, 2.3] | 1<br>[0.8, 1.2] |
| Coleman | 40.9<br>[34, 48.9] | 16.2<br>[12.9, 19.7] | 19<br>[15.8, 22.3] | 5.8<br>[4.7, 6.8] | 7.1<br>[5.8, 8.5] | 4.1<br>[3.2, 4.9] | 2.2<br>[1.8, 2.6] | 0.8<br>[0.7, 1] |
| Collin | 63.8<br>[51.7, 78.8] | 24.9<br>[19.7, 31.2] | 31.3<br>[25.9, 38.6] | 7.6<br>[6.2, 9.3] | 10.6<br>[8.6, 13] | 6.1<br>[4.8, 7.9] | 3.4<br>[2.9, 4.2] | 1<br>[0.9, 1.3] |
| Collingsworth | 40.2<br>[34.7, 47.9] | 15.1<br>[12.7, 17.8] | 20.2<br>[17.5, 24] | 5.1<br>[4.3, 6.1] | 6.9<br>[5.9, 8.3] | 3.9<br>[3.3, 4.7] | 2.3<br>[2, 2.7] | 0.7<br>[0.6, 0.9] |
| Colorado | 27.9<br>[23.2, 32.8] | 12.2<br>[9.7, 14.8] | 12.6<br>[10.8, 14.6] | 3<br>[2.5, 3.5] | 5.1<br>[4.2, 6.1] | 3.1<br>[2.5, 3.8] | 1.5<br>[1.3, 1.8] | 0.4<br>[0.4, 0.5] |
| Comal | 54<br>[44.5, 71] | 21.6<br>[17.5, 30.1] | 24.7<br>[20.7, 30.9] | 7.4<br>[6.1, 9.7] | 9.1<br>[7.4, 11.9] | 5.4<br>[4.3, 7.3] | 2.8<br>[2.3, 3.4] | 1<br>[0.9, 1.3] |
| Comanche | 37.8<br>[31.6, 44.5] | 14.8<br>[11.9, 18] | 18.7<br>[15.8, 21.9] | 4.3<br>[3.5, 5] | 6.5<br>[5.3, 7.8] | 3.7<br>[3, 4.5] | 2.2<br>[1.8, 2.5] | 0.6<br>[0.5, 0.7] |
| Concho | 67.9<br>[58.8, 77.2] | 16.1<br>[14, 18.7] | 39.6<br>[34.4, 44.7] | 12<br>[10.1, 13.9] | 10.3<br>[8.9, 12] | 4.1<br>[3.5, 4.9] | 4.5<br>[3.9, 5.1] | 1.7<br>[1.4, 2.1] |
| Cooke | 49.1<br>[39.9, 58.7] | 20.1<br>[16.2, 24.6] | 23.2<br>[19.3, 28] | 5.7<br>[4.7, 6.9] | 8.5<br>[6.9, 10.3] | 5<br>[4.1, 6.2] | 2.6<br>[2.2, 3.1] | 0.8<br>[0.6, 1] |
| Coryell | 45.4<br>[38.5, 56.3] | 20<br>[16.6, 24.8] | 20.1<br>[17.3, 24.6] | 5.4<br>[4.6, 6.7] | 8<br>[6.7, 10.1] | 5<br>[4.1, 6.3] | 2.3<br>[2, 2.8] | 0.8<br>[0.6, 0.9] |
| Cottle | 39.2<br>[33.9, 43.8] | 8.8<br>[7.7, 10] | 24.5<br>[21.3, 27.2] | 5.8<br>[4.9, 6.7] | 6<br>[5.1, 6.9] | 2.3<br>[2, 2.7] | 2.9<br>[2.4, 3.3] | 0.8<br>[0.7, 1] |
| Crockett | 23.8<br>[20.6, 28.1] | 9<br>[7.6, 10.9] | 12.6<br>[11, 14.6] | 2.2<br>[1.9, 2.6] | 4.2<br>[3.5, 5.1] | 2.3<br>[1.9, 2.9] | 1.5<br>[1.3, 1.8] | 0.3<br>[0.3, 0.4] |
| Crosby | 38<br>[32.2, 44.2] | 12.6<br>[10.5, 15.2] | 21.8<br>[18.5, 25.5] | 3.6<br>[3, 4.3] | 6.1<br>[5.2, 7.4] | 3.2<br>[2.6, 3.9] | 2.5<br>[2.1, 2.9] | 0.5<br>[0.4, 0.6] |
| Culberson | 46<br>[39.2, 53.6] | 14.5<br>[12.2, 17] | 26.9<br>[23.1, 31.4] | 4.6<br>[3.8, 5.5] | 7.5<br>[6.2, 8.9] | 3.7<br>[3, 4.4] | 3.1<br>[2.6, 3.6] | 0.7<br>[0.5, 0.8] |
| Dallam | 37.4<br>[31.3, 46.4] | 19.5<br>[15.9, 24.5] | 15.3<br>[12.8, 18.6] | 2.6<br>[2.2, 3.3] | 7.1<br>[5.7, 9] | 5<br>[3.8, 6.4] | 1.8<br>[1.5, 2.2] | 0.4<br>[0.3, 0.5] |
| Dallas | 41.3<br>[33.8, 51.8] | 17<br>[13.3, 21.6] | 20.3<br>[16.9, 25.2] | 4.1<br>[3.4, 5.2] | 7.1<br>[5.7, 8.9] | 4.2<br>[3.2, 5.5] | 2.3<br>[1.9, 2.8] | 0.6<br>[0.5, 0.7] |
| Dawson | 63.8<br>[53.1, 78.5] | 29.3<br>[23.9, 37] | 27.3<br>[22.7, 32.5] | 7.5<br>[6.2, 9.2] | 11.4<br>[9.2, 14.1] | 7.3<br>[5.9, 9.3] | 3<br>[2.5, 3.7] | 1.1<br>[0.8, 1.3] |
| DeWitt | 29<br>[24.8, 34.9] | 11.3<br>[9.2, 13.9] | 14.6<br>[12.4, 17.4] | 3.3<br>[2.8, 4] | 5<br>[4.2, 6.2] | 2.9<br>[2.3, 3.6] | 1.7<br>[1.5, 2.1] | 0.5<br>[0.4, 0.6] |
| Deaf Smith | 27.6<br>[22.8, 33.5] | 11.7<br>[9.3, 14.2] | 14.2<br>[11.8, 16.9] | 1.8<br>[1.5, 2.2] | 4.9<br>[3.9, 6.1] | 3<br>[2.3, 3.7] | 1.7<br>[1.4, 2] | 0.3<br>[0.2, 0.3] |
| Delta | 25.7<br>[21.3, 30.7] | 11.2<br>[8.8, 13.7] | 11.5<br>[9.6, 13.5] | 3<br>[2.4, 3.5] | 4.7<br>[3.8, 5.7] | 2.9<br>[2.2, 3.6] | 1.4<br>[1.2, 1.7] | 0.4<br>[0.4, 0.5] |
| Denton | 85<br>[66.5, 109.3] | 35.2<br>[26.3, 45.8] | 38.4<br>[29.9, 49.1] | 11.2<br>[8.7, 14.5] | 14.2<br>[10.8, 18.5] | 8.6<br>[6.2, 11] | 4.1<br>[3.3, 5.2] | 1.5<br>[1.2, 2] |
| Dickens | 42.5<br>[37.2, 49.3] | 12.4<br>[10.7, 14.1] | 24<br>[21.2, 27.6] | 6.3<br>[5.3, 7.4] | 6.9<br>[5.8, 8] | 3.2<br>[2.7, 3.8] | 2.8<br>[2.4, 3.3] | 0.9<br>[0.8, 1.1] |
| Dimmit | 30.9<br>[25.7, 37] | 10.9<br>[8.8, 13.4] | 17.7<br>[15, 20.9] | 2.2<br>[1.8, 2.6] | 5.2<br>[4.2, 6.2] | 2.8<br>[2.1, 3.4] | 2.1<br>[1.7, 2.5] | 0.3<br>[0.3, 0.4] |
| Donley | 118.5<br>[102.1, 138.4] | 22.9<br>[19.7, 25.8] | 83.2<br>[72, 96.5] | 12.5<br>[10.5, 15] | 16.4<br>[14.1, 19.3] | 5.8<br>[4.9, 6.7] | 9<br>[7.7, 10.8] | 1.7<br>[1.4, 2.1] |
| Duval | 34.9<br>[29.4, 42.4] | 12.9<br>[10.6, 16.4] | 19.1<br>[16.2, 22.9] | 3<br>[2.6, 3.7] | 5.9<br>[5, 7.2] | 3.2<br>[2.7, 4.1] | 2.2<br>[1.9, 2.7] | 0.4<br>[0.4, 0.5] |
| Eastland | 100.7<br>[81.7, 124] | 33.9<br>[27.2, 42.2] | 56.1<br>[45.5, 68.3] | 11<br>[8.8, 13.7] | 15.8<br>[12.5, 19.8] | 8.3<br>[6.5, 10.4] | 5.9<br>[4.8, 7.5] | 1.5<br>[1.2, 1.9] |
| Ector | 35.3<br>[28.8, 42.9] | 16.1<br>[12.8, 20.2] | 16.6<br>[13.6, 19.7] | 2.7<br>[2.2, 3.3] | 6.3<br>[5.1, 7.7] | 4<br>[3.2, 5] | 1.9<br>[1.6, 2.3] | 0.4<br>[0.3, 0.5] |
| Edwards | 28.7<br>[25.1, 32.6] | 10.2<br>[8.8, 11.8] | 14.4<br>[12.7, 16.4] | 4<br>[3.5, 4.7] | 5<br>[4.3, 5.8] | 2.7<br>[2.3, 3.2] | 1.7<br>[1.5, 2] | 0.6<br>[0.5, 0.7] |
| El Paso | 37<br>[30.2, 45.2] | 14<br>[10.9, 17.6] | 19.9<br>[16.3, 23.9] | 3.4<br>[2.7, 4.2] | 6.3<br>[5, 7.7] | 3.5<br>[2.7, 4.5] | 2.3<br>[1.9, 2.7] | 0.5<br>[0.4, 0.6] |
| Ellis | 47.5<br>[38.8, 58] | 19.4<br>[15.1, 24.7] | 23.1<br>[19.5, 27.7] | 4.9<br>[4.1, 6] | 8<br>[6.5, 10] | 4.8<br>[3.7, 6] | 2.6<br>[2.1, 3.1] | 0.7<br>[0.6, 0.9] |
| Erath | 235<br>[189, 301.6] | 53.9<br>[41.8, 68.6] | 162.7<br>[131.1, 209.9] | 19<br>[15.1, 24.9] | 31.1<br>[25, 40] | 12.6<br>[10, 16.7] | 15.9<br>[12.7, 20.8] | 2.5<br>[2, 3.2] |
| Falls | 32.4<br>[27.4, 39] | 13.2<br>[10.7, 16.2] | 15.2<br>[12.8, 17.7] | 4.1<br>[3.4, 4.9] | 5.7<br>[4.7, 6.9] | 3.3<br>[2.7, 4.1] | 1.8<br>[1.5, 2.1] | 0.6<br>[0.5, 0.7] |
| Fannin | 53.7<br>[45.9, 68.5] | 20.1<br>[16.7, 26.6] | 26.3<br>[22.4, 33] | 7.3<br>[6.1, 9.4] | 8.9<br>[7.6, 11.6] | 4.9<br>[4.1, 6.7] | 2.9<br>[2.5, 3.7] | 1<br>[0.8, 1.3] |
| Fayette | 32.5<br>[26.9, 40.1] | 12.2<br>[9.8, 15.5] | 16.2<br>[13.5, 20] | 4.1<br>[3.3, 5] | 5.6<br>[4.6, 7] | 3.1<br>[2.5, 4] | 1.9<br>[1.6, 2.4] | 0.6<br>[0.5, 0.7] |
| Fisher | 53.2<br>[45.3, 60.9] | 12.3<br>[10.4, 14.5] | 34.9<br>[29.8, 39.9] | 6<br>[5, 7] | 7.8<br>[6.7, 9.3] | 3.1<br>[2.6, 3.7] | 3.8<br>[3.3, 4.7] | 0.8<br>[0.7, 1] |
| Floyd | 29.5<br>[25.3, 36.4] | 10<br>[8.3, 12.2] | 16.6<br>[14.3, 20.3] | 3<br>[2.5, 3.7] | 4.9<br>[4.2, 6.1] | 2.5<br>[2.1, 3.2] | 1.9<br>[1.7, 2.4] | 0.4<br>[0.4, 0.5] |
| Foard | 27.8<br>[24.4, 31.7] | 7.6<br>[6.7, 8.5] | 15.7<br>[13.8, 17.8] | 4.6<br>[3.9, 5.3] | 4.6<br>[3.9, 5.3] | 2<br>[1.7, 2.3] | 1.9<br>[1.6, 2.2] | 0.7<br>[0.6, 0.8] |
| Fort Bend | 27.7<br>[22.3, 35.4] | 10.1<br>[7.8, 13.4] | 14.9<br>[12.3, 18.4] | 2.7<br>[2.2, 3.4] | 4.7<br>[3.7, 6] | 2.5<br>[2, 3.4] | 1.7<br>[1.5, 2.1] | 0.4<br>[0.3, 0.5] |
| Franklin | 38<br>[32.3, 46.1] | 14<br>[11.6, 17.4] | 19.5<br>[16.5, 23.6] | 4.6<br>[3.9, 5.6] | 6.4<br>[5.3, 7.9] | 3.5<br>[2.8, 4.4] | 2.2<br>[1.9, 2.7] | 0.7<br>[0.5, 0.8] |
| Freestone | 44<br>[36.3, 54.5] | 18.1<br>[14.4, 23.3] | 20.2<br>[17.1, 24.6] | 5.8<br>[4.8, 7.2] | 7.7<br>[6.2, 9.7] | 4.5<br>[3.5, 5.7] | 2.3<br>[1.9, 2.8] | 0.8<br>[0.7, 1] |
| Frio | 35.4<br>[29.5, 42.1] | 12.3<br>[9.7, 15] | 20.2<br>[17.2, 23.6] | 3<br>[2.5, 3.6] | 5.8<br>[4.8, 7] | 3.1<br>[2.4, 3.8] | 2.3<br>[1.9, 2.8] | 0.4<br>[0.4, 0.5] |
| Gaines | 375.3<br>[294.2, 501.9] | 199.4<br>[154.1, 259.6] | 136.7<br>[104.6, 179.1] | 39.5<br>[29.7, 52.9] | 66.3<br>[51.1, 87.9] | 47.8<br>[36.9, 63.7] | 13.5<br>[10.4, 17.9] | 5.1<br>[3.8, 6.9] |
| Galveston | 43<br>[36.4, 55] | 16.8<br>[13.5, 22.5] | 21.5<br>[18.4, 26.8] | 5<br>[4.2, 6.3] | 7.3<br>[6.1, 9.3] | 4.2<br>[3.4, 5.5] | 2.4<br>[2.1, 3] | 0.7<br>[0.6, 0.9] |
| Garza | 36.3<br>[30.3, 42.3] | 12<br>[9.7, 14.5] | 19.3<br>[16.3, 22.3] | 4.9<br>[4.1, 5.8] | 6<br>[4.9, 7.1] | 3.1<br>[2.5, 3.7] | 2.2<br>[1.8, 2.6] | 0.7<br>[0.6, 0.8] |
| Gillespie | 66.2<br>[53.2, 81] | 26<br>[20.3, 32.6] | 29.3<br>[23.8, 36.1] | 10.9<br>[8.8, 13.6] | 11.2<br>[8.7, 13.8] | 6.4<br>[4.9, 8] | 3.2<br>[2.6, 4] | 1.5<br>[1.2, 1.9] |
| Glasscock | 33.3<br>[29.7, 37.4] | 12.2<br>[10.7, 13.9] | 16.5<br>[14.7, 18.7] | 4.5<br>[3.9, 5.2] | 5.9<br>[5.2, 6.7] | 3.2<br>[2.8, 3.7] | 2<br>[1.7, 2.3] | 0.7<br>[0.6, 0.8] |
| Goliad | 34.1<br>[28.4, 40.5] | 12<br>[9.5, 14.3] | 17.1<br>[14.6, 20.1] | 5.1<br>[4.2, 6.2] | 5.8<br>[4.7, 6.9] | 3<br>[2.4, 3.7] | 2<br>[1.7, 2.4] | 0.7<br>[0.6, 0.9] |
| Gonzales | 35.9<br>[29.3, 44] | 14.5<br>[11.4, 18.2] | 18.1<br>[15.1, 22.2] | 3.2<br>[2.6, 3.9] | 6.2<br>[4.9, 7.6] | 3.6<br>[2.8, 4.7] | 2.1<br>[1.8, 2.5] | 0.5<br>[0.4, 0.6] |
| Gray | 36.4<br>[30.3, 45] | 14<br>[11.3, 17.9] | 18.5<br>[15.5, 22.1] | 3.9<br>[3.2, 4.8] | 6.2<br>[5, 7.7] | 3.5<br>[2.8, 4.5] | 2.1<br>[1.7, 2.6] | 0.6<br>[0.4, 0.7] |
| Grayson | 49.9<br>[40.9, 62.4] | 20.7<br>[16.2, 27.5] | 23.3<br>[19.5, 28.6] | 5.6<br>[4.7, 7.1] | 8.5<br>[6.6, 10.8] | 5.1<br>[3.9, 6.6] | 2.6<br>[2.2, 3.2] | 0.8<br>[0.6, 1] |
| Gregg | 45<br>[37.3, 54.4] | 18<br>[14.5, 22.5] | 22.4<br>[19.2, 27.1] | 4.4<br>[3.6, 5.3] | 7.6<br>[6.3, 9.4] | 4.5<br>[3.5, 5.6] | 2.5<br>[2.1, 3.1] | 0.6<br>[0.5, 0.8] |
| Grimes | 41.8<br>[34.7, 52.5] | 17.7<br>[13.9, 23.1] | 19.4<br>[16.3, 23.6] | 5<br>[4.1, 6.2] | 7.3<br>[5.9, 9.2] | 4.4<br>[3.5, 5.7] | 2.2<br>[1.8, 2.7] | 0.7<br>[0.6, 0.9] |
| Guadalupe | 38.2<br>[30.7, 45.4] | 14.2<br>[10.8, 17.7] | 19.9<br>[16.6, 23.3] | 4.1<br>[3.4, 4.9] | 6.4<br>[5.1, 7.8] | 3.6<br>[2.7, 4.4] | 2.3<br>[1.9, 2.7] | 0.6<br>[0.5, 0.7] |
| Hale | 37.3<br>[30, 44.5] | 13<br>[10.1, 15.8] | 21<br>[17.2, 25] | 3.2<br>[2.6, 3.8] | 6.1<br>[4.8, 7.3] | 3.3<br>[2.5, 4] | 2.4<br>[1.9, 2.9] | 0.4<br>[0.4, 0.5] |
| Hall | 59.4<br>[49.4, 69.8] | 18.3<br>[15.2, 21.1] | 31.5<br>[26.6, 37.5] | 9.6<br>[7.7, 11.5] | 9.7<br>[7.9, 11.3] | 4.7<br>[3.8, 5.6] | 3.6<br>[2.9, 4.2] | 1.4<br>[1, 1.6] |
| Hamilton | 28.4<br>[23.4, 34.3] | 10.7<br>[8.5, 13] | 13.9<br>[11.3, 16.4] | 3.8<br>[3.1, 4.6] | 4.9<br>[4, 6] | 2.7<br>[2.2, 3.4] | 1.6<br>[1.4, 1.9] | 0.5<br>[0.4, 0.7] |
| Hansford | 38.4<br>[31.3, 46] | 13.6<br>[10.8, 16.7] | 21.6<br>[17.9, 25.5] | 3.2<br>[2.6, 3.9] | 6.4<br>[5.1, 7.6] | 3.4<br>[2.7, 4.2] | 2.4<br>[2, 3] | 0.5<br>[0.4, 0.6] |
| Hardeman | 38.5<br>[33.1, 45] | 13.4<br>[11.3, 15.7] | 20.8<br>[17.9, 24.6] | 4.3<br>[3.6, 5.2] | 6.5<br>[5.4, 7.7] | 3.4<br>[2.8, 4.1] | 2.4<br>[2, 2.8] | 0.6<br>[0.5, 0.8] |
| Hardin | 41.6<br>[33.5, 50.2] | 18.1<br>[13.8, 22.4] | 18.9<br>[15.5, 22.5] | 4.9<br>[3.9, 5.9] | 7.4<br>[5.9, 9] | 4.5<br>[3.5, 5.6] | 2.2<br>[1.8, 2.6] | 0.7<br>[0.6, 0.9] |
| Harris | 38.2<br>[31.9, 47.9] | 15.6<br>[12.1, 19.8] | 18.9<br>[15.9, 23.2] | 3.7<br>[3.1, 4.6] | 6.6<br>[5.3, 8.4] | 3.9<br>[3.1, 5.1] | 2.2<br>[1.8, 2.6] | 0.5<br>[0.4, 0.7] |
| Harrison | 48.4<br>[39, 60.5] | 17.3<br>[13.1, 22.5] | 26.4<br>[21.5, 32.2] | 4.9<br>[3.9, 6.2] | 7.9<br>[6.2, 9.9] | 4.3<br>[3.2, 5.5] | 2.9<br>[2.4, 3.6] | 0.7<br>[0.5, 0.9] |
| Hartley | 49<br>[41.4, 58.1] | 20.8<br>[17, 25] | 21.3<br>[18.1, 25.2] | 6.9<br>[5.8, 8.3] | 8.8<br>[7.4, 10.7] | 5.3<br>[4.3, 6.5] | 2.4<br>[2.1, 2.9] | 1<br>[0.8, 1.2] |
| Haskell | 41.2<br>[35, 49.9] | 14.3<br>[11.7, 17.4] | 20.6<br>[17.5, 24.6] | 6.6<br>[5.6, 8.1] | 6.9<br>[5.8, 8.4] | 3.6<br>[2.9, 4.5] | 2.4<br>[2, 2.8] | 0.9<br>[0.8, 1.2] |
| Hays | 86<br>[72.6, 108.6] | 30.5<br>[24.2, 39] | 46.6<br>[39, 58.2] | 9.5<br>[7.8, 11.9] | 13.5<br>[11, 17.4] | 7.4<br>[5.9, 9.7] | 4.9<br>[4, 6.2] | 1.3<br>[1, 1.6] |
| Hemphill | 40.8<br>[34.4, 47.5] | 12.2<br>[10.4, 14.5] | 24.3<br>[20.7, 28.7] | 4.2<br>[3.5, 5] | 6.5<br>[5.5, 7.7] | 3.1<br>[2.6, 3.8] | 2.8<br>[2.3, 3.3] | 0.6<br>[0.5, 0.7] |
| Henderson | 50.6<br>[41.9, 61.5] | 20.4<br>[16.3, 25.7] | 23.3<br>[19.8, 28] | 6.5<br>[5.4, 7.9] | 8.6<br>[7, 10.6] | 5.1<br>[4, 6.6] | 2.6<br>[2.2, 3.2] | 0.9<br>[0.8, 1.1] |
| Hidalgo | 30.8<br>[25.8, 38.3] | 11.7<br>[9.6, 15.7] | 16.8<br>[14.1, 20.1] | 2.2<br>[1.8, 2.7] | 5.2<br>[4.3, 6.7] | 3<br>[2.3, 3.9] | 1.9<br>[1.6, 2.3] | 0.3<br>[0.3, 0.4] |
| Hill | 44.8<br>[35.6, 55.2] | 17.9<br>[14, 23.5] | 21.4<br>[17.6, 26] | 5.4<br>[4.3, 6.6] | 7.7<br>[6, 9.6] | 4.5<br>[3.4, 5.9] | 2.4<br>[2, 3] | 0.8<br>[0.6, 0.9] |
| Hockley | 43.7<br>[35.8, 52.4] | 16.7<br>[13.2, 20.6] | 22.9<br>[19.1, 27.1] | 4<br>[3.3, 4.8] | 7.4<br>[6, 8.7] | 4.1<br>[3.3, 5.2] | 2.6<br>[2.1, 3.1] | 0.6<br>[0.5, 0.7] |
| Hood | 58.5<br>[48.5, 74] | 24<br>[18.9, 31] | 25.5<br>[21.3, 31.9] | 9<br>[7.4, 11.4] | 10.1<br>[8.1, 12.7] | 5.9<br>[4.6, 7.5] | 2.8<br>[2.3, 3.5] | 1.3<br>[1, 1.6] |
| Hopkins | 36.8<br>[30.5, 45.6] | 15<br>[12.1, 18.9] | 17.9<br>[15, 21.6] | 4<br>[3.3, 4.9] | 6.4<br>[5.2, 8] | 3.8<br>[2.9, 4.8] | 2.1<br>[1.7, 2.5] | 0.6<br>[0.5, 0.7] |
| Houston | 46<br>[38.1, 56.6] | 19.4<br>[15.5, 24.2] | 20.3<br>[17, 24.4] | 6.5<br>[5.4, 8] | 8<br>[6.5, 10] | 4.8<br>[3.8, 6.1] | 2.3<br>[1.9, 2.8] | 0.9<br>[0.7, 1.2] |
| Howard | 40.5<br>[33.6, 50.2] | 15.1<br>[11.9, 19.4] | 21.1<br>[18, 26] | 4.3<br>[3.6, 5.3] | 6.8<br>[5.5, 8.5] | 3.8<br>[2.9, 4.9] | 2.4<br>[2, 3] | 0.6<br>[0.5, 0.8] |
| Hudspeth | 78.7<br>[68.6, 93] | 16.3<br>[14.1, 18.9] | 47.5<br>[42, 55.6] | 14.9<br>[12.7, 18.2] | 11.6<br>[9.9, 14] | 4.2<br>[3.5, 5] | 5.3<br>[4.6, 6.3] | 2.1<br>[1.8, 2.6] |
| Hunt | 49.3<br>[39, 59.7] | 19.2<br>[14.9, 24.1] | 25<br>[20.3, 30] | 5<br>[4, 6] | 8.2<br>[6.5, 10.3] | 4.7<br>[3.6, 6] | 2.8<br>[2.3, 3.4] | 0.7<br>[0.6, 0.8] |
| Hutchinson | 44.3<br>[37.1, 53.1] | 16.5<br>[13.5, 20.4] | 22.8<br>[19.4, 27.4] | 4.9<br>[4.1, 5.9] | 7.3<br>[6.2, 9.2] | 4.1<br>[3.3, 5.1] | 2.6<br>[2.2, 3.2] | 0.7<br>[0.6, 0.9] |
| Irion | 28.2<br>[24.2, 32.4] | 10.8<br>[9.1, 12.5] | 13.3<br>[11.5, 15.3] | 4.1<br>[3.4, 4.8] | 5.1<br>[4.2, 6] | 2.8<br>[2.4, 3.4] | 1.6<br>[1.4, 1.9] | 0.6<br>[0.5, 0.7] |
| Jack | 35.9<br>[29.5, 43.4] | 12.7<br>[10.1, 15.8] | 18.5<br>[15.5, 22] | 4.6<br>[3.8, 5.6] | 6<br>[4.9, 7.4] | 3.2<br>[2.5, 4.1] | 2.1<br>[1.8, 2.5] | 0.7<br>[0.5, 0.8] |
| Jackson | 33<br>[27.4, 40.6] | 14.1<br>[11.2, 17.5] | 16<br>[13.5, 19.4] | 3.1<br>[2.5, 3.8] | 5.9<br>[4.8, 7.3] | 3.6<br>[2.7, 4.5] | 1.9<br>[1.5, 2.3] | 0.4<br>[0.4, 0.5] |
| Jasper | 60.1<br>[48.7, 73.8] | 25.3<br>[19.8, 32.3] | 27<br>[22.2, 32.6] | 7.8<br>[6.4, 9.6] | 10.4<br>[8.2, 13] | 6.3<br>[4.8, 8.1] | 3<br>[2.5, 3.7] | 1.1<br>[0.9, 1.4] |
| Jeff Davis | 38.1<br>[33.7, 41.8] | 8.1<br>[7.3, 8.7] | 8.8<br>[8, 9.5] | 21.2<br>[18.4, 23.6] | 6.6<br>[5.8, 7.4] | 2.2<br>[2, 2.4] | 1.2<br>[1, 1.3] | 3.2<br>[2.7, 3.7] |
| Jefferson | 33.8<br>[28.2, 41] | 14.1<br>[11.4, 17.3] | 16.2<br>[13.7, 19.5] | 3.5<br>[2.9, 4.3] | 5.9<br>[4.9, 7.3] | 3.5<br>[2.9, 4.5] | 1.9<br>[1.6, 2.3] | 0.5<br>[0.4, 0.6] |
| Jim Hogg | 39.4<br>[32.6, 46.9] | 13.4<br>[10.7, 16.1] | 22.7<br>[18.8, 26.2] | 3.5<br>[2.9, 4.2] | 6.5<br>[5.2, 7.5] | 3.4<br>[2.7, 4.1] | 2.6<br>[2.1, 3] | 0.5<br>[0.4, 0.6] |
| Jim Wells | 30.3<br>[25, 36.8] | 11.6<br>[9.5, 14.5] | 16<br>[13.8, 19] | 2.7<br>[2.3, 3.3] | 5.2<br>[4.3, 6.3] | 2.9<br>[2.4, 3.7] | 1.9<br>[1.6, 2.2] | 0.4<br>[0.3, 0.5] |
| Johnson | 51.9<br>[41.3, 64.9] | 21.8<br>[16.8, 27.6] | 24.3<br>[19.9, 30.1] | 5.7<br>[4.6, 7.1] | 8.8<br>[7.2, 11.2] | 5.3<br>[4.2, 6.8] | 2.7<br>[2.2, 3.3] | 0.8<br>[0.6, 1] |
| Jones | 39.7<br>[33.4, 49.2] | 12.7<br>[10.3, 16.1] | 21.1<br>[18, 25.8] | 5.9<br>[5, 7.3] | 6.4<br>[5.3, 8.1] | 3.2<br>[2.5, 4.1] | 2.4<br>[2, 3] | 0.8<br>[0.7, 1.1] |
| Karnes | 38.2<br>[31.4, 45] | 13.1<br>[10.4, 15.8] | 20.4<br>[17.2, 23.5] | 4.6<br>[3.8, 5.5] | 6.3<br>[5.1, 7.6] | 3.3<br>[2.6, 4.1] | 2.3<br>[1.9, 2.7] | 0.7<br>[0.5, 0.8] |
| Kaufman | 37.5<br>[30.3, 47.2] | 16.2<br>[12.7, 21.1] | 17.7<br>[14.7, 21.3] | 3.4<br>[2.8, 4.2] | 6.6<br>[5.2, 8.2] | 4<br>[3.1, 5.2] | 2<br>[1.6, 2.5] | 0.5<br>[0.4, 0.6] |
| Kendall | 69.6<br>[56.5, 85.2] | 25.5<br>[20.4, 31.8] | 33.9<br>[27.4, 41.4] | 10.1<br>[8.1, 12.5] | 11.3<br>[9.1, 13.9] | 6.2<br>[4.9, 7.9] | 3.7<br>[2.9, 4.5] | 1.4<br>[1.1, 1.8] |
| Kenedy | 17.5<br>[16.4, 18.5] | 3.3<br>[3.1, 3.5] | 10.8<br>[10.2, 11.3] | 3.4<br>[3.1, 3.7] | 2.9<br>[2.6, 3.1] | 0.9<br>[0.8, 1] | 1.4<br>[1.3, 1.5] | 0.5<br>[0.5, 0.6] |
| Kent | 58.3<br>[52.7, 63.4] | 13.8<br>[12.4, 15.1] | 34.3<br>[31.4, 37.1] | 10.2<br>[8.9, 11.7] | 9.2<br>[8.2, 10.3] | 3.7<br>[3.3, 4.1] | 4.1<br>[3.6, 4.6] | 1.5<br>[1.3, 1.8] |
| Kerr | 73.5<br>[60.9, 92.3] | 26.5<br>[21.7, 34] | 36.2<br>[29.7, 44.7] | 11.3<br>[9.1, 14.2] | 12<br>[9.7, 15.1] | 6.5<br>[5.2, 8.3] | 3.9<br>[3.2, 4.8] | 1.6<br>[1.2, 2] |
| Kimble | 35.6<br>[30.1, 42.1] | 13.5<br>[11, 16.2] | 16.8<br>[14.4, 19.7] | 5.3<br>[4.4, 6.3] | 6.2<br>[5.2, 7.5] | 3.5<br>[2.8, 4.2] | 2<br>[1.7, 2.3] | 0.8<br>[0.6, 0.9] |
| King | 12.9<br>[11.9, 13.8] | 3.1<br>[2.8, 3.4] | 6.3<br>[5.9, 6.6] | 3.5<br>[3.2, 3.8] | 2.3<br>[2, 2.5] | 0.9<br>[0.8, 0.9] | 0.9<br>[0.8, 0.9] | 0.5<br>[0.5, 0.6] |
| Kinney | 48.2<br>[42.3, 56] | 12.8<br>[11.1, 15] | 28.6<br>[25.3, 33] | 6.8<br>[5.9, 8] | 7.5<br>[6.4, 8.9] | 3.3<br>[2.7, 3.9] | 3.2<br>[2.8, 3.9] | 1<br>[0.8, 1.2] |
| Kleberg | 82.5<br>[67.4, 103.7] | 24.5<br>[18.9, 31.6] | 52<br>[44.1, 64.2] | 6.3<br>[5.2, 8] | 12.3<br>[9.9, 15.4] | 5.9<br>[4.5, 7.5] | 5.5<br>[4.6, 6.9] | 0.9<br>[0.7, 1.1] |
| Knox | 20.6<br>[17.4, 24.2] | 7.7<br>[6.3, 9.4] | 10.8<br>[9.2, 12.5] | 2<br>[1.7, 2.4] | 3.6<br>[3, 4.3] | 2<br>[1.6, 2.5] | 1.3<br>[1.1, 1.5] | 0.3<br>[0.2, 0.4] |
| La Salle | 73.9<br>[59.2, 87.7] | 19.4<br>[16, 23.4] | 46.9<br>[37.9, 55.4] | 7.8<br>[6.1, 9.4] | 11<br>[8.8, 13.1] | 4.9<br>[4, 5.9] | 5.1<br>[4.1, 6.2] | 1.1<br>[0.9, 1.3] |
| Lamar | 36.2<br>[29.8, 43.3] | 15.4<br>[12.4, 19] | 17<br>[14.3, 19.9] | 3.7<br>[3.1, 4.5] | 6.3<br>[5.2, 7.7] | 3.9<br>[3.1, 4.8] | 2<br>[1.6, 2.3] | 0.5<br>[0.4, 0.6] |
| Lamb | 28.8<br>[25, 35.4] | 10.4<br>[8.6, 13] | 15.9<br>[14, 19.3] | 2.5<br>[2.2, 3.1] | 4.9<br>[4.1, 6] | 2.7<br>[2.2, 3.4] | 1.9<br>[1.6, 2.3] | 0.4<br>[0.3, 0.5] |
| Lampasas | 60.3<br>[48.5, 73.7] | 22.8<br>[18, 28.7] | 28.4<br>[22.8, 33.7] | 9<br>[7.1, 11] | 10.1<br>[7.9, 12.4] | 5.6<br>[4.3, 7.2] | 3.1<br>[2.5, 3.7] | 1.3<br>[1, 1.5] |
| Lavaca | 43.4<br>[35.7, 53.7] | 17.3<br>[13.6, 22.2] | 20.9<br>[17.6, 25.7] | 4.9<br>[4.1, 6.1] | 7.4<br>[6, 9.4] | 4.3<br>[3.4, 5.7] | 2.4<br>[2, 2.9] | 0.7<br>[0.6, 0.9] |
| Lee | 50.8<br>[41.5, 62] | 18.5<br>[14.6, 23.1] | 26.2<br>[21.9, 32.1] | 6<br>[4.9, 7.4] | 8.4<br>[6.8, 10.4] | 4.6<br>[3.6, 5.9] | 2.9<br>[2.5, 3.5] | 0.8<br>[0.7, 1] |
| Leon | 31.9<br>[26.4, 39] | 12.6<br>[10.1, 15.9] | 15.6<br>[13.3, 18.5] | 3.8<br>[3.1, 4.6] | 5.5<br>[4.5, 6.9] | 3.2<br>[2.5, 4.1] | 1.8<br>[1.5, 2.2] | 0.5<br>[0.4, 0.7] |
| Liberty | 28.2<br>[23.4, 35] | 11.8<br>[9.5, 14.9] | 14.1<br>[11.6, 16.8] | 2.3<br>[1.9, 2.8] | 5<br>[4, 6.3] | 3<br>[2.3, 3.9] | 1.7<br>[1.4, 2] | 0.3<br>[0.3, 0.4] |
| Limestone | 32.6<br>[26.4, 39.4] | 12.6<br>[9.9, 16.3] | 16.3<br>[13.5, 19.2] | 3.7<br>[3, 4.5] | 5.6<br>[4.5, 6.8] | 3.2<br>[2.4, 4] | 1.9<br>[1.6, 2.3] | 0.5<br>[0.4, 0.7] |
| Lipscomb | 39.8<br>[33.4, 47.6] | 13.5<br>[11.2, 16.2] | 21.7<br>[18.1, 25.7] | 4.5<br>[3.8, 5.5] | 6.6<br>[5.4, 8] | 3.5<br>[2.8, 4.3] | 2.5<br>[2.1, 3] | 0.7<br>[0.5, 0.8] |
| Live Oak | 29.2<br>[24.3, 35] | 10.7<br>[8.7, 13.4] | 14.2<br>[12.1, 16.7] | 4.1<br>[3.5, 5] | 5<br>[4.2, 6.1] | 2.8<br>[2.2, 3.4] | 1.7<br>[1.4, 2] | 0.6<br>[0.5, 0.7] |
| Llano | 55.4<br>[44.4, 68] | 22.1<br>[17, 27.7] | 23<br>[19, 28] | 10.4<br>[8.4, 12.9] | 9.5<br>[7.6, 11.7] | 5.4<br>[4.2, 6.9] | 2.6<br>[2.1, 3.1] | 1.5<br>[1.2, 1.9] |
| Lubbock | 84<br>[68.4, 102.6] | 28.3<br>[21.8, 35.4] | 47.8<br>[39.6, 57.6] | 7.8<br>[6.4, 9.5] | 13.1<br>[10.5, 15.9] | 6.8<br>[5.4, 8.5] | 5<br>[4.1, 6.2] | 1.1<br>[0.9, 1.3] |
| Lynn | 35.2<br>[29.5, 41.8] | 14.3<br>[11.6, 17.5] | 17.1<br>[14.7, 19.7] | 3.9<br>[3.2, 4.5] | 6.2<br>[5.1, 7.4] | 3.6<br>[2.9, 4.5] | 2<br>[1.7, 2.3] | 0.6<br>[0.5, 0.7] |
| Madison | 38.6<br>[31.2, 46.3] | 15.3<br>[12, 18.9] | 19.4<br>[16.1, 23] | 3.9<br>[3.2, 4.7] | 6.6<br>[5.3, 8.2] | 3.9<br>[3, 4.8] | 2.2<br>[1.8, 2.7] | 0.6<br>[0.4, 0.7] |
| Marion | 30.5<br>[24.1, 36] | 10.1<br>[7.9, 12.7] | 15.7<br>[12.7, 18.4] | 4.4<br>[3.5, 5.2] | 5.1<br>[4, 6.1] | 2.6<br>[2, 3.2] | 1.8<br>[1.5, 2.2] | 0.6<br>[0.5, 0.8] |
| Martin | 33.4<br>[28.4, 40.9] | 12.8<br>[10.7, 16.7] | 17.4<br>[14.7, 20.9] | 3.2<br>[2.7, 3.9] | 5.7<br>[4.9, 7.1] | 3.3<br>[2.7, 4.2] | 2<br>[1.7, 2.5] | 0.5<br>[0.4, 0.6] |
| Mason | 55.3<br>[45.9, 65.3] | 16.6<br>[13.8, 19.8] | 28.9<br>[24, 33.9] | 9.8<br>[7.9, 11.7] | 8.9<br>[7.2, 10.8] | 4.2<br>[3.5, 5.2] | 3.2<br>[2.6, 3.9] | 1.4<br>[1.1, 1.7] |
| Matagorda | 32.7<br>[27.3, 40] | 14.2<br>[11.4, 17.7] | 15.5<br>[13.2, 18.7] | 3.1<br>[2.6, 3.8] | 5.8<br>[4.8, 7.3] | 3.6<br>[2.8, 4.6] | 1.8<br>[1.5, 2.2] | 0.4<br>[0.4, 0.6] |
| Maverick | 29.8<br>[24.9, 36.3] | 12.2<br>[9.7, 15] | 15.5<br>[13.1, 18.7] | 2<br>[1.7, 2.4] | 5.2<br>[4.2, 6.4] | 3.1<br>[2.4, 3.8] | 1.8<br>[1.5, 2.3] | 0.3<br>[0.2, 0.4] |
| McCulloch | 39.4<br>[31.7, 46.3] | 14.3<br>[11.4, 17.3] | 19.4<br>[15.9, 22.8] | 5.8<br>[4.6, 6.8] | 6.7<br>[5.3, 8] | 3.6<br>[2.9, 4.4] | 2.2<br>[1.8, 2.6] | 0.8<br>[0.7, 1] |
| McLennan | 71.6<br>[59.6, 89.9] | 24.4<br>[19.8, 31.6] | 40.9<br>[34.1, 49.9] | 6.3<br>[5.3, 7.9] | 11.2<br>[9.1, 14.1] | 5.9<br>[4.7, 7.8] | 4.4<br>[3.6, 5.5] | 0.9<br>[0.7, 1.1] |
| McMullen | 41.4<br>[38, 44.3] | 7.8<br>[7, 8.6] | 27.1<br>[25.2, 28.9] | 6.4<br>[5.7, 7.1] | 6.3<br>[5.6, 7.1] | 2.1<br>[1.8, 2.4] | 3.3<br>[2.9, 3.7] | 1<br>[0.8, 1.1] |
| Medina | 41.9<br>[32.5, 51.9] | 15<br>[11.4, 19.2] | 22.5<br>[17.7, 27.4] | 4.5<br>[3.5, 5.6] | 6.9<br>[5.2, 8.6] | 3.8<br>[2.7, 4.8] | 2.5<br>[2, 3.1] | 0.6<br>[0.5, 0.8] |
| Menard | 24.3<br>[21.6, 28.1] | 7.2<br>[6.2, 8.3] | 13.5<br>[12, 15.4] | 3.6<br>[3.2, 4.3] | 4<br>[3.6, 4.7] | 1.9<br>[1.6, 2.2] | 1.6<br>[1.4, 1.9] | 0.5<br>[0.5, 0.6] |
| Midland | 32.7<br>[26.8, 39.5] | 15.7<br>[12.4, 19.7] | 14.4<br>[12, 16.9] | 2.8<br>[2.3, 3.4] | 6<br>[4.9, 7.3] | 3.9<br>[3.2, 4.9] | 1.7<br>[1.4, 2] | 0.4<br>[0.3, 0.5] |
| Milam | 40.2<br>[33, 47.9] | 15.3<br>[12.2, 18.6] | 20.1<br>[17, 23.7] | 4.6<br>[3.8, 5.5] | 6.8<br>[5.4, 8.2] | 3.8<br>[2.9, 4.7] | 2.3<br>[1.9, 2.7] | 0.7<br>[0.5, 0.8] |
| Mills | 34.1<br>[29.5, 40.2] | 11.3<br>[9.6, 13.5] | 17.7<br>[15.3, 20.6] | 5.1<br>[4.4, 6] | 5.7<br>[4.9, 6.9] | 2.9<br>[2.5, 3.6] | 2<br>[1.8, 2.4] | 0.7<br>[0.6, 0.9] |
| Mitchell | 34.9<br>[29.1, 41.4] | 11<br>[9, 13.8] | 19<br>[16.3, 22.7] | 4.5<br>[3.8, 5.5] | 5.6<br>[4.7, 6.8] | 2.8<br>[2.3, 3.5] | 2.2<br>[1.9, 2.6] | 0.6<br>[0.5, 0.8] |
| Montague | 74.6<br>[60.5, 95.5] | 30.8<br>[24.6, 40] | 32.2<br>[26.4, 40.8] | 11.4<br>[9.2, 14.8] | 12.8<br>[10.3, 15.9] | 7.6<br>[5.9, 9.7] | 3.5<br>[2.9, 4.4] | 1.6<br>[1.3, 2] |
| Montgomery | 46<br>[38.7, 57.2] | 19.4<br>[15.8, 24.7] | 21.7<br>[18.4, 26.3] | 5.1<br>[4.3, 6.3] | 8<br>[6.5, 9.9] | 4.8<br>[3.9, 6.1] | 2.5<br>[2.1, 3] | 0.7<br>[0.6, 0.9] |
| Moore | 29.4<br>[24.7, 36] | 13.3<br>[10.8, 16.6] | 14.1<br>[12.1, 16.9] | 2<br>[1.7, 2.4] | 5.4<br>[4.4, 6.7] | 3.4<br>[2.6, 4.2] | 1.7<br>[1.4, 2] | 0.3<br>[0.2, 0.4] |
| Morris | 28.1<br>[22.4, 33.4] | 10<br>[7.7, 12.3] | 14.9<br>[12.3, 17.4] | 3.2<br>[2.6, 3.8] | 4.8<br>[3.7, 5.7] | 2.5<br>[1.9, 3.1] | 1.8<br>[1.4, 2.1] | 0.5<br>[0.4, 0.6] |
| Motley | 46.6<br>[41.8, 50.9] | 12.6<br>[11.2, 13.9] | 25.3<br>[22.9, 27.5] | 8.7<br>[7.6, 9.7] | 7.7<br>[6.7, 8.6] | 3.3<br>[2.9, 3.7] | 3<br>[2.7, 3.4] | 1.3<br>[1.1, 1.5] |
| Nacogdoches | 111.9<br>[89.8, 141.7] | 32.7<br>[25.6, 42.3] | 70.4<br>[56.4, 88.3] | 8.7<br>[6.9, 11.1] | 16.4<br>[13.1, 20.6] | 7.9<br>[6.2, 10.1] | 7.4<br>[5.8, 9.2] | 1.2<br>[0.9, 1.5] |
| Navarro | 38<br>[31.2, 47.6] | 14.1<br>[11.2, 18.3] | 20.5<br>[17.1, 24.8] | 3.3<br>[2.7, 4.1] | 6.3<br>[5.1, 8.1] | 3.5<br>[2.8, 4.6] | 2.3<br>[1.9, 2.8] | 0.5<br>[0.4, 0.6] |
| Newton | 55.1<br>[45, 65.9] | 21.7<br>[17.2, 26.4] | 24.4<br>[20.4, 28.7] | 9<br>[7.3, 10.9] | 9.4<br>[7.6, 11.4] | 5.4<br>[4.3, 6.7] | 2.8<br>[2.3, 3.3] | 1.3<br>[1, 1.5] |
| Nolan | 39<br>[32.7, 46.4] | 14.1<br>[11.6, 17.6] | 21.2<br>[17.7, 24.7] | 3.8<br>[3.2, 4.6] | 6.5<br>[5.3, 7.9] | 3.6<br>[2.8, 4.4] | 2.4<br>[2, 2.8] | 0.5<br>[0.4, 0.7] |
| Nueces | 40.2<br>[33.4, 49] | 15.2<br>[12.4, 18.9] | 21<br>[17.7, 25] | 4<br>[3.4, 4.9] | 6.8<br>[5.5, 8.3] | 3.8<br>[3, 4.8] | 2.4<br>[2, 2.9] | 0.6<br>[0.5, 0.7] |
| Ochiltree | 39.5<br>[32.7, 45.9] | 14.7<br>[11.7, 17.8] | 21.4<br>[18.1, 25] | 3.1<br>[2.5, 3.6] | 6.6<br>[5.4, 8] | 3.7<br>[2.9, 4.6] | 2.4<br>[2, 2.9] | 0.4<br>[0.4, 0.5] |
| Oldham | 39<br>[33.7, 45.9] | 12.2<br>[10.7, 14.3] | 19.4<br>[16.5, 22.9] | 7.4<br>[6.2, 9] | 6.5<br>[5.6, 7.9] | 3.2<br>[2.7, 3.9] | 2.3<br>[1.9, 2.7] | 1.1<br>[0.9, 1.3] |
| Orange | 49.1<br>[40.2, 63.4] | 22<br>[17.3, 28.7] | 21.4<br>[17.9, 27.3] | 5.7<br>[4.7, 7.4] | 8.6<br>[7, 11.1] | 5.4<br>[4.2, 7.2] | 2.4<br>[2, 3.1] | 0.8<br>[0.7, 1.1] |
| Palo Pinto | 44.3<br>[34.6, 53.4] | 18.5<br>[14.2, 22.8] | 20.4<br>[16.3, 24.4] | 5.4<br>[4.2, 6.6] | 7.7<br>[6, 9.4] | 4.6<br>[3.5, 5.7] | 2.3<br>[1.9, 2.8] | 0.8<br>[0.6, 1] |
| Panola | 46.6<br>[38.2, 56.7] | 17.7<br>[14.2, 22] | 23.6<br>[19.8, 28.1] | 5.3<br>[4.4, 6.4] | 7.8<br>[6.3, 9.6] | 4.4<br>[3.5, 5.5] | 2.7<br>[2.2, 3.2] | 0.7<br>[0.6, 0.9] |
| Parker | 54.5<br>[44.2, 72.1] | 23.3<br>[18.4, 31.4] | 24.5<br>[20, 30.9] | 6.9<br>[5.6, 9] | 9.3<br>[7.5, 12.7] | 5.7<br>[4.5, 7.8] | 2.7<br>[2.2, 3.4] | 1<br>[0.7, 1.2] |
| Parmer | 35.2<br>[29.9, 42.9] | 14<br>[11.3, 17.2] | 18.3<br>[15.9, 22] | 3.1<br>[2.7, 3.8] | 6.1<br>[5.1, 7.4] | 3.5<br>[2.8, 4.4] | 2.1<br>[1.8, 2.5] | 0.4<br>[0.4, 0.6] |
| Pecos | 43.9<br>[35.8, 52.9] | 17.6<br>[14, 22] | 21.3<br>[17.6, 25.3] | 5<br>[4, 6] | 7.5<br>[6.2, 9.2] | 4.4<br>[3.6, 5.5] | 2.4<br>[2, 2.9] | 0.7<br>[0.6, 0.9] |
| Polk | 31.3<br>[25.1, 37.1] | 12.2<br>[9.7, 14.7] | 14.8<br>[12.1, 17.5] | 4.1<br>[3.3, 5] | 5.4<br>[4.3, 6.4] | 3.1<br>[2.4, 3.9] | 1.7<br>[1.4, 2.1] | 0.6<br>[0.5, 0.7] |
| Potter | 35.7<br>[29.7, 43.2] | 14.9<br>[11.9, 18.5] | 17.6<br>[14.9, 20.7] | 3.4<br>[2.9, 4.1] | 6.2<br>[5.1, 7.4] | 3.7<br>[3, 4.6] | 2<br>[1.7, 2.4] | 0.5<br>[0.4, 0.6] |
| Presidio | 30.4<br>[25.2, 36.2] | 11.7<br>[9.5, 14.2] | 16<br>[13.6, 19.1] | 2.7<br>[2.3, 3.3] | 5.2<br>[4.4, 6.3] | 3<br>[2.4, 3.6] | 1.9<br>[1.6, 2.2] | 0.4<br>[0.3, 0.5] |
| Rains | 100.6<br>[80.4, 123.4] | 33.9<br>[26.9, 41.4] | 51.6<br>[41.1, 62.9] | 15.5<br>[12.1, 19.3] | 16<br>[12.6, 19.8] | 8.4<br>[6.6, 10.4] | 5.5<br>[4.4, 7] | 2.2<br>[1.7, 2.7] |
| Randall | 63.9<br>[50.4, 83.6] | 24.7<br>[19.2, 33] | 32.3<br>[25.7, 41.5] | 7.1<br>[5.6, 9.2] | 10.6<br>[8.1, 13.7] | 6<br>[4.6, 7.9] | 3.5<br>[2.8, 4.4] | 1<br>[0.8, 1.3] |
| Reagan | 22.9<br>[19.7, 27.4] | 8.5<br>[7.1, 10.5] | 12.6<br>[10.8, 15] | 1.8<br>[1.5, 2.2] | 4<br>[3.4, 4.9] | 2.2<br>[1.8, 2.7] | 1.5<br>[1.3, 1.8] | 0.3<br>[0.2, 0.3] |
| Red River | 32<br>[26.9, 39.1] | 13.4<br>[11, 16.9] | 14.2<br>[12.1, 17.1] | 4.6<br>[3.8, 5.6] | 5.7<br>[4.7, 7.2] | 3.4<br>[2.7, 4.4] | 1.7<br>[1.4, 2.1] | 0.7<br>[0.5, 0.8] |
| Reeves | 29.8<br>[25.3, 36.7] | 12.3<br>[10, 15.8] | 14.8<br>[12.8, 18] | 2.6<br>[2.2, 3.2] | 5.3<br>[4.4, 6.6] | 3.2<br>[2.6, 4] | 1.8<br>[1.5, 2.1] | 0.4<br>[0.3, 0.5] |
| Refugio | 39.4<br>[33.4, 47.8] | 11.9<br>[9.9, 14.7] | 22.3<br>[19.1, 26.9] | 5.2<br>[4.4, 6.4] | 6.3<br>[5.2, 7.7] | 3<br>[2.4, 3.8] | 2.5<br>[2.2, 3] | 0.7<br>[0.6, 0.9] |
| Roberts | 16.3<br>[14.5, 18.3] | 5.9<br>[5.2, 6.7] | 8.7<br>[7.8, 9.8] | 1.7<br>[1.5, 2] | 2.9<br>[2.6, 3.4] | 1.6<br>[1.4, 1.8] | 1.1<br>[1, 1.3] | 0.3<br>[0.2, 0.3] |
| Robertson | 30.6<br>[25.5, 36] | 12.5<br>[10.1, 15.4] | 15<br>[12.5, 17.4] | 3.2<br>[2.6, 3.7] | 5.4<br>[4.5, 6.6] | 3.2<br>[2.5, 4] | 1.8<br>[1.5, 2] | 0.5<br>[0.4, 0.6] |
| Rockwall | 55.6<br>[44.7, 70.9] | 22.9<br>[17.3, 30.2] | 26.1<br>[21.3, 32.9] | 6.4<br>[5.1, 8.3] | 9.4<br>[7.4, 12.1] | 5.6<br>[4.2, 7.5] | 2.9<br>[2.3, 3.7] | 0.9<br>[0.7, 1.2] |
| Runnels | 34.2<br>[28.1, 41] | 13.4<br>[10.8, 16.5] | 16.8<br>[14.1, 19.9] | 3.8<br>[3.1, 4.6] | 5.9<br>[4.8, 7.3] | 3.4<br>[2.7, 4.3] | 2<br>[1.6, 2.4] | 0.6<br>[0.4, 0.7] |
| Rusk | 37<br>[30.9, 45.3] | 14<br>[11.1, 17.1] | 18.7<br>[15.5, 22.9] | 4.5<br>[3.7, 5.5] | 6.3<br>[5.1, 7.7] | 3.5<br>[2.8, 4.4] | 2.2<br>[1.8, 2.6] | 0.6<br>[0.5, 0.8] |
| Sabine | 48.1<br>[39.9, 59.8] | 19.9<br>[15.9, 24.7] | 20.8<br>[17.3, 25.5] | 7.7<br>[6.3, 9.6] | 8.4<br>[6.9, 10.9] | 5<br>[3.9, 6.4] | 2.4<br>[1.9, 3] | 1.1<br>[0.9, 1.4] |
| San Augustine | 37.1<br>[30.7, 44.3] | 15<br>[12.3, 18.6] | 16.4<br>[13.8, 19.8] | 5.6<br>[4.6, 6.8] | 6.5<br>[5.2, 8.2] | 3.8<br>[3, 4.8] | 1.9<br>[1.6, 2.3] | 0.8<br>[0.6, 1] |
| San Jacinto | 39.5<br>[33.2, 47.2] | 14.9<br>[12.3, 19] | 19.4<br>[16.3, 23] | 5.1<br>[4.2, 6.1] | 6.7<br>[5.6, 8.1] | 3.7<br>[3, 4.8] | 2.2<br>[1.9, 2.6] | 0.7<br>[0.6, 0.9] |
| San Patricio | 33.5<br>[27.9, 39.6] | 13.1<br>[10.5, 16.2] | 17.1<br>[14.5, 20.1] | 3.1<br>[2.6, 3.7] | 5.7<br>[4.7, 6.9] | 3.3<br>[2.6, 4.1] | 2<br>[1.6, 2.4] | 0.4<br>[0.4, 0.5] |
| San Saba | 38.5<br>[32.5, 46.5] | 12.9<br>[10.6, 16.3] | 19.8<br>[16.8, 23.8] | 5.8<br>[4.8, 7] | 6.4<br>[5.2, 7.8] | 3.3<br>[2.7, 4.1] | 2.3<br>[1.9, 2.8] | 0.8<br>[0.7, 1] |
| Schleicher | 46.8<br>[39.7, 54.9] | 13.3<br>[10.9, 15.7] | 29.6<br>[24.8, 34.4] | 4<br>[3.3, 4.8] | 7.3<br>[6, 8.7] | 3.4<br>[2.7, 4.1] | 3.3<br>[2.8, 4] | 0.6<br>[0.5, 0.7] |
| Scurry | 34.7<br>[28.7, 42.5] | 12.4<br>[9.9, 15.9] | 19.4<br>[16.4, 23.6] | 3<br>[2.5, 3.7] | 5.8<br>[4.7, 7] | 3.1<br>[2.4, 3.9] | 2.2<br>[1.9, 2.7] | 0.4<br>[0.4, 0.5] |
| Shackelford | 69.6<br>[59.6, 81.1] | 22.7<br>[19.4, 26.4] | 36<br>[30.8, 41.5] | 11<br>[9.2, 13.2] | 11.3<br>[9.7, 13.2] | 5.8<br>[5, 6.7] | 4<br>[3.4, 4.8] | 1.6<br>[1.3, 1.9] |
| Shelby | 35.7<br>[29.2, 43.4] | 15.1<br>[11.9, 19.3] | 17.3<br>[14.2, 21] | 3.4<br>[2.8, 4.2] | 6.3<br>[5.1, 7.9] | 3.8<br>[3.1, 5] | 2<br>[1.7, 2.4] | 0.5<br>[0.4, 0.6] |
| Sherman | 39.4<br>[33.7, 45.8] | 14.6<br>[12.5, 17.5] | 20.7<br>[17.7, 23.9] | 3.9<br>[3.3, 4.6] | 6.7<br>[5.7, 8] | 3.8<br>[3.1, 4.5] | 2.4<br>[2.1, 2.8] | 0.6<br>[0.5, 0.7] |
| Smith | 58.3<br>[46.3, 71.8] | 23.2<br>[17.4, 30.2] | 28.5<br>[23.1, 34.1] | 6.4<br>[5.1, 7.7] | 9.8<br>[7.7, 12.1] | 5.7<br>[4.3, 7.3] | 3.1<br>[2.5, 3.8] | 0.9<br>[0.7, 1.1] |
| Somervell | 51.5<br>[43, 63.1] | 19.6<br>[15.8, 24.4] | 24.2<br>[20.2, 29.1] | 7.7<br>[6.3, 9.4] | 8.7<br>[7.3, 10.9] | 4.9<br>[3.9, 6.2] | 2.7<br>[2.3, 3.3] | 1.1<br>[0.9, 1.3] |
| Starr | 25.1<br>[21.1, 31] | 10<br>[8.1, 13] | 13.4<br>[11.4, 16.2] | 1.6<br>[1.4, 2] | 4.4<br>[3.6, 5.4] | 2.5<br>[2, 3.3] | 1.6<br>[1.4, 1.9] | 0.2<br>[0.2, 0.3] |
| Stephens | 79.7<br>[67.2, 98.4] | 27<br>[22.3, 33.5] | 41.5<br>[34.7, 50.9] | 11.6<br>[9.6, 14.6] | 12.8<br>[10.6, 15.8] | 6.7<br>[5.4, 8.3] | 4.5<br>[3.7, 5.6] | 1.6<br>[1.3, 2.1] |
| Sterling | 53.6<br>[46.9, 60.3] | 22.1<br>[19.7, 24.8] | 25.6<br>[22.5, 28.6] | 5.9<br>[5, 6.8] | 9.6<br>[8.4, 11.1] | 5.8<br>[5.1, 6.8] | 3<br>[2.5, 3.5] | 0.9<br>[0.7, 1] |
| Sutton | 31.3<br>[26.6, 36.4] | 10.6<br>[9, 12.6] | 16.8<br>[14.5, 19.6] | 3.7<br>[3.2, 4.4] | 5.3<br>[4.5, 6.2] | 2.7<br>[2.3, 3.4] | 2<br>[1.7, 2.3] | 0.5<br>[0.4, 0.6] |
| Swisher | 38.9<br>[32.8, 45.5] | 13.4<br>[11.2, 16] | 21.4<br>[18.1, 24.6] | 4.2<br>[3.5, 4.9] | 6.4<br>[5.4, 7.7] | 3.3<br>[2.7, 4.2] | 2.4<br>[2, 2.9] | 0.6<br>[0.5, 0.7] |
| Tarrant | 51.9<br>[42.7, 63.6] | 21<br>[16.4, 27.2] | 25.3<br>[21.2, 30.4] | 5.5<br>[4.6, 6.8] | 8.7<br>[7.2, 10.8] | 5.2<br>[4.1, 6.6] | 2.8<br>[2.3, 3.4] | 0.8<br>[0.6, 1] |
| Taylor | 63.5<br>[51.2, 79.7] | 23.2<br>[18.3, 29.4] | 34.5<br>[28.2, 43.1] | 5.7<br>[4.5, 7.1] | 10.2<br>[8.2, 13] | 5.6<br>[4.4, 7.4] | 3.8<br>[3, 4.6] | 0.8<br>[0.6, 1] |
| Terrell | 14.9<br>[13.1, 16.9] | 3.4<br>[2.9, 3.9] | 9.4<br>[8.4, 10.6] | 2.1<br>[1.8, 2.4] | 2.4<br>[2.1, 2.8] | 0.9<br>[0.8, 1.1] | 1.2<br>[1.1, 1.4] | 0.3<br>[0.3, 0.4] |
| Terry | 38.4<br>[32.6, 46.5] | 16.2<br>[13.4, 20.5] | 18.7<br>[16.3, 22.5] | 3.4<br>[2.9, 4.1] | 6.7<br>[5.6, 8.3] | 4.1<br>[3.4, 5.2] | 2.2<br>[1.8, 2.6] | 0.5<br>[0.4, 0.6] |
| Throckmorton | 50.7<br>[44.1, 57.4] | 18.5<br>[15.9, 20.6] | 23.8<br>[20.5, 26.8] | 8.6<br>[7.2, 9.9] | 8.9<br>[7.5, 10] | 4.9<br>[4.2, 5.6] | 2.8<br>[2.3, 3.1] | 1.2<br>[1, 1.5] |
| Titus | 32.3<br>[26.8, 39.6] | 12.7<br>[10.4, 16] | 16.6<br>[14.1, 20.1] | 2.7<br>[2.3, 3.3] | 5.5<br>[4.6, 6.9] | 3.2<br>[2.6, 4.1] | 1.9<br>[1.7, 2.3] | 0.4<br>[0.3, 0.5] |
| Tom Green | 45.8<br>[38.3, 55.7] | 16.8<br>[13.7, 20.9] | 24.2<br>[20.8, 28.9] | 4.5<br>[3.7, 5.4] | 7.6<br>[6.4, 9.1] | 4.2<br>[3.4, 5.2] | 2.7<br>[2.3, 3.3] | 0.6<br>[0.5, 0.8] |
| Travis | 76.2<br>[60.9, 98.7] | 30.8<br>[23.7, 40.3] | 35.6<br>[28.7, 45.5] | 10.3<br>[8.2, 13.6] | 12.7<br>[10.1, 16.7] | 7.4<br>[5.8, 10] | 3.8<br>[3.1, 4.9] | 1.4<br>[1.1, 1.9] |
| Trinity | 44.1<br>[36.4, 52.6] | 16.5<br>[13, 20.1] | 21.2<br>[17.8, 25.5] | 6.3<br>[5.1, 7.6] | 7.5<br>[6.2, 9.2] | 4.2<br>[3.3, 5.1] | 2.4<br>[2, 2.9] | 0.9<br>[0.7, 1.1] |
| Tyler | 53.1<br>[44.2, 63.2] | 20<br>[16.4, 24.4] | 24.9<br>[20.6, 28.8] | 8.1<br>[6.7, 9.6] | 8.9<br>[7.2, 10.7] | 5<br>[3.9, 6.1] | 2.8<br>[2.3, 3.3] | 1.1<br>[0.9, 1.4] |
| Upshur | 49.5<br>[39.5, 58.8] | 20.4<br>[16.1, 24.8] | 23.1<br>[18.8, 27.3] | 6<br>[4.8, 7.3] | 8.4<br>[6.7, 10.2] | 5<br>[3.9, 6.2] | 2.6<br>[2.1, 3.1] | 0.8<br>[0.7, 1] |
| Upton | 32.4<br>[27.5, 38.2] | 9.1<br>[7.6, 10.8] | 20<br>[16.8, 23.7] | 3.2<br>[2.6, 3.8] | 5.1<br>[4.2, 6] | 2.3<br>[1.9, 2.8] | 2.3<br>[1.9, 2.7] | 0.5<br>[0.4, 0.6] |
| Uvalde | 46<br>[37.7, 55.4] | 18.5<br>[14.9, 23.2] | 23.6<br>[20.2, 27.9] | 3.8<br>[3.2, 4.6] | 7.8<br>[6.5, 9.6] | 4.6<br>[3.7, 5.8] | 2.7<br>[2.3, 3.2] | 0.5<br>[0.5, 0.7] |
| Val Verde | 34<br>[28, 40.3] | 14.1<br>[11.6, 17.5] | 17.1<br>[14.3, 20] | 2.7<br>[2.2, 3.1] | 6<br>[4.9, 7.3] | 3.6<br>[2.9, 4.6] | 2<br>[1.6, 2.4] | 0.4<br>[0.3, 0.5] |
| Van Zandt | 55.6<br>[44.4, 68.5] | 22.8<br>[17.3, 28.7] | 25.8<br>[21.2, 31.4] | 7.3<br>[5.9, 9] | 9.4<br>[7.4, 11.9] | 5.6<br>[4.2, 7.1] | 2.8<br>[2.3, 3.5] | 1<br>[0.8, 1.3] |
| Victoria | 34.6<br>[28.3, 41.3] | 13.9<br>[10.9, 17.1] | 17.4<br>[14.6, 20.2] | 3.3<br>[2.7, 3.9] | 6<br>[4.8, 7.3] | 3.5<br>[2.7, 4.3] | 2<br>[1.7, 2.4] | 0.5<br>[0.4, 0.6] |
| Walker | 246.6<br>[196.6, 310.5] | 61.4<br>[47.2, 82.2] | 152.8<br>[118.8, 190.4] | 32.4<br>[25.3, 41.1] | 33.8<br>[26.3, 43.4] | 14.3<br>[10.6, 19.6] | 15.3<br>[11.9, 19.2] | 4.2<br>[3.3, 5.3] |
| Waller | 178.2<br>[140.8, 222.8] | 41.7<br>[30.9, 55.1] | 126.1<br>[101.6, 154.7] | 11.5<br>[9.1, 14.4] | 23.8<br>[18.6, 30.3] | 9.8<br>[7.2, 13] | 12.6<br>[10, 15.6] | 1.5<br>[1.2, 1.9] |
| Ward | 34.7<br>[28.5, 42.4] | 14.6<br>[11.8, 18.7] | 17<br>[14.1, 20.2] | 3.1<br>[2.5, 3.7] | 6.1<br>[5, 7.5] | 3.7<br>[3, 4.8] | 2<br>[1.6, 2.4] | 0.4<br>[0.4, 0.5] |
| Washington | 114.7<br>[90.9, 139.4] | 29.7<br>[23.4, 37] | 74.4<br>[59, 91.3] | 10.5<br>[8.2, 12.9] | 16.3<br>[12.8, 20.2] | 7.1<br>[5.4, 8.9] | 7.6<br>[6.2, 9.5] | 1.4<br>[1.1, 1.8] |
| Webb | 28.3<br>[23.6, 33.8] | 10.9<br>[8.7, 13] | 15.5<br>[13.2, 18.5] | 1.9<br>[1.6, 2.3] | 4.8<br>[4, 5.7] | 2.7<br>[2.2, 3.4] | 1.8<br>[1.5, 2.1] | 0.3<br>[0.2, 0.3] |
| Wharton | 34.9<br>[28.7, 42.6] | 13.6<br>[11.1, 17.1] | 18.1<br>[15.1, 22.2] | 3.1<br>[2.5, 3.8] | 5.9<br>[4.8, 7.5] | 3.4<br>[2.7, 4.4] | 2.1<br>[1.7, 2.6] | 0.4<br>[0.4, 0.6] |
| Wheeler | 35.9<br>[29.4, 44.8] | 12.9<br>[10.4, 16.3] | 18.7<br>[15.6, 22.9] | 4.3<br>[3.5, 5.4] | 6.1<br>[4.9, 7.7] | 3.3<br>[2.6, 4.2] | 2.2<br>[1.8, 2.6] | 0.6<br>[0.5, 0.8] |
| Wichita | 68.1<br>[57, 82.9] | 23.3<br>[18.7, 28.9] | 38.8<br>[33, 47.1] | 6.2<br>[5.2, 7.6] | 10.7<br>[8.7, 13.2] | 5.6<br>[4.5, 7] | 4.2<br>[3.5, 5.2] | 0.9<br>[0.7, 1.1] |
| Wilbarger | 47.8<br>[40.2, 56.7] | 16.8<br>[13.8, 20.9] | 26.1<br>[22.1, 30.8] | 4.9<br>[4.1, 5.9] | 7.9<br>[6.4, 9.5] | 4.2<br>[3.4, 5.3] | 2.9<br>[2.4, 3.5] | 0.7<br>[0.6, 0.9] |
| Willacy | 38.4<br>[31.6, 46.5] | 12.3<br>[9.7, 15.3] | 22.7<br>[18.8, 27.6] | 3.4<br>[2.7, 4.1] | 6.1<br>[4.9, 7.5] | 3.1<br>[2.4, 4] | 2.6<br>[2.1, 3.1] | 0.5<br>[0.4, 0.6] |
| Williamson | 55.1<br>[43.8, 67.8] | 23.9<br>[18.2, 30.4] | 24.4<br>[19.7, 29.2] | 6.9<br>[5.5, 8.4] | 9.6<br>[7.5, 11.9] | 5.9<br>[4.4, 7.5] | 2.7<br>[2.2, 3.2] | 1<br>[0.8, 1.2] |
| Wilson | 32.6<br>[26, 40.9] | 12.3<br>[9.4, 16.2] | 16.7<br>[13.4, 20] | 3.7<br>[2.9, 4.6] | 5.6<br>[4.3, 7] | 3.1<br>[2.4, 4] | 1.9<br>[1.6, 2.3] | 0.5<br>[0.4, 0.6] |
| Winkler | 31.2<br>[25.7, 37.5] | 12.5<br>[10.1, 15.3] | 16.2<br>[13.7, 19.2] | 2.4<br>[2, 3] | 5.4<br>[4.5, 6.7] | 3.2<br>[2.5, 4] | 1.9<br>[1.6, 2.3] | 0.4<br>[0.3, 0.4] |
| Wise | 60.5<br>[48.6, 77] | 26.1<br>[20.1, 34.4] | 27.2<br>[22.1, 33.5] | 7.5<br>[6, 9.4] | 10.5<br>[8.4, 13.4] | 6.4<br>[4.9, 8.6] | 3<br>[2.4, 3.7] | 1.1<br>[0.8, 1.3] |
| Wood | 56.6<br>[46.6, 69.3] | 19.7<br>[15.3, 24.3] | 28.8<br>[23.4, 34.7] | 8.3<br>[6.7, 10.2] | 9.2<br>[7.3, 11.3] | 4.9<br>[3.8, 6.1] | 3.2<br>[2.5, 3.8] | 1.2<br>[1, 1.4] |
| Yoakum | 41.4<br>[34.4, 51] | 17.3<br>[13.6, 22.1] | 21.2<br>[17.9, 25.9] | 3<br>[2.5, 3.7] | 7.2<br>[5.8, 9] | 4.4<br>[3.4, 5.6] | 2.4<br>[2, 3] | 0.4<br>[0.3, 0.6] |
| Young | 37.7<br>[31.2, 45.2] | 14.8<br>[11.9, 18.5] | 18.8<br>[15.8, 21.8] | 4.3<br>[3.5, 5] | 6.4<br>[5.3, 7.8] | 3.7<br>[2.9, 4.6] | 2.2<br>[1.8, 2.6] | 0.6<br>[0.5, 0.7] |
| Zapata | 24.2<br>[20.7, 29] | 8.5<br>[7, 10.7] | 14.1<br>[12.2, 16.9] | 1.6<br>[1.4, 2] | 4<br>[3.4, 5] | 2.1<br>[1.8, 2.7] | 1.7<br>[1.4, 2] | 0.2<br>[0.2, 0.3] |
| Zavala | 27.4<br>[22.8, 32.1] | 10.3<br>[8.1, 12.3] | 15.4<br>[12.8, 17.8] | 1.9<br>[1.6, 2.3] | 4.7<br>[3.8, 5.6] | 2.6<br>[2.1, 3.1] | 1.8<br>[1.5, 2.1] | 0.3<br>[0.2, 0.3] |

**Table S.5.**
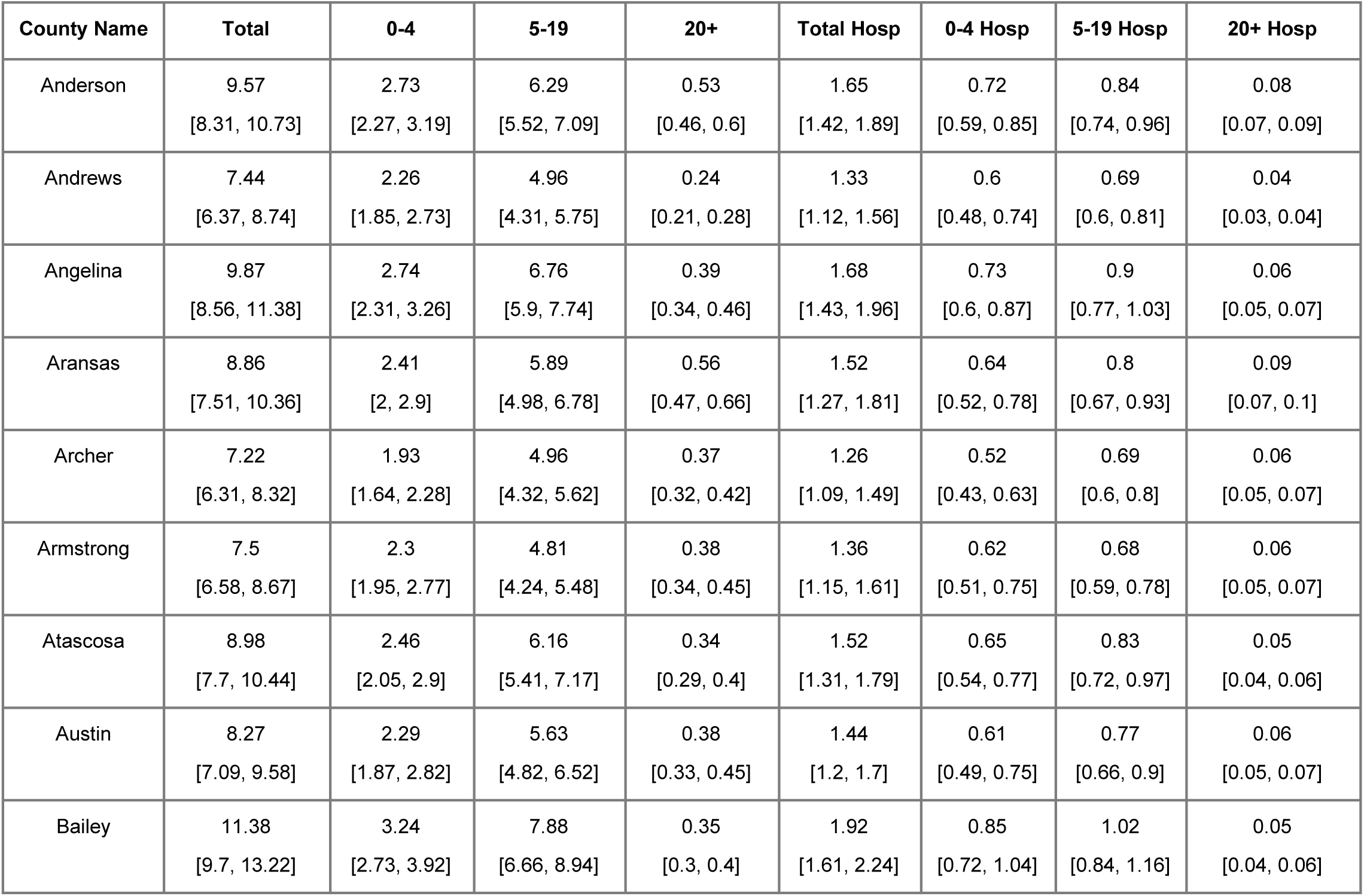
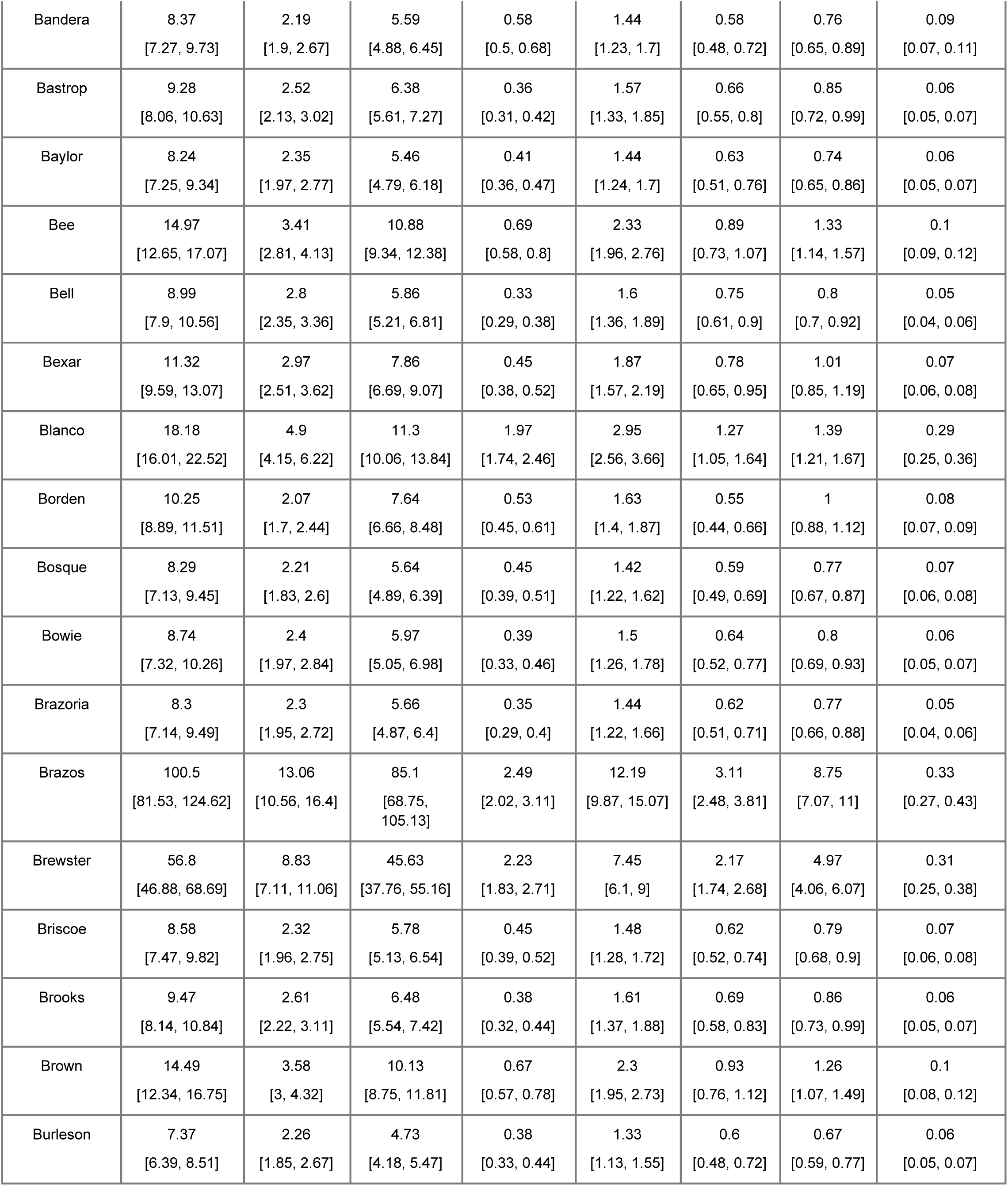
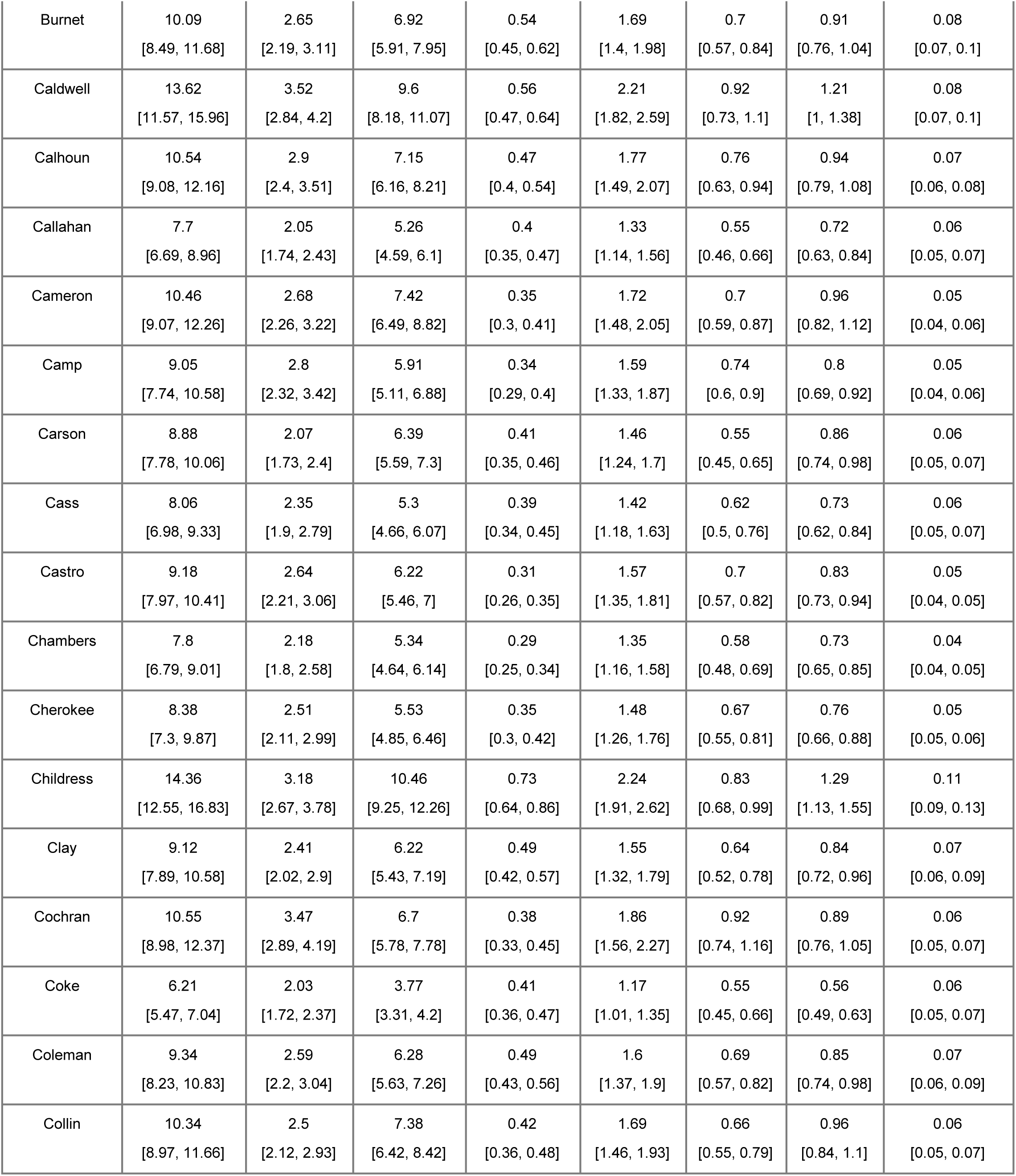
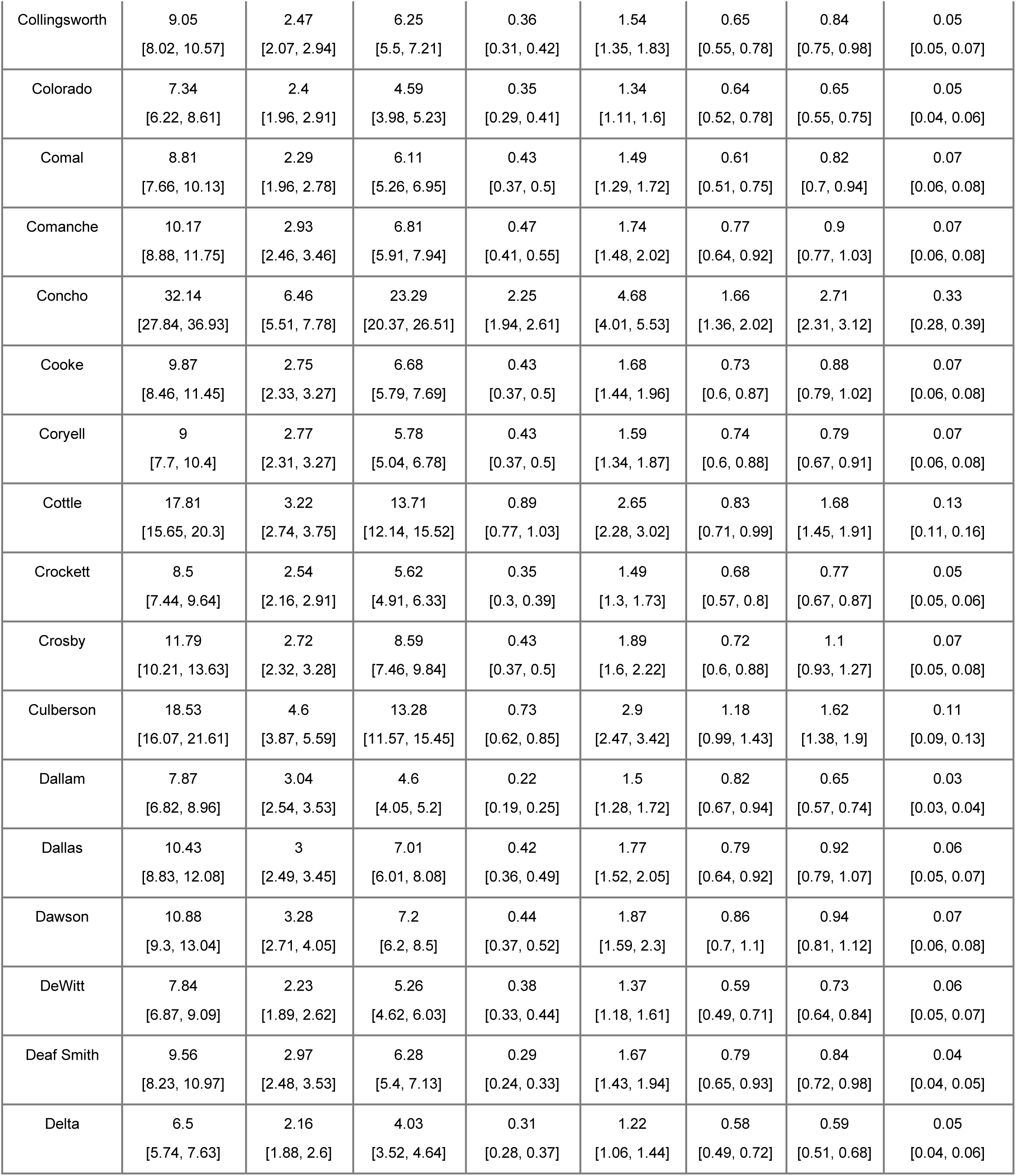
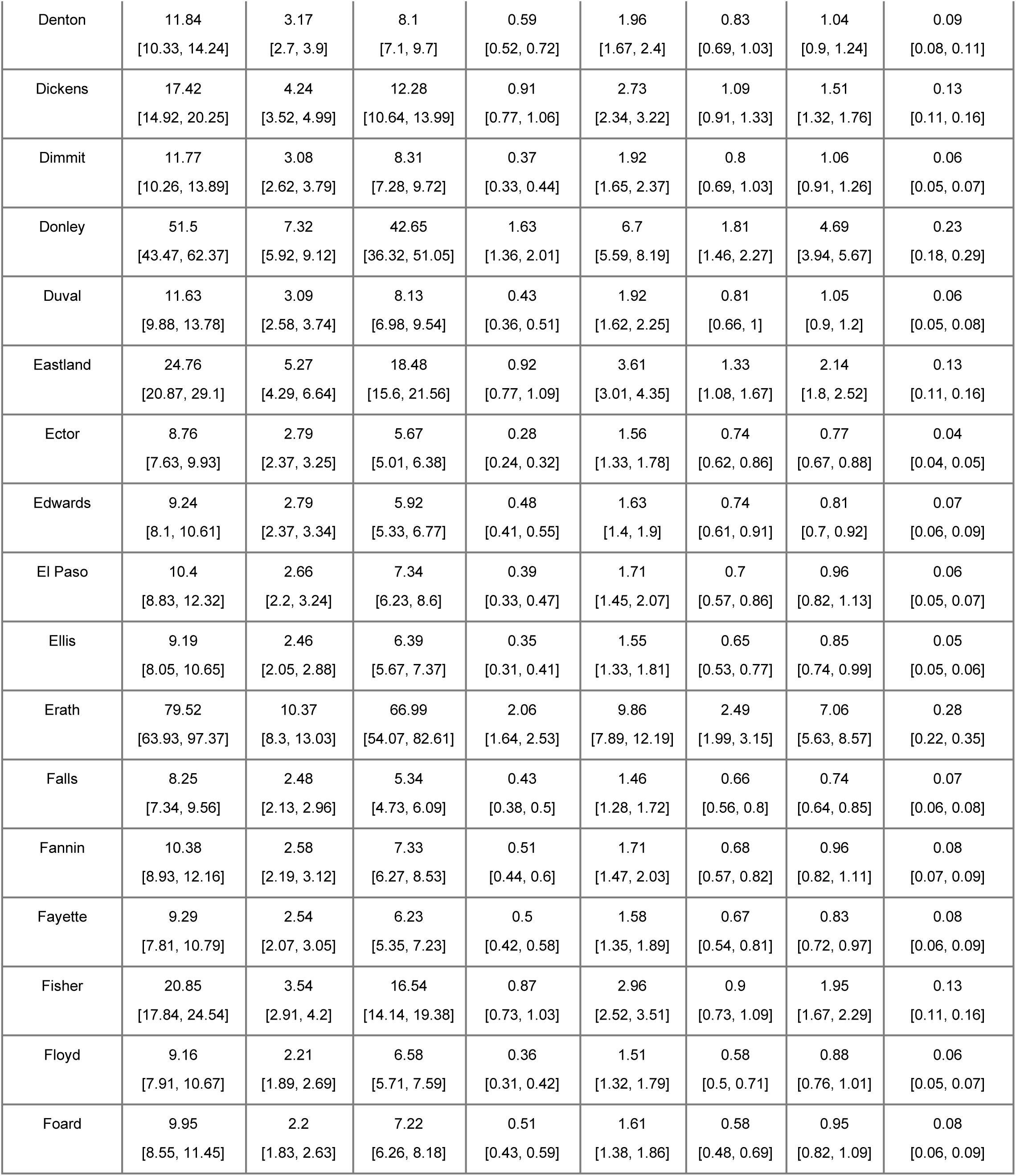
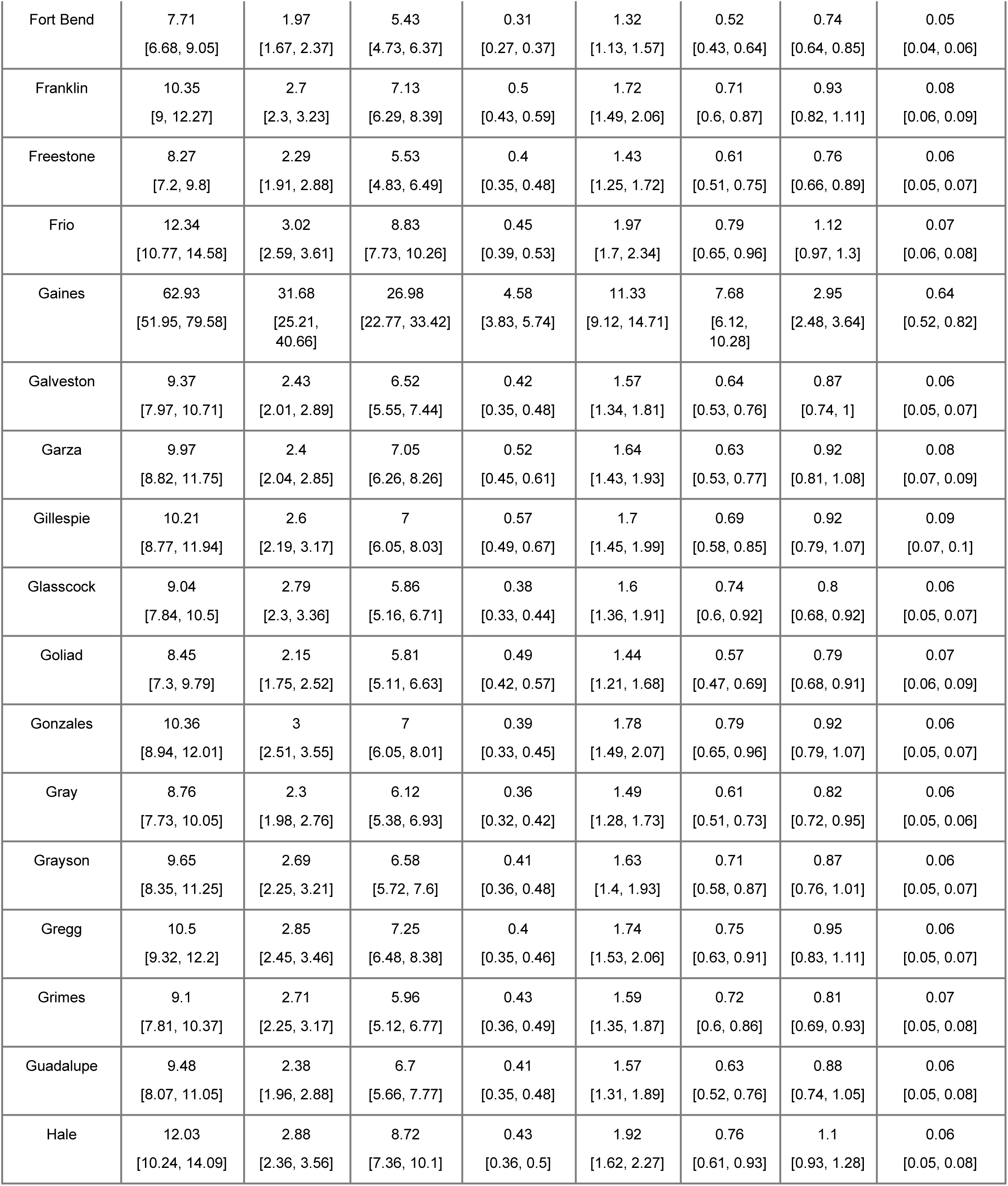
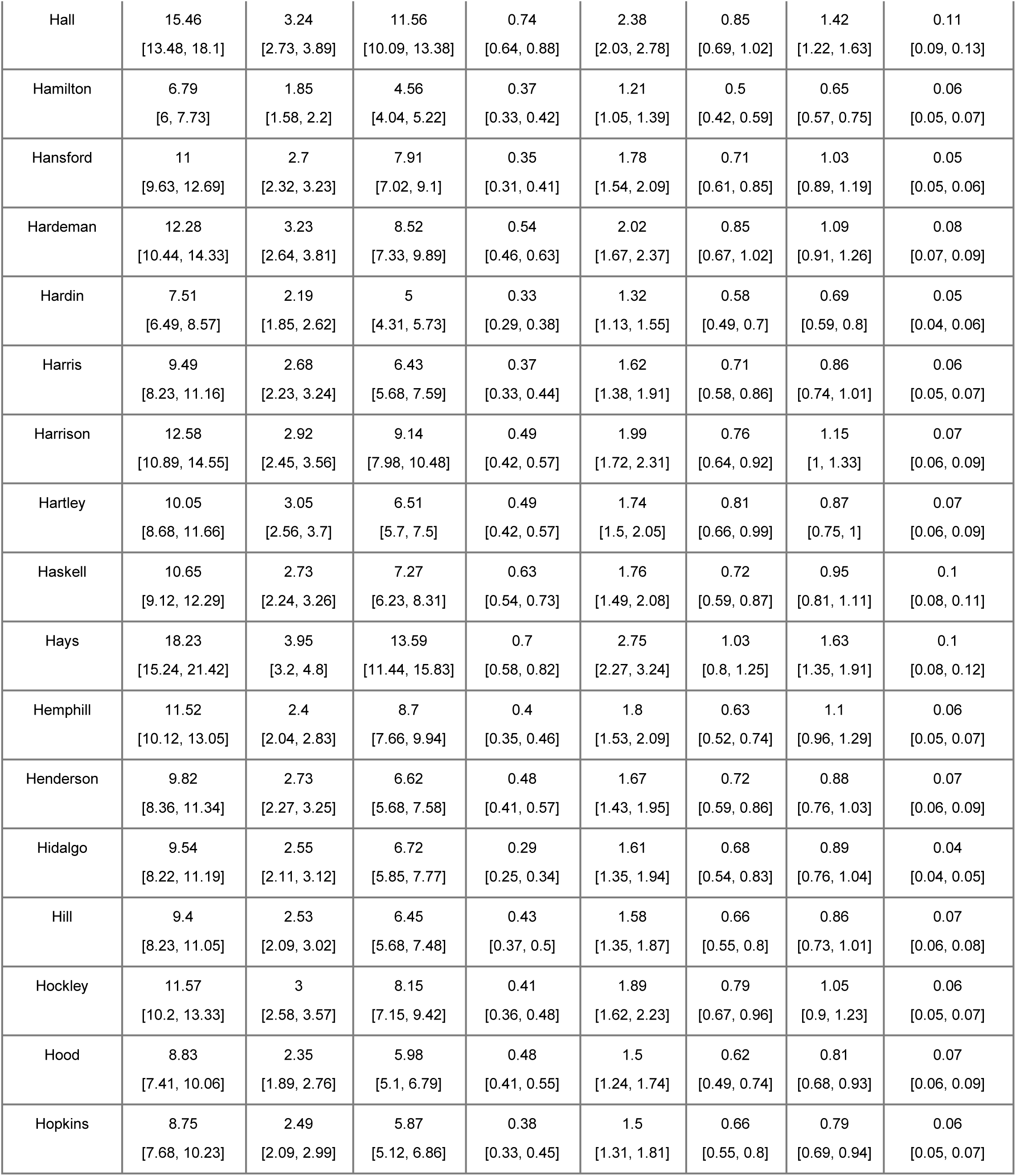
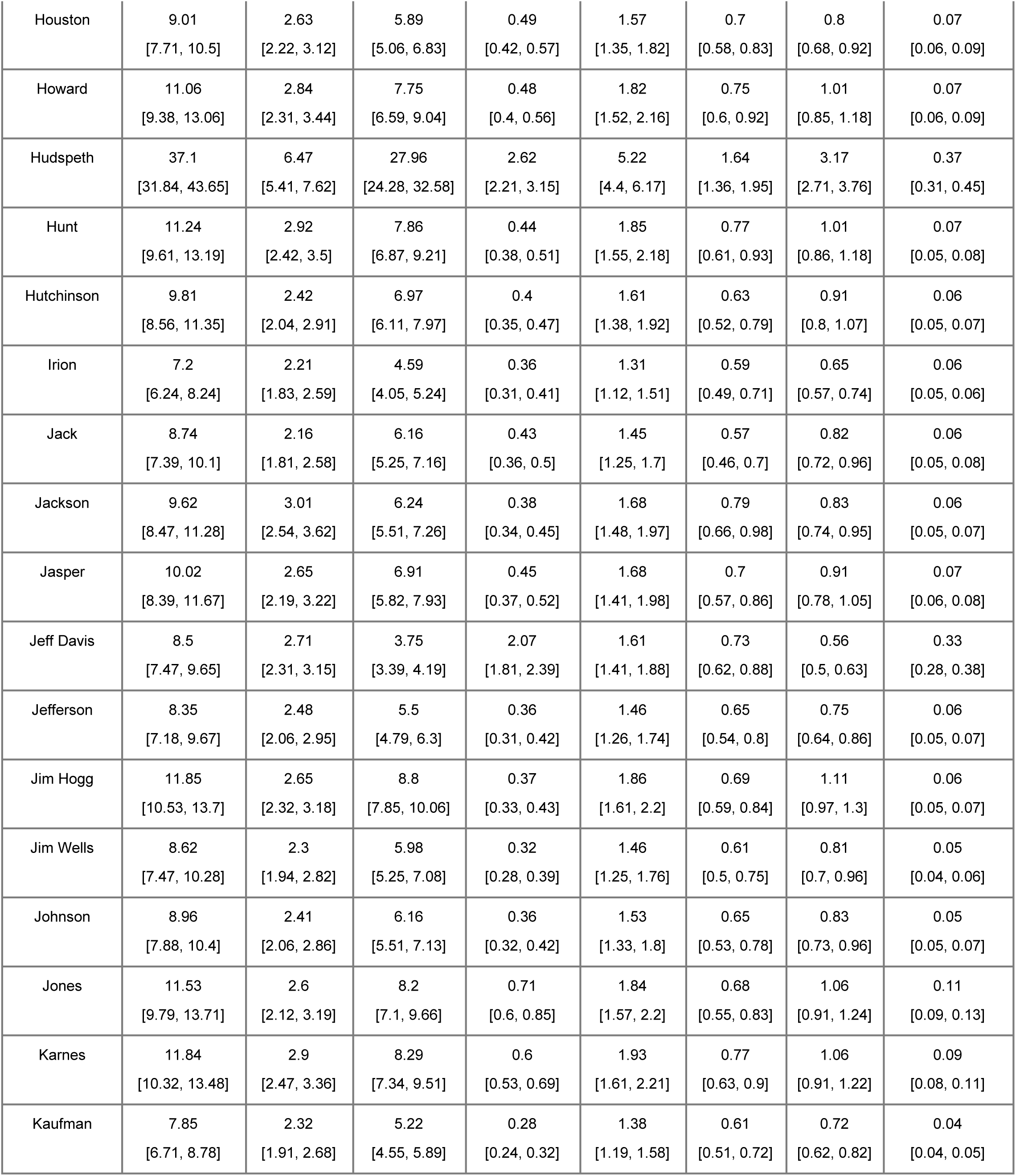
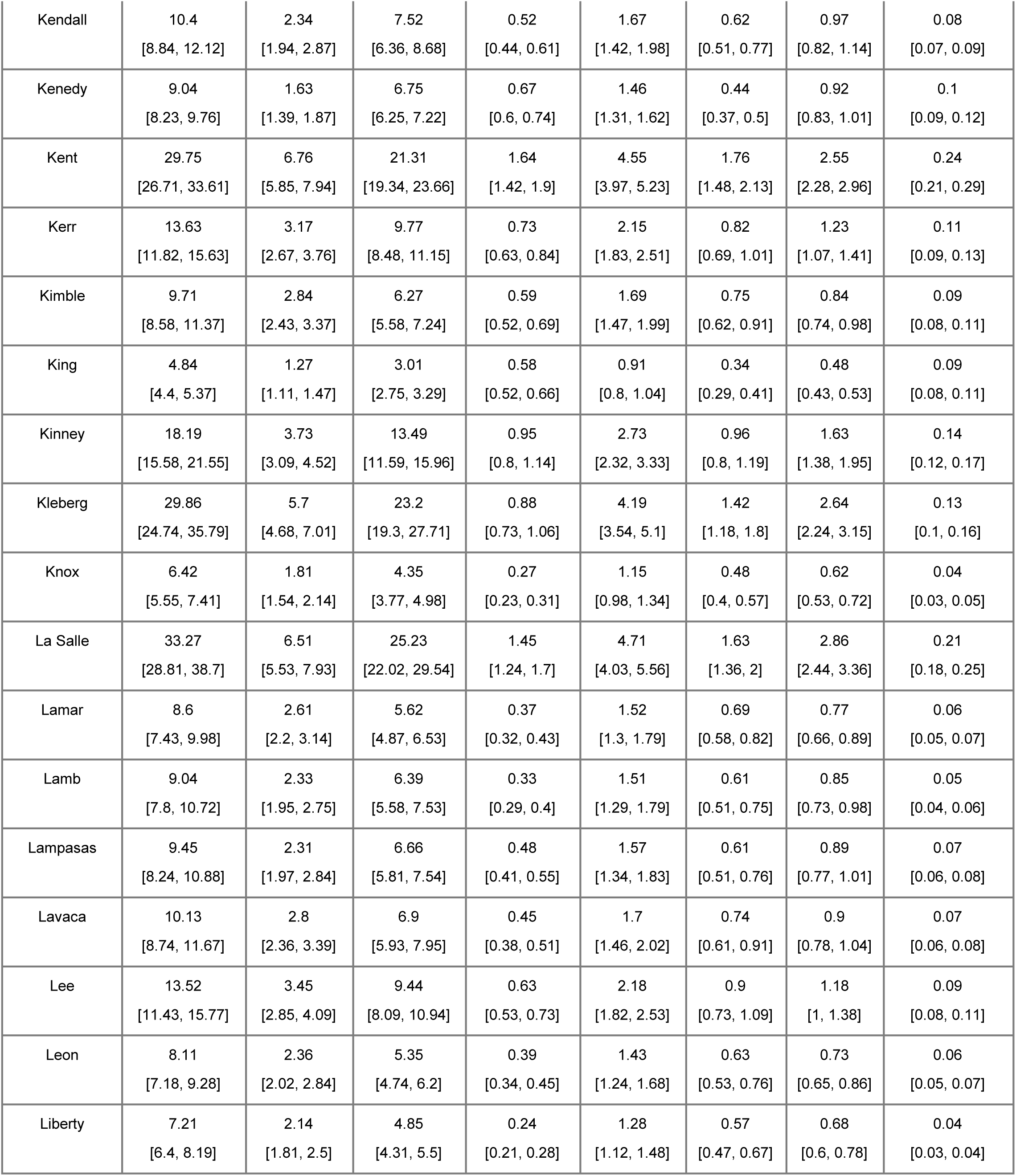
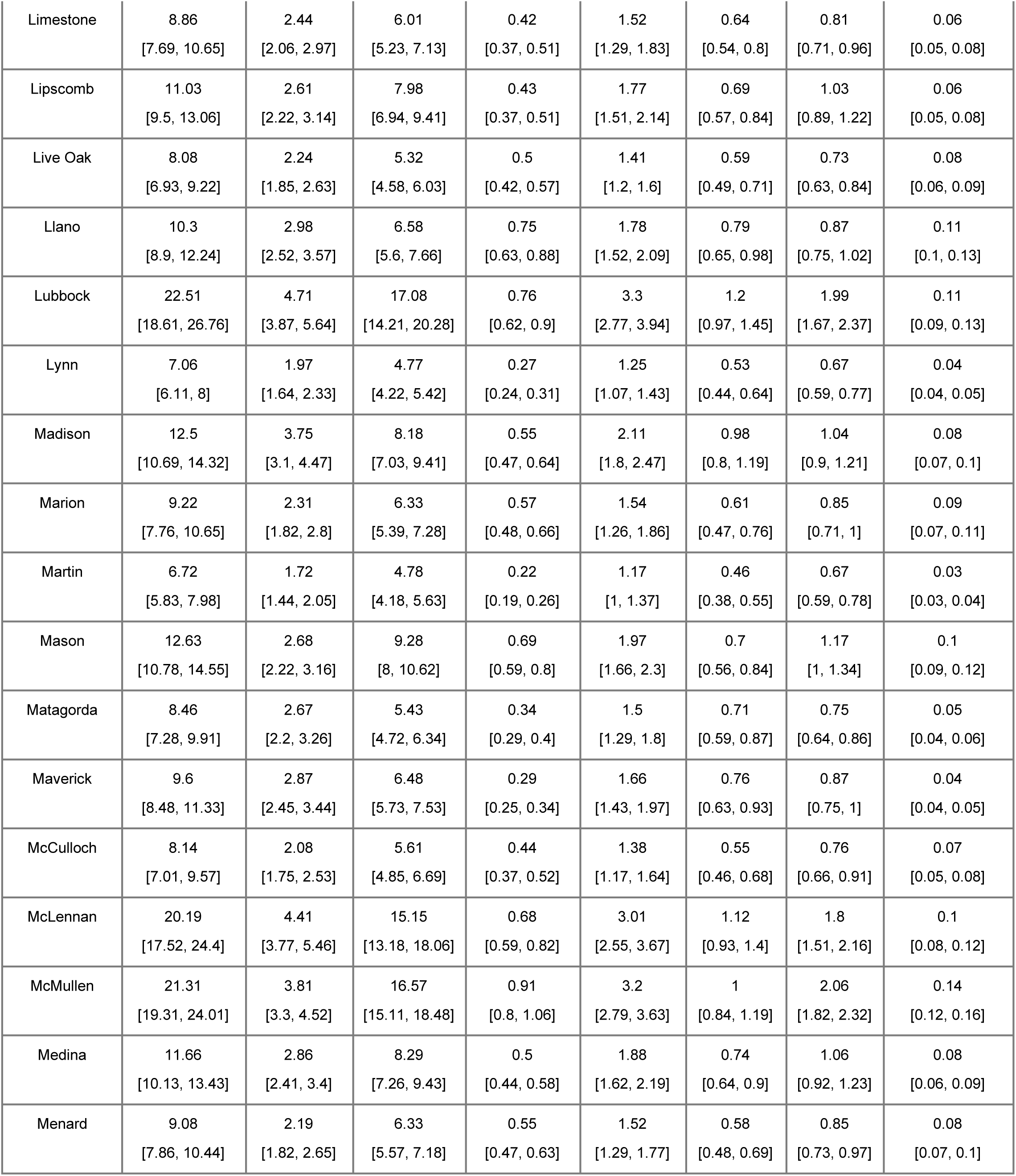
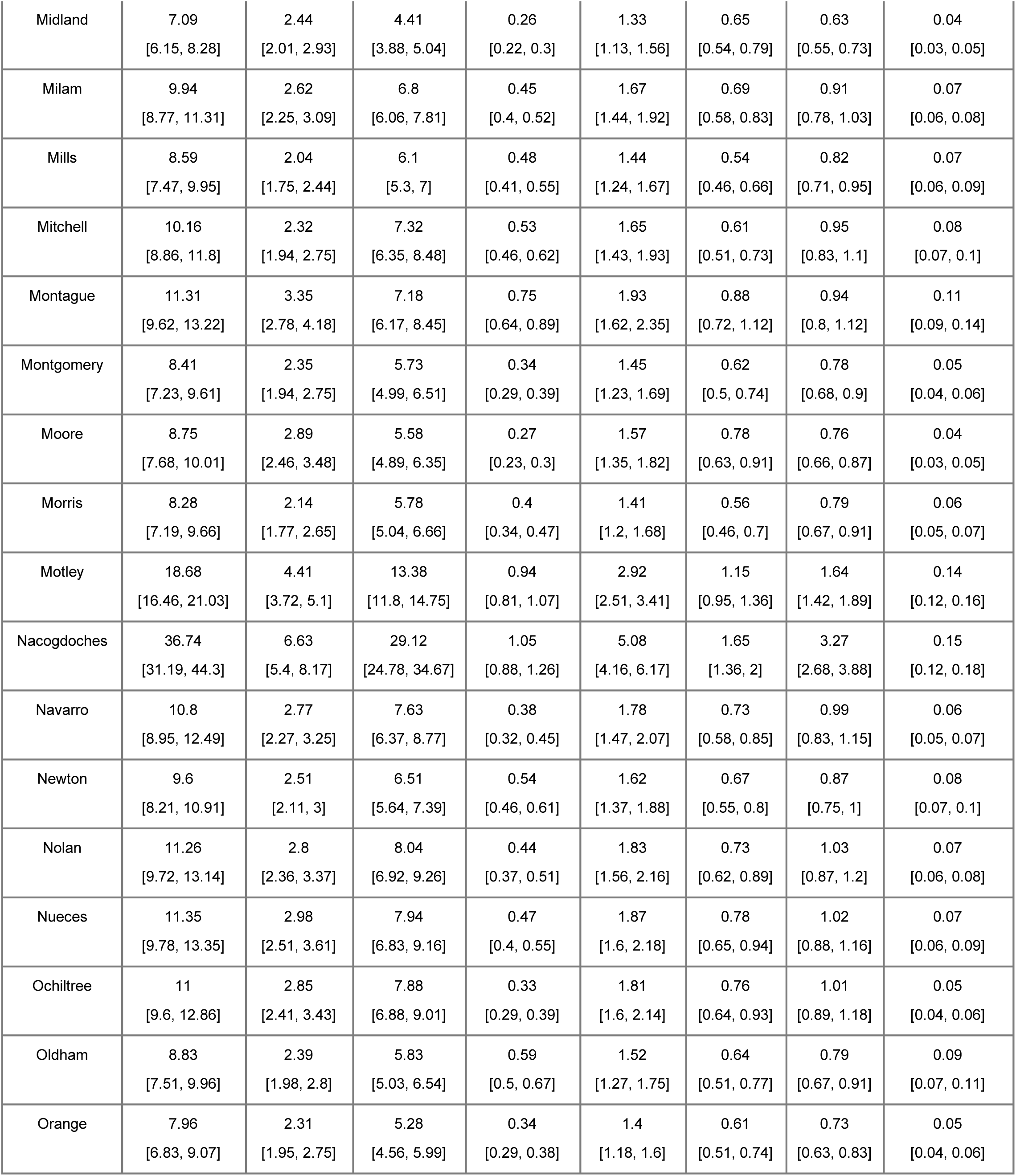
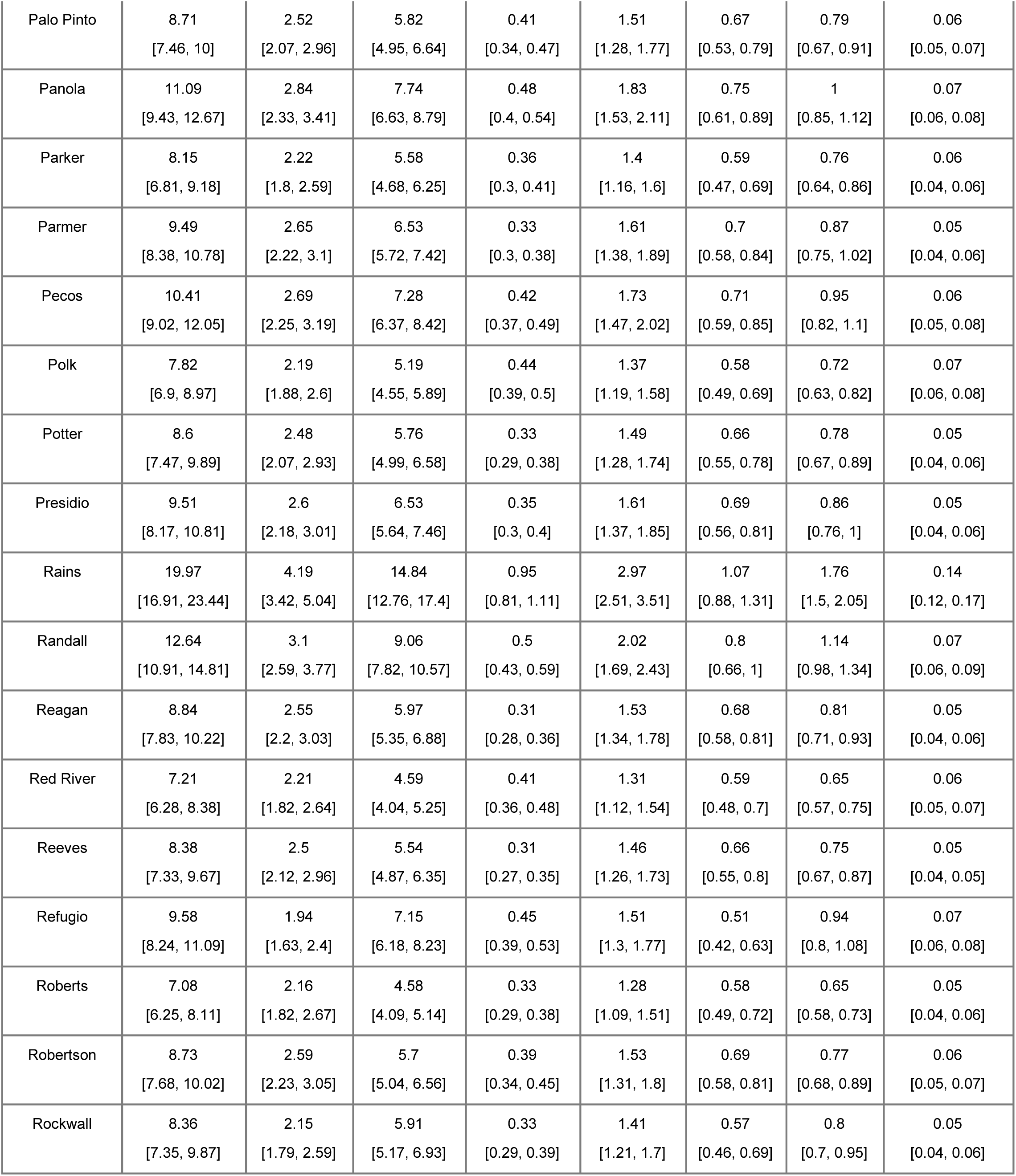
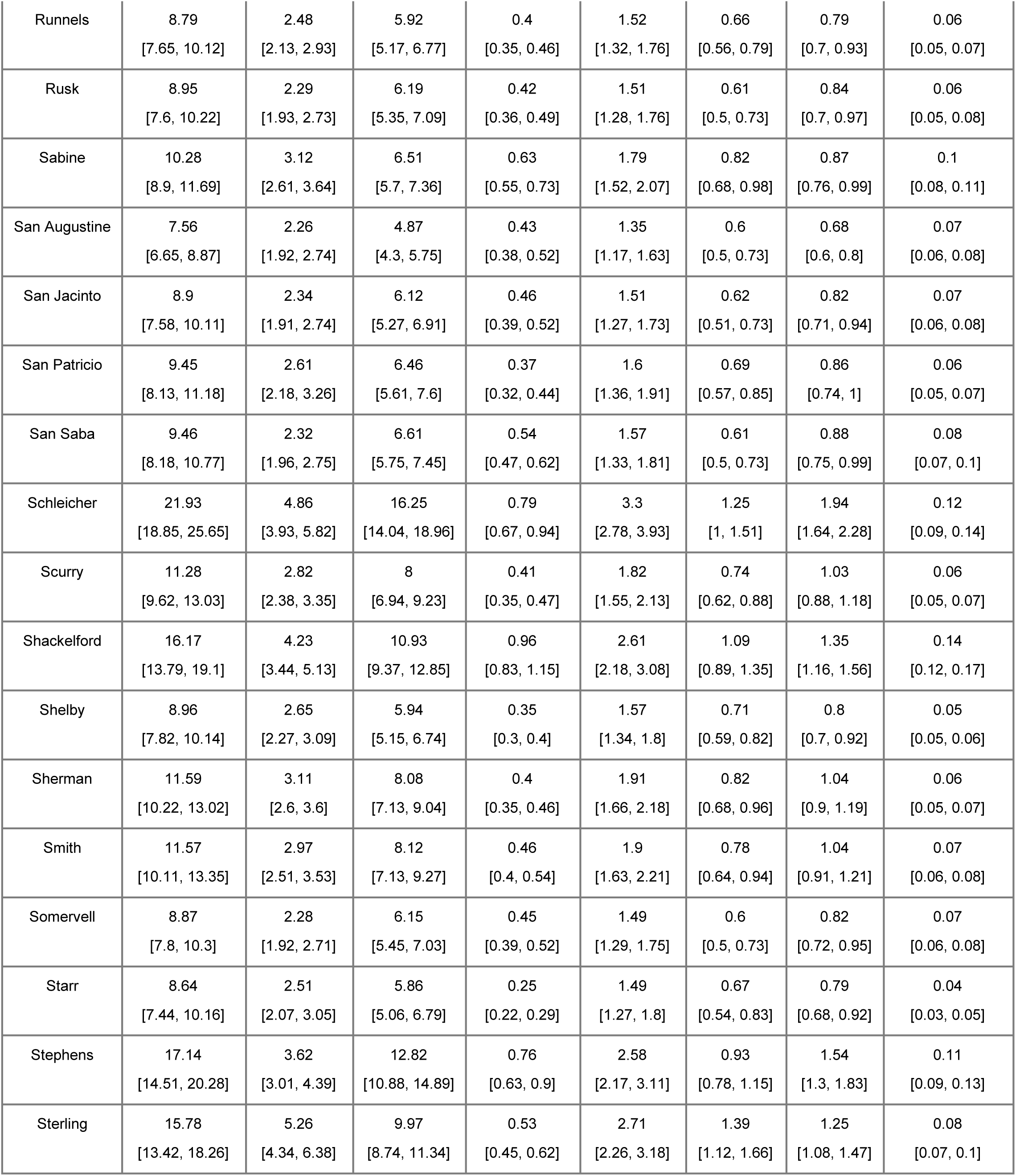
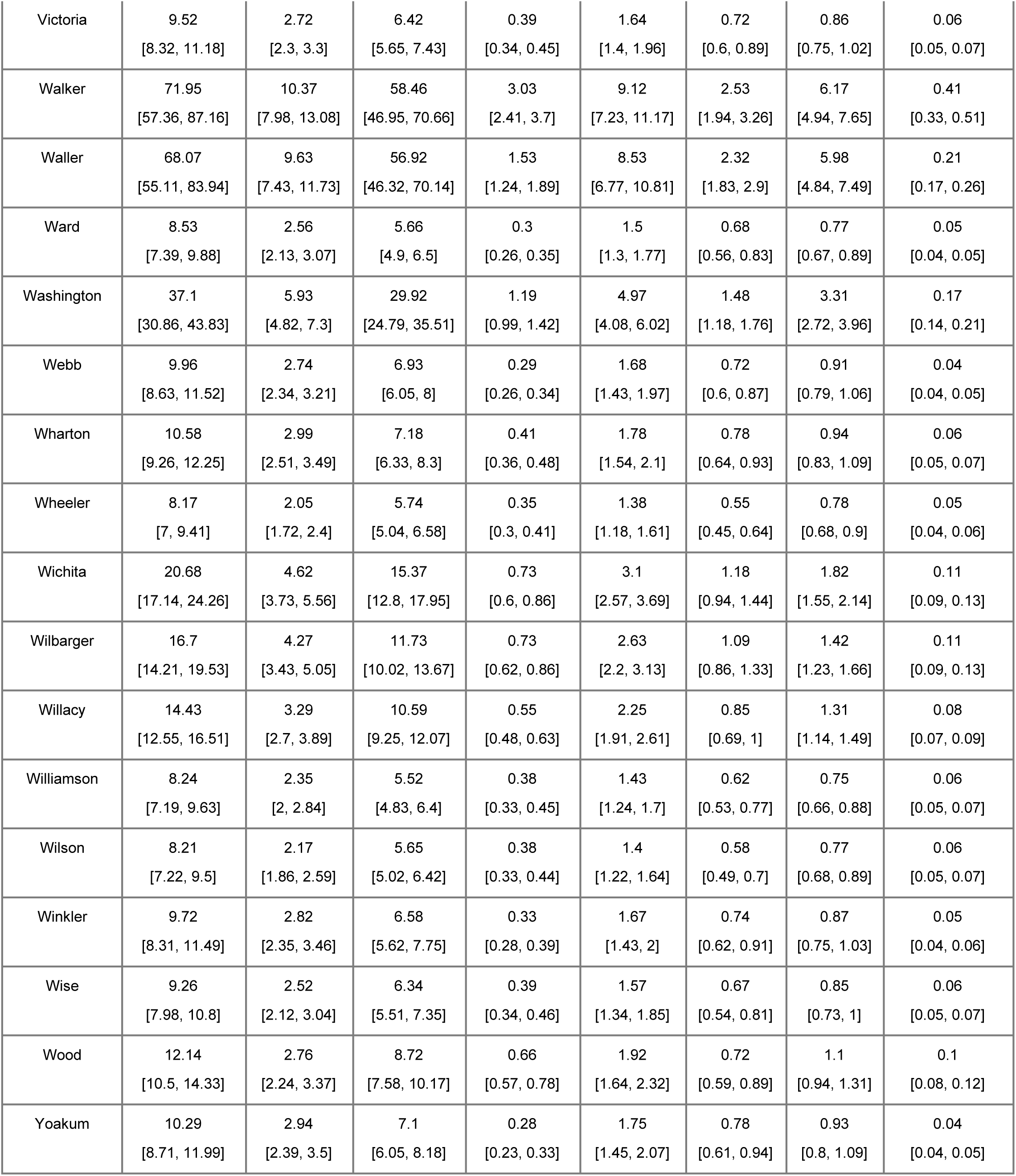
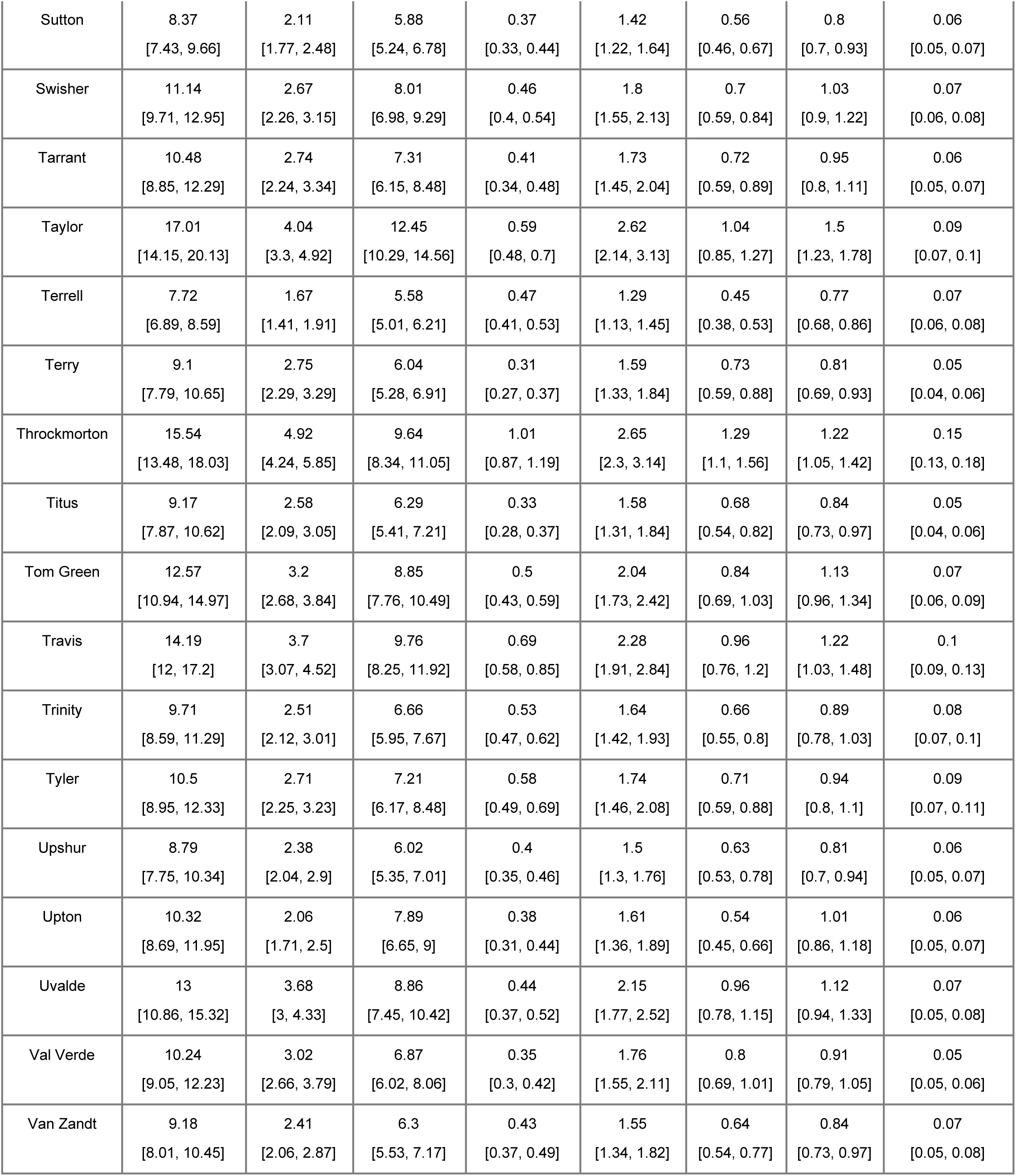
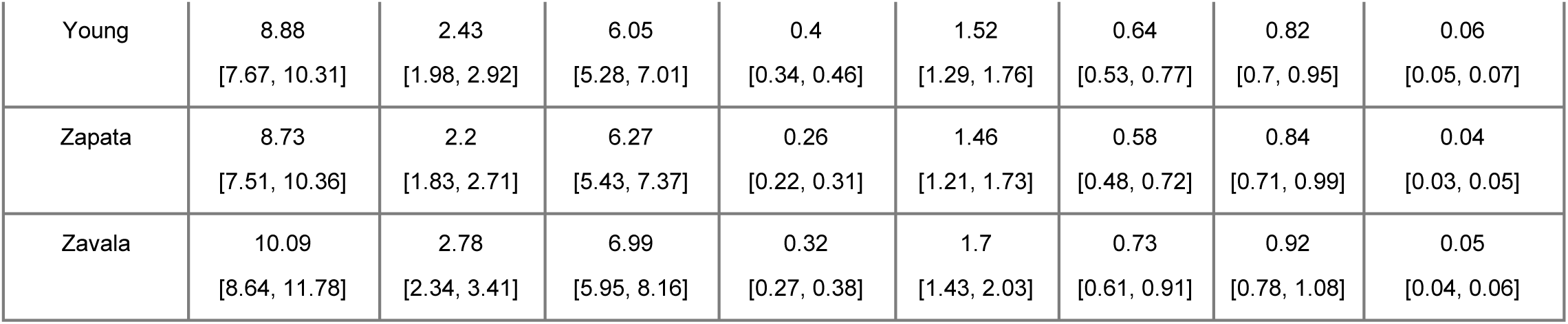
Age-specific cumulative case counts and hospitalizations derived from 5% increased vaccination uptake scenario projections.

| County Name | Total | 0-4 | 5-19 | 20+ | Total Hosp | 0-4 Hosp | 5-19 Hosp | 20+ Hosp |
| --- | --- | --- | --- | --- | --- | --- | --- | --- |
| Anderson | 9.57<br>[8.31, 10.73] | 2.73<br>[2.27, 3.19] | 6.29<br>[5.52, 7.09] | 0.53<br>[0.46, 0.6] | 1.65<br>[1.42, 1.89] | 0.72<br>[0.59, 0.85] | 0.84<br>[0.74, 0.96] | 0.08<br>[0.07, 0.09] |
| Andrews | 7.44<br>[6.37, 8.74] | 2.26<br>[1.85, 2.73] | 4.96<br>[4.31, 5.75] | 0.24<br>[0.21, 0.28] | 1.33<br>[1.12, 1.56] | 0.6<br>[0.48, 0.74] | 0.69<br>[0.6, 0.81] | 0.04<br>[0.03, 0.04] |
| Angelina | 9.87<br>[8.56, 11.38] | 2.74<br>[2.31, 3.26] | 6.76<br>[5.9, 7.74] | 0.39<br>[0.34, 0.46] | 1.68<br>[1.43, 1.96] | 0.73<br>[0.6, 0.87] | 0.9<br>[0.77, 1.03] | 0.06<br>[0.05, 0.07] |
| Aransas | 8.86<br>[7.51, 10.36] | 2.41<br>[2, 2.9] | 5.89<br>[4.98, 6.78] | 0.56<br>[0.47, 0.66] | 1.52<br>[1.27, 1.81] | 0.64<br>[0.52, 0.78] | 0.8<br>[0.67, 0.93] | 0.09<br>[0.07, 0.1] |
| Archer | 7.22<br>[6.31, 8.32] | 1.93<br>[1.64, 2.28] | 4.96<br>[4.32, 5.62] | 0.37<br>[0.32, 0.42] | 1.26<br>[1.09, 1.49] | 0.52<br>[0.43, 0.63] | 0.69<br>[0.6, 0.8] | 0.06<br>[0.05, 0.07] |
| Armstrong | 7.5<br>[6.58, 8.67] | 2.3<br>[1.95, 2.77] | 4.81<br>[4.24, 5.48] | 0.38<br>[0.34, 0.45] | 1.36<br>[1.15, 1.61] | 0.62<br>[0.51, 0.75] | 0.68<br>[0.59, 0.78] | 0.06<br>[0.05, 0.07] |
| Atascosa | 8.98<br>[7.7, 10.44] | 2.46<br>[2.05, 2.9] | 6.16<br>[5.41, 7.17] | 0.34<br>[0.29, 0.4] | 1.52<br>[1.31, 1.79] | 0.65<br>[0.54, 0.77] | 0.83<br>[0.72, 0.97] | 0.05<br>[0.04, 0.06] |
| Austin | 8.27<br>[7.09, 9.58] | 2.29<br>[1.87, 2.82] | 5.63<br>[4.82, 6.52] | 0.38<br>[0.33, 0.45] | 1.44<br>[1.2, 1.7] | 0.61<br>[0.49, 0.75] | 0.77<br>[0.66, 0.9] | 0.06<br>[0.05, 0.07] |
| Bailey | 11.38<br>[9.7, 13.22] | 3.24<br>[2.73, 3.92] | 7.88<br>[6.66, 8.94] | 0.35<br>[0.3, 0.4] | 1.92<br>[1.61, 2.24] | 0.85<br>[0.72, 1.04] | 1.02<br>[0.84, 1.16] | 0.05<br>[0.04, 0.06] |
| Bandera | 8.37<br>[7.27, 9.73] | 2.19<br>[1.9, 2.67] | 5.59<br>[4.88, 6.45] | 0.58<br>[0.5, 0.68] | 1.44<br>[1.23, 1.7] | 0.58<br>[0.48, 0.72] | 0.76<br>[0.65, 0.89] | 0.09<br>[0.07, 0.11] |
| Bastrop | 9.28<br>[8.06, 10.63] | 2.52<br>[2.13, 3.02] | 6.38<br>[5.61, 7.27] | 0.36<br>[0.31, 0.42] | 1.57<br>[1.33, 1.85] | 0.66<br>[0.55, 0.8] | 0.85<br>[0.72, 0.99] | 0.06<br>[0.05, 0.07] |
| Baylor | 8.24<br>[7.25, 9.34] | 2.35<br>[1.97, 2.77] | 5.46<br>[4.79, 6.18] | 0.41<br>[0.36, 0.47] | 1.44<br>[1.24, 1.7] | 0.63<br>[0.51, 0.76] | 0.74<br>[0.65, 0.86] | 0.06<br>[0.05, 0.07] |
| Bee | 14.97<br>[12.65, 17.07] | 3.41<br>[2.81, 4.13] | 10.88<br>[9.34, 12.38] | 0.69<br>[0.58, 0.8] | 2.33<br>[1.96, 2.76] | 0.89<br>[0.73, 1.07] | 1.33<br>[1.14, 1.57] | 0.1<br>[0.09, 0.12] |
| Bell | 8.99<br>[7.9, 10.56] | 2.8<br>[2.35, 3.36] | 5.86<br>[5.21, 6.81] | 0.33<br>[0.29, 0.38] | 1.6<br>[1.36, 1.89] | 0.75<br>[0.61, 0.9] | 0.8<br>[0.7, 0.92] | 0.05<br>[0.04, 0.06] |
| Bexar | 11.32<br>[9.59, 13.07] | 2.97<br>[2.51, 3.62] | 7.86<br>[6.69, 9.07] | 0.45<br>[0.38, 0.52] | 1.87<br>[1.57, 2.19] | 0.78<br>[0.65, 0.95] | 1.01<br>[0.85, 1.19] | 0.07<br>[0.06, 0.08] |
| Blanco | 18.18<br>[16.01, 22.52] | 4.9<br>[4.15, 6.22] | 11.3<br>[10.06, 13.84] | 1.97<br>[1.74, 2.46] | 2.95<br>[2.56, 3.66] | 1.27<br>[1.05, 1.64] | 1.39<br>[1.21, 1.67] | 0.29<br>[0.25, 0.36] |
| Borden | 10.25<br>[8.89, 11.51] | 2.07<br>[1.7, 2.44] | 7.64<br>[6.66, 8.48] | 0.53<br>[0.45, 0.61] | 1.63<br>[1.4, 1.87] | 0.55<br>[0.44, 0.66] | 1<br>[0.88, 1.12] | 0.08<br>[0.07, 0.09] |
| Bosque | 8.29<br>[7.13, 9.45] | 2.21<br>[1.83, 2.6] | 5.64<br>[4.89, 6.39] | 0.45<br>[0.39, 0.51] | 1.42<br>[1.22, 1.62] | 0.59<br>[0.49, 0.69] | 0.77<br>[0.67, 0.87] | 0.07<br>[0.06, 0.08] |
| Bowie | 8.74<br>[7.32, 10.26] | 2.4<br>[1.97, 2.84] | 5.97<br>[5.05, 6.98] | 0.39<br>[0.33, 0.46] | 1.5<br>[1.26, 1.78] | 0.64<br>[0.52, 0.77] | 0.8<br>[0.69, 0.93] | 0.06<br>[0.05, 0.07] |
| Brazoria | 8.3<br>[7.14, 9.49] | 2.3<br>[1.95, 2.72] | 5.66<br>[4.87, 6.4] | 0.35<br>[0.29, 0.4] | 1.44<br>[1.22, 1.66] | 0.62<br>[0.51, 0.71] | 0.77<br>[0.66, 0.88] | 0.05<br>[0.04, 0.06] |
| Brazos | 100.5<br>[81.53, 124.62] | 13.06<br>[10.56, 16.4] | 85.1<br>[68.75, 105.13] | 2.49<br>[2.02, 3.11] | 12.19<br>[9.87, 15.07] | 3.11<br>[2.48, 3.81] | 8.75<br>[7.07, 11] | 0.33<br>[0.27, 0.43] |
| Brewster | 56.8<br>[46.88, 68.69] | 8.83<br>[7.11, 11.06] | 45.63<br>[37.76, 55.16] | 2.23<br>[1.83, 2.71] | 7.45<br>[6.1, 9] | 2.17<br>[1.74, 2.68] | 4.97<br>[4.06, 6.07] | 0.31<br>[0.25, 0.38] |
| Briscoe | 8.58<br>[7.47, 9.82] | 2.32<br>[1.96, 2.75] | 5.78<br>[5.13, 6.54] | 0.45<br>[0.39, 0.52] | 1.48<br>[1.28, 1.72] | 0.62<br>[0.52, 0.74] | 0.79<br>[0.68, 0.9] | 0.07<br>[0.06, 0.08] |
| Brooks | 9.47<br>[8.14, 10.84] | 2.61<br>[2.22, 3.11] | 6.48<br>[5.54, 7.42] | 0.38<br>[0.32, 0.44] | 1.61<br>[1.37, 1.88] | 0.69<br>[0.58, 0.83] | 0.86<br>[0.73, 0.99] | 0.06<br>[0.05, 0.07] |
| Brown | 14.49<br>[12.34, 16.75] | 3.58<br>[3, 4.32] | 10.13<br>[8.75, 11.81] | 0.67<br>[0.57, 0.78] | 2.3<br>[1.95, 2.73] | 0.93<br>[0.76, 1.12] | 1.26<br>[1.07, 1.49] | 0.1<br>[0.08, 0.12] |
| Burleson | 7.37<br>[6.39, 8.51] | 2.26<br>[1.85, 2.67] | 4.73<br>[4.18, 5.47] | 0.38<br>[0.33, 0.44] | 1.33<br>[1.13, 1.55] | 0.6<br>[0.48, 0.72] | 0.67<br>[0.59, 0.77] | 0.06<br>[0.05, 0.07] |
| Burnet | 10.09<br>[8.49, 11.68] | 2.65<br>[2.19, 3.11] | 6.92<br>[5.91, 7.95] | 0.54<br>[0.45, 0.62] | 1.69<br>[1.4, 1.98] | 0.7<br>[0.57, 0.84] | 0.91<br>[0.76, 1.04] | 0.08<br>[0.07, 0.1] |
| Caldwell | 13.62<br>[11.57, 15.96] | 3.52<br>[2.84, 4.2] | 9.6<br>[8.18, 11.07] | 0.56<br>[0.47, 0.64] | 2.21<br>[1.82, 2.59] | 0.92<br>[0.73, 1.1] | 1.21<br>[1, 1.38] | 0.08<br>[0.07, 0.1] |
| Calhoun | 10.54<br>[9.08, 12.16] | 2.9<br>[2.4, 3.51] | 7.15<br>[6.16, 8.21] | 0.47<br>[0.4, 0.54] | 1.77<br>[1.49, 2.07] | 0.76<br>[0.63, 0.94] | 0.94<br>[0.79, 1.08] | 0.07<br>[0.06, 0.08] |
| Callahan | 7.7<br>[6.69, 8.96] | 2.05<br>[1.74, 2.43] | 5.26<br>[4.59, 6.1] | 0.4<br>[0.35, 0.47] | 1.33<br>[1.14, 1.56] | 0.55<br>[0.46, 0.66] | 0.72<br>[0.63, 0.84] | 0.06<br>[0.05, 0.07] |
| Cameron | 10.46<br>[9.07, 12.26] | 2.68<br>[2.26, 3.22] | 7.42<br>[6.49, 8.82] | 0.35<br>[0.3, 0.41] | 1.72<br>[1.48, 2.05] | 0.7<br>[0.59, 0.87] | 0.96<br>[0.82, 1.12] | 0.05<br>[0.04, 0.06] |
| Camp | 9.05<br>[7.74, 10.58] | 2.8<br>[2.32, 3.42] | 5.91<br>[5.11, 6.88] | 0.34<br>[0.29, 0.4] | 1.59<br>[1.33, 1.87] | 0.74<br>[0.6, 0.9] | 0.8<br>[0.69, 0.92] | 0.05<br>[0.04, 0.06] |
| Carson | 8.88<br>[7.78, 10.06] | 2.07<br>[1.73, 2.4] | 6.39<br>[5.59, 7.3] | 0.41<br>[0.35, 0.46] | 1.46<br>[1.24, 1.7] | 0.55<br>[0.45, 0.65] | 0.86<br>[0.74, 0.98] | 0.06<br>[0.05, 0.07] |
| Cass | 8.06<br>[6.98, 9.33] | 2.35<br>[1.9, 2.79] | 5.3<br>[4.66, 6.07] | 0.39<br>[0.34, 0.45] | 1.42<br>[1.18, 1.63] | 0.62<br>[0.5, 0.76] | 0.73<br>[0.62, 0.84] | 0.06<br>[0.05, 0.07] |
| Castro | 9.18<br>[7.97, 10.41] | 2.64<br>[2.21, 3.06] | 6.22<br>[5.46, 7] | 0.31<br>[0.26, 0.35] | 1.57<br>[1.35, 1.81] | 0.7<br>[0.57, 0.82] | 0.83<br>[0.73, 0.94] | 0.05<br>[0.04, 0.05] |
| Chambers | 7.8<br>[6.79, 9.01] | 2.18<br>[1.8, 2.58] | 5.34<br>[4.64, 6.14] | 0.29<br>[0.25, 0.34] | 1.35<br>[1.16, 1.58] | 0.58<br>[0.48, 0.69] | 0.73<br>[0.65, 0.85] | 0.04<br>[0.04, 0.05] |
| Cherokee | 8.38<br>[7.3, 9.87] | 2.51<br>[2.11, 2.99] | 5.53<br>[4.85, 6.46] | 0.35<br>[0.3, 0.42] | 1.48<br>[1.26, 1.76] | 0.67<br>[0.55, 0.81] | 0.76<br>[0.66, 0.88] | 0.05<br>[0.05, 0.06] |
| Childress | 14.36<br>[12.55, 16.83] | 3.18<br>[2.67, 3.78] | 10.46<br>[9.25, 12.26] | 0.73<br>[0.64, 0.86] | 2.24<br>[1.91, 2.62] | 0.83<br>[0.68, 0.99] | 1.29<br>[1.13, 1.55] | 0.11<br>[0.09, 0.13] |
| Clay | 9.12<br>[7.89, 10.58] | 2.41<br>[2.02, 2.9] | 6.22<br>[5.43, 7.19] | 0.49<br>[0.42, 0.57] | 1.55<br>[1.32, 1.79] | 0.64<br>[0.52, 0.78] | 0.84<br>[0.72, 0.96] | 0.07<br>[0.06, 0.09] |
| Cochran | 10.55<br>[8.98, 12.37] | 3.47<br>[2.89, 4.19] | 6.7<br>[5.78, 7.78] | 0.38<br>[0.33, 0.45] | 1.86<br>[1.56, 2.27] | 0.92<br>[0.74, 1.16] | 0.89<br>[0.76, 1.05] | 0.06<br>[0.05, 0.07] |
| Coke | 6.21<br>[5.47, 7.04] | 2.03<br>[1.72, 2.37] | 3.77<br>[3.31, 4.2] | 0.41<br>[0.36, 0.47] | 1.17<br>[1.01, 1.35] | 0.55<br>[0.45, 0.66] | 0.56<br>[0.49, 0.63] | 0.06<br>[0.05, 0.07] |
| Coleman | 9.34<br>[8.23, 10.83] | 2.59<br>[2.2, 3.04] | 6.28<br>[5.63, 7.26] | 0.49<br>[0.43, 0.56] | 1.6<br>[1.37, 1.9] | 0.69<br>[0.57, 0.82] | 0.85<br>[0.74, 0.98] | 0.07<br>[0.06, 0.09] |
| Collin | 10.34<br>[8.97, 11.66] | 2.5<br>[2.12, 2.93] | 7.38<br>[6.42, 8.42] | 0.42<br>[0.36, 0.48] | 1.69<br>[1.46, 1.93] | 0.66<br>[0.55, 0.79] | 0.96<br>[0.84, 1.1] | 0.06<br>[0.05, 0.07] |
| Collingsworth | 9.05<br>[8.02, 10.57] | 2.47<br>[2.07, 2.94] | 6.25<br>[5.5, 7.21] | 0.36<br>[0.31, 0.42] | 1.54<br>[1.35, 1.83] | 0.65<br>[0.55, 0.78] | 0.84<br>[0.75, 0.98] | 0.05<br>[0.05, 0.07] |
| Colorado | 7.34<br>[6.22, 8.61] | 2.4<br>[1.96, 2.91] | 4.59<br>[3.98, 5.23] | 0.35<br>[0.29, 0.41] | 1.34<br>[1.11, 1.6] | 0.64<br>[0.52, 0.78] | 0.65<br>[0.55, 0.75] | 0.05<br>[0.04, 0.06] |
| Comal | 8.81<br>[7.66, 10.13] | 2.29<br>[1.96, 2.78] | 6.11<br>[5.26, 6.95] | 0.43<br>[0.37, 0.5] | 1.49<br>[1.29, 1.72] | 0.61<br>[0.51, 0.75] | 0.82<br>[0.7, 0.94] | 0.07<br>[0.06, 0.08] |
| Comanche | 10.17<br>[8.88, 11.75] | 2.93<br>[2.46, 3.46] | 6.81<br>[5.91, 7.94] | 0.47<br>[0.41, 0.55] | 1.74<br>[1.48, 2.02] | 0.77<br>[0.64, 0.92] | 0.9<br>[0.77, 1.03] | 0.07<br>[0.06, 0.08] |
| Concho | 32.14<br>[27.84, 36.93] | 6.46<br>[5.51, 7.78] | 23.29<br>[20.37, 26.51] | 2.25<br>[1.94, 2.61] | 4.68<br>[4.01, 5.53] | 1.66<br>[1.36, 2.02] | 2.71<br>[2.31, 3.12] | 0.33<br>[0.28, 0.39] |
| Cooke | 9.87<br>[8.46, 11.45] | 2.75<br>[2.33, 3.27] | 6.68<br>[5.79, 7.69] | 0.43<br>[0.37, 0.5] | 1.68<br>[1.44, 1.96] | 0.73<br>[0.6, 0.87] | 0.88<br>[0.79, 1.02] | 0.07<br>[0.06, 0.08] |
| Coryell | 9<br>[7.7, 10.4] | 2.77<br>[2.31, 3.27] | 5.78<br>[5.04, 6.78] | 0.43<br>[0.37, 0.5] | 1.59<br>[1.34, 1.87] | 0.74<br>[0.6, 0.88] | 0.79<br>[0.67, 0.91] | 0.07<br>[0.06, 0.08] |
| Cottle | 17.81<br>[15.65, 20.3] | 3.22<br>[2.74, 3.75] | 13.71<br>[12.14, 15.52] | 0.89<br>[0.77, 1.03] | 2.65<br>[2.28, 3.02] | 0.83<br>[0.71, 0.99] | 1.68<br>[1.45, 1.91] | 0.13<br>[0.11, 0.16] |
| Crockett | 8.5<br>[7.44, 9.64] | 2.54<br>[2.16, 2.91] | 5.62<br>[4.91, 6.33] | 0.35<br>[0.3, 0.39] | 1.49<br>[1.3, 1.73] | 0.68<br>[0.57, 0.8] | 0.77<br>[0.67, 0.87] | 0.05<br>[0.05, 0.06] |
| Crosby | 11.79<br>[10.21, 13.63] | 2.72<br>[2.32, 3.28] | 8.59<br>[7.46, 9.84] | 0.43<br>[0.37, 0.5] | 1.89<br>[1.6, 2.22] | 0.72<br>[0.6, 0.88] | 1.1<br>[0.93, 1.27] | 0.07<br>[0.05, 0.08] |
| Culberson | 18.53<br>[16.07, 21.61] | 4.6<br>[3.87, 5.59] | 13.28<br>[11.57, 15.45] | 0.73<br>[0.62, 0.85] | 2.9<br>[2.47, 3.42] | 1.18<br>[0.99, 1.43] | 1.62<br>[1.38, 1.9] | 0.11<br>[0.09, 0.13] |
| Dallam | 7.87<br>[6.82, 8.96] | 3.04<br>[2.54, 3.53] | 4.6<br>[4.05, 5.2] | 0.22<br>[0.19, 0.25] | 1.5<br>[1.28, 1.72] | 0.82<br>[0.67, 0.94] | 0.65<br>[0.57, 0.74] | 0.03<br>[0.03, 0.04] |
| Dallas | 10.43<br>[8.83, 12.08] | 3<br>[2.49, 3.45] | 7.01<br>[6.01, 8.08] | 0.42<br>[0.36, 0.49] | 1.77<br>[1.52, 2.05] | 0.79<br>[0.64, 0.92] | 0.92<br>[0.79, 1.07] | 0.06<br>[0.05, 0.07] |
| Dawson | 10.88<br>[9.3, 13.04] | 3.28<br>[2.71, 4.05] | 7.2<br>[6.2, 8.5] | 0.44<br>[0.37, 0.52] | 1.87<br>[1.59, 2.3] | 0.86<br>[0.7, 1.1] | 0.94<br>[0.81, 1.12] | 0.07<br>[0.06, 0.08] |
| DeWitt | 7.84<br>[6.87, 9.09] | 2.23<br>[1.89, 2.62] | 5.26<br>[4.62, 6.03] | 0.38<br>[0.33, 0.44] | 1.37<br>[1.18, 1.61] | 0.59<br>[0.49, 0.71] | 0.73<br>[0.64, 0.84] | 0.06<br>[0.05, 0.07] |
| Deaf Smith | 9.56<br>[8.23, 10.97] | 2.97<br>[2.48, 3.53] | 6.28<br>[5.4, 7.13] | 0.29<br>[0.24, 0.33] | 1.67<br>[1.43, 1.94] | 0.79<br>[0.65, 0.93] | 0.84<br>[0.72, 0.98] | 0.04<br>[0.04, 0.05] |
| Delta | 6.5<br>[5.74, 7.63] | 2.16<br>[1.88, 2.6] | 4.03<br>[3.52, 4.64] | 0.31<br>[0.28, 0.37] | 1.22<br>[1.06, 1.44] | 0.58<br>[0.49, 0.72] | 0.59<br>[0.51, 0.68] | 0.05<br>[0.04, 0.06] |
| Denton | 11.84<br>[10.33, 14.24] | 3.17<br>[2.7, 3.9] | 8.1<br>[7.1, 9.7] | 0.59<br>[0.52, 0.72] | 1.96<br>[1.67, 2.4] | 0.83<br>[0.69, 1.03] | 1.04<br>[0.9, 1.24] | 0.09<br>[0.08, 0.11] |
| Dickens | 17.42<br>[14.92, 20.25] | 4.24<br>[3.52, 4.99] | 12.28<br>[10.64, 13.99] | 0.91<br>[0.77, 1.06] | 2.73<br>[2.34, 3.22] | 1.09<br>[0.91, 1.33] | 1.51<br>[1.32, 1.76] | 0.13<br>[0.11, 0.16] |
| Dimmit | 11.77<br>[10.26, 13.89] | 3.08<br>[2.62, 3.79] | 8.31<br>[7.28, 9.72] | 0.37<br>[0.33, 0.44] | 1.92<br>[1.65, 2.37] | 0.8<br>[0.69, 1.03] | 1.06<br>[0.91, 1.26] | 0.06<br>[0.05, 0.07] |
| Donley | 51.5<br>[43.47, 62.37] | 7.32<br>[5.92, 9.12] | 42.65<br>[36.32, 51.05] | 1.63<br>[1.36, 2.01] | 6.7<br>[5.59, 8.19] | 1.81<br>[1.46, 2.27] | 4.69<br>[3.94, 5.67] | 0.23<br>[0.18, 0.29] |
| Duval | 11.63<br>[9.88, 13.78] | 3.09<br>[2.58, 3.74] | 8.13<br>[6.98, 9.54] | 0.43<br>[0.36, 0.51] | 1.92<br>[1.62, 2.25] | 0.81<br>[0.66, 1] | 1.05<br>[0.9, 1.2] | 0.06<br>[0.05, 0.08] |
| Eastland | 24.76<br>[20.87, 29.1] | 5.27<br>[4.29, 6.64] | 18.48<br>[15.6, 21.56] | 0.92<br>[0.77, 1.09] | 3.61<br>[3.01, 4.35] | 1.33<br>[1.08, 1.67] | 2.14<br>[1.8, 2.52] | 0.13<br>[0.11, 0.16] |
| Ector | 8.76<br>[7.63, 9.93] | 2.79<br>[2.37, 3.25] | 5.67<br>[5.01, 6.38] | 0.28<br>[0.24, 0.32] | 1.56<br>[1.33, 1.78] | 0.74<br>[0.62, 0.86] | 0.77<br>[0.67, 0.88] | 0.04<br>[0.04, 0.05] |
| Edwards | 9.24<br>[8.1, 10.61] | 2.79<br>[2.37, 3.34] | 5.92<br>[5.33, 6.77] | 0.48<br>[0.41, 0.55] | 1.63<br>[1.4, 1.9] | 0.74<br>[0.61, 0.91] | 0.81<br>[0.7, 0.92] | 0.07<br>[0.06, 0.09] |
| El Paso | 10.4<br>[8.83, 12.32] | 2.66<br>[2.2, 3.24] | 7.34<br>[6.23, 8.6] | 0.39<br>[0.33, 0.47] | 1.71<br>[1.45, 2.07] | 0.7<br>[0.57, 0.86] | 0.96<br>[0.82, 1.13] | 0.06<br>[0.05, 0.07] |
| Ellis | 9.19<br>[8.05, 10.65] | 2.46<br>[2.05, 2.88] | 6.39<br>[5.67, 7.37] | 0.35<br>[0.31, 0.41] | 1.55<br>[1.33, 1.81] | 0.65<br>[0.53, 0.77] | 0.85<br>[0.74, 0.99] | 0.05<br>[0.05, 0.06] |
| Erath | 79.52<br>[63.93, 97.37] | 10.37<br>[8.3, 13.03] | 66.99<br>[54.07, 82.61] | 2.06<br>[1.64, 2.53] | 9.86<br>[7.89, 12.19] | 2.49<br>[1.99, 3.15] | 7.06<br>[5.63, 8.57] | 0.28<br>[0.22, 0.35] |
| Falls | 8.25<br>[7.34, 9.56] | 2.48<br>[2.13, 2.96] | 5.34<br>[4.73, 6.09] | 0.43<br>[0.38, 0.5] | 1.46<br>[1.28, 1.72] | 0.66<br>[0.56, 0.8] | 0.74<br>[0.64, 0.85] | 0.07<br>[0.06, 0.08] |
| Fannin | 10.38<br>[8.93, 12.16] | 2.58<br>[2.19, 3.12] | 7.33<br>[6.27, 8.53] | 0.51<br>[0.44, 0.6] | 1.71<br>[1.47, 2.03] | 0.68<br>[0.57, 0.82] | 0.96<br>[0.82, 1.11] | 0.08<br>[0.07, 0.09] |
| Fayette | 9.29<br>[7.81, 10.79] | 2.54<br>[2.07, 3.05] | 6.23<br>[5.35, 7.23] | 0.5<br>[0.42, 0.58] | 1.58<br>[1.35, 1.89] | 0.67<br>[0.54, 0.81] | 0.83<br>[0.72, 0.97] | 0.08<br>[0.06, 0.09] |
| Fisher | 20.85<br>[17.84, 24.54] | 3.54<br>[2.91, 4.2] | 16.54<br>[14.14, 19.38] | 0.87<br>[0.73, 1.03] | 2.96<br>[2.52, 3.51] | 0.9<br>[0.73, 1.09] | 1.95<br>[1.67, 2.29] | 0.13<br>[0.11, 0.16] |
| Floyd | 9.16<br>[7.91, 10.67] | 2.21<br>[1.89, 2.69] | 6.58<br>[5.71, 7.59] | 0.36<br>[0.31, 0.42] | 1.51<br>[1.32, 1.79] | 0.58<br>[0.5, 0.71] | 0.88<br>[0.76, 1.01] | 0.06<br>[0.05, 0.07] |
| Foard | 9.95<br>[8.55, 11.45] | 2.2<br>[1.83, 2.63] | 7.22<br>[6.26, 8.18] | 0.51<br>[0.43, 0.59] | 1.61<br>[1.38, 1.86] | 0.58<br>[0.48, 0.69] | 0.95<br>[0.82, 1.09] | 0.08<br>[0.06, 0.09] |
| Fort Bend | 7.71<br>[6.68, 9.05] | 1.97<br>[1.67, 2.37] | 5.43<br>[4.73, 6.37] | 0.31<br>[0.27, 0.37] | 1.32<br>[1.13, 1.57] | 0.52<br>[0.43, 0.64] | 0.74<br>[0.64, 0.85] | 0.05<br>[0.04, 0.06] |
| Franklin | 10.35<br>[9, 12.27] | 2.7<br>[2.3, 3.23] | 7.13<br>[6.29, 8.39] | 0.5<br>[0.43, 0.59] | 1.72<br>[1.49, 2.06] | 0.71<br>[0.6, 0.87] | 0.93<br>[0.82, 1.11] | 0.08<br>[0.06, 0.09] |
| Freestone | 8.27<br>[7.2, 9.8] | 2.29<br>[1.91, 2.88] | 5.53<br>[4.83, 6.49] | 0.4<br>[0.35, 0.48] | 1.43<br>[1.25, 1.72] | 0.61<br>[0.51, 0.75] | 0.76<br>[0.66, 0.89] | 0.06<br>[0.05, 0.07] |
| Frio | 12.34<br>[10.77, 14.58] | 3.02<br>[2.59, 3.61] | 8.83<br>[7.73, 10.26] | 0.45<br>[0.39, 0.53] | 1.97<br>[1.7, 2.34] | 0.79<br>[0.65, 0.96] | 1.12<br>[0.97, 1.3] | 0.07<br>[0.06, 0.08] |
| Gaines | 62.93<br>[51.95, 79.58] | 31.68<br>[25.21, 40.66] | 26.98<br>[22.77, 33.42] | 4.58<br>[3.83, 5.74] | 11.33<br>[9.12, 14.71] | 7.68<br>[6.12, 10.28] | 2.95<br>[2.48, 3.64] | 0.64<br>[0.52, 0.82] |
| Galveston | 9.37<br>[7.97, 10.71] | 2.43<br>[2.01, 2.89] | 6.52<br>[5.55, 7.44] | 0.42<br>[0.35, 0.48] | 1.57<br>[1.34, 1.81] | 0.64<br>[0.53, 0.76] | 0.87<br>[0.74, 1] | 0.06<br>[0.05, 0.07] |
| Garza | 9.97<br>[8.82, 11.75] | 2.4<br>[2.04, 2.85] | 7.05<br>[6.26, 8.26] | 0.52<br>[0.45, 0.61] | 1.64<br>[1.43, 1.93] | 0.63<br>[0.53, 0.77] | 0.92<br>[0.81, 1.08] | 0.08<br>[0.07, 0.09] |
| Gillespie | 10.21<br>[8.77, 11.94] | 2.6<br>[2.19, 3.17] | 7<br>[6.05, 8.03] | 0.57<br>[0.49, 0.67] | 1.7<br>[1.45, 1.99] | 0.69<br>[0.58, 0.85] | 0.92<br>[0.79, 1.07] | 0.09<br>[0.07, 0.1] |
| Glasscock | 9.04<br>[7.84, 10.5] | 2.79<br>[2.3, 3.36] | 5.86<br>[5.16, 6.71] | 0.38<br>[0.33, 0.44] | 1.6<br>[1.36, 1.91] | 0.74<br>[0.6, 0.92] | 0.8<br>[0.68, 0.92] | 0.06<br>[0.05, 0.07] |
| Goliad | 8.45<br>[7.3, 9.79] | 2.15<br>[1.75, 2.52] | 5.81<br>[5.11, 6.63] | 0.49<br>[0.42, 0.57] | 1.44<br>[1.21, 1.68] | 0.57<br>[0.47, 0.69] | 0.79<br>[0.68, 0.91] | 0.07<br>[0.06, 0.09] |
| Gonzales | 10.36<br>[8.94, 12.01] | 3<br>[2.51, 3.55] | 7<br>[6.05, 8.01] | 0.39<br>[0.33, 0.45] | 1.78<br>[1.49, 2.07] | 0.79<br>[0.65, 0.96] | 0.92<br>[0.79, 1.07] | 0.06<br>[0.05, 0.07] |
| Gray | 8.76<br>[7.73, 10.05] | 2.3<br>[1.98, 2.76] | 6.12<br>[5.38, 6.93] | 0.36<br>[0.32, 0.42] | 1.49<br>[1.28, 1.73] | 0.61<br>[0.51, 0.73] | 0.82<br>[0.72, 0.95] | 0.06<br>[0.05, 0.06] |
| Grayson | 9.65<br>[8.35, 11.25] | 2.69<br>[2.25, 3.21] | 6.58<br>[5.72, 7.6] | 0.41<br>[0.36, 0.48] | 1.63<br>[1.4, 1.93] | 0.71<br>[0.58, 0.87] | 0.87<br>[0.76, 1.01] | 0.06<br>[0.05, 0.07] |
| Gregg | 10.5<br>[9.32, 12.2] | 2.85<br>[2.45, 3.46] | 7.25<br>[6.48, 8.38] | 0.4<br>[0.35, 0.46] | 1.74<br>[1.53, 2.06] | 0.75<br>[0.63, 0.91] | 0.95<br>[0.83, 1.11] | 0.06<br>[0.05, 0.07] |
| Grimes | 9.1<br>[7.81, 10.37] | 2.71<br>[2.25, 3.17] | 5.96<br>[5.12, 6.77] | 0.43<br>[0.36, 0.49] | 1.59<br>[1.35, 1.87] | 0.72<br>[0.6, 0.86] | 0.81<br>[0.69, 0.93] | 0.07<br>[0.05, 0.08] |
| Guadalupe | 9.48<br>[8.07, 11.05] | 2.38<br>[1.96, 2.88] | 6.7<br>[5.66, 7.77] | 0.41<br>[0.35, 0.48] | 1.57<br>[1.31, 1.89] | 0.63<br>[0.52, 0.76] | 0.88<br>[0.74, 1.05] | 0.06<br>[0.05, 0.08] |
| Hale | 12.03<br>[10.24, 14.09] | 2.88<br>[2.36, 3.56] | 8.72<br>[7.36, 10.1] | 0.43<br>[0.36, 0.5] | 1.92<br>[1.62, 2.27] | 0.76<br>[0.61, 0.93] | 1.1<br>[0.93, 1.28] | 0.06<br>[0.05, 0.08] |
| Hall | 15.46<br>[13.48, 18.1] | 3.24<br>[2.73, 3.89] | 11.56<br>[10.09, 13.38] | 0.74<br>[0.64, 0.88] | 2.38<br>[2.03, 2.78] | 0.85<br>[0.69, 1.02] | 1.42<br>[1.22, 1.63] | 0.11<br>[0.09, 0.13] |
| Hamilton | 6.79<br>[6, 7.73] | 1.85<br>[1.58, 2.2] | 4.56<br>[4.04, 5.22] | 0.37<br>[0.33, 0.42] | 1.21<br>[1.05, 1.39] | 0.5<br>[0.42, 0.59] | 0.65<br>[0.57, 0.75] | 0.06<br>[0.05, 0.07] |
| Hansford | 11<br>[9.63, 12.69] | 2.7<br>[2.32, 3.23] | 7.91<br>[7.02, 9.1] | 0.35<br>[0.31, 0.41] | 1.78<br>[1.54, 2.09] | 0.71<br>[0.61, 0.85] | 1.03<br>[0.89, 1.19] | 0.05<br>[0.05, 0.06] |
| Hardeman | 12.28<br>[10.44, 14.33] | 3.23<br>[2.64, 3.81] | 8.52<br>[7.33, 9.89] | 0.54<br>[0.46, 0.63] | 2.02<br>[1.67, 2.37] | 0.85<br>[0.67, 1.02] | 1.09<br>[0.91, 1.26] | 0.08<br>[0.07, 0.09] |
| Hardin | 7.51<br>[6.49, 8.57] | 2.19<br>[1.85, 2.62] | 5<br>[4.31, 5.73] | 0.33<br>[0.29, 0.38] | 1.32<br>[1.13, 1.55] | 0.58<br>[0.49, 0.7] | 0.69<br>[0.59, 0.8] | 0.05<br>[0.04, 0.06] |
| Harris | 9.49<br>[8.23, 11.16] | 2.68<br>[2.23, 3.24] | 6.43<br>[5.68, 7.59] | 0.37<br>[0.33, 0.44] | 1.62<br>[1.38, 1.91] | 0.71<br>[0.58, 0.86] | 0.86<br>[0.74, 1.01] | 0.06<br>[0.05, 0.07] |
| Harrison | 12.58<br>[10.89, 14.55] | 2.92<br>[2.45, 3.56] | 9.14<br>[7.98, 10.48] | 0.49<br>[0.42, 0.57] | 1.99<br>[1.72, 2.31] | 0.76<br>[0.64, 0.92] | 1.15<br>[1, 1.33] | 0.07<br>[0.06, 0.09] |
| Hartley | 10.05<br>[8.68, 11.66] | 3.05<br>[2.56, 3.7] | 6.51<br>[5.7, 7.5] | 0.49<br>[0.42, 0.57] | 1.74<br>[1.5, 2.05] | 0.81<br>[0.66, 0.99] | 0.87<br>[0.75, 1] | 0.07<br>[0.06, 0.09] |
| Haskell | 10.65<br>[9.12, 12.29] | 2.73<br>[2.24, 3.26] | 7.27<br>[6.23, 8.31] | 0.63<br>[0.54, 0.73] | 1.76<br>[1.49, 2.08] | 0.72<br>[0.59, 0.87] | 0.95<br>[0.81, 1.11] | 0.1<br>[0.08, 0.11] |
| Hays | 18.23<br>[15.24, 21.42] | 3.95<br>[3.2, 4.8] | 13.59<br>[11.44, 15.83] | 0.7<br>[0.58, 0.82] | 2.75<br>[2.27, 3.24] | 1.03<br>[0.8, 1.25] | 1.63<br>[1.35, 1.91] | 0.1<br>[0.08, 0.12] |
| Hemphill | 11.52<br>[10.12, 13.05] | 2.4<br>[2.04, 2.83] | 8.7<br>[7.66, 9.94] | 0.4<br>[0.35, 0.46] | 1.8<br>[1.53, 2.09] | 0.63<br>[0.52, 0.74] | 1.1<br>[0.96, 1.29] | 0.06<br>[0.05, 0.07] |
| Henderson | 9.82<br>[8.36, 11.34] | 2.73<br>[2.27, 3.25] | 6.62<br>[5.68, 7.58] | 0.48<br>[0.41, 0.57] | 1.67<br>[1.43, 1.95] | 0.72<br>[0.59, 0.86] | 0.88<br>[0.76, 1.03] | 0.07<br>[0.06, 0.09] |
| Hidalgo | 9.54<br>[8.22, 11.19] | 2.55<br>[2.11, 3.12] | 6.72<br>[5.85, 7.77] | 0.29<br>[0.25, 0.34] | 1.61<br>[1.35, 1.94] | 0.68<br>[0.54, 0.83] | 0.89<br>[0.76, 1.04] | 0.04<br>[0.04, 0.05] |
| Hill | 9.4<br>[8.23, 11.05] | 2.53<br>[2.09, 3.02] | 6.45<br>[5.68, 7.48] | 0.43<br>[0.37, 0.5] | 1.58<br>[1.35, 1.87] | 0.66<br>[0.55, 0.8] | 0.86<br>[0.73, 1.01] | 0.07<br>[0.06, 0.08] |
| Hockley | 11.57<br>[10.2, 13.33] | 3<br>[2.58, 3.57] | 8.15<br>[7.15, 9.42] | 0.41<br>[0.36, 0.48] | 1.89<br>[1.62, 2.23] | 0.79<br>[0.67, 0.96] | 1.05<br>[0.9, 1.23] | 0.06<br>[0.05, 0.07] |
| Hood | 8.83<br>[7.41, 10.06] | 2.35<br>[1.89, 2.76] | 5.98<br>[5.1, 6.79] | 0.48<br>[0.41, 0.55] | 1.5<br>[1.24, 1.74] | 0.62<br>[0.49, 0.74] | 0.81<br>[0.68, 0.93] | 0.07<br>[0.06, 0.09] |
| Hopkins | 8.75<br>[7.68, 10.23] | 2.49<br>[2.09, 2.99] | 5.87<br>[5.12, 6.86] | 0.38<br>[0.33, 0.45] | 1.5<br>[1.31, 1.81] | 0.66<br>[0.55, 0.8] | 0.79<br>[0.69, 0.94] | 0.06<br>[0.05, 0.07] |
| Houston | 9.01<br>[7.71, 10.5] | 2.63<br>[2.22, 3.12] | 5.89<br>[5.06, 6.83] | 0.49<br>[0.42, 0.57] | 1.57<br>[1.35, 1.82] | 0.7<br>[0.58, 0.83] | 0.8<br>[0.68, 0.92] | 0.07<br>[0.06, 0.09] |
| Howard | 11.06<br>[9.38, 13.06] | 2.84<br>[2.31, 3.44] | 7.75<br>[6.59, 9.04] | 0.48<br>[0.4, 0.56] | 1.82<br>[1.52, 2.16] | 0.75<br>[0.6, 0.92] | 1.01<br>[0.85, 1.18] | 0.07<br>[0.06, 0.09] |
| Hudspeth | 37.1<br>[31.84, 43.65] | 6.47<br>[5.41, 7.62] | 27.96<br>[24.28, 32.58] | 2.62<br>[2.21, 3.15] | 5.22<br>[4.4, 6.17] | 1.64<br>[1.36, 1.95] | 3.17<br>[2.71, 3.76] | 0.37<br>[0.31, 0.45] |
| Hunt | 11.24<br>[9.61, 13.19] | 2.92<br>[2.42, 3.5] | 7.86<br>[6.87, 9.21] | 0.44<br>[0.38, 0.51] | 1.85<br>[1.55, 2.18] | 0.77<br>[0.61, 0.93] | 1.01<br>[0.86, 1.18] | 0.07<br>[0.05, 0.08] |
| Hutchinson | 9.81<br>[8.56, 11.35] | 2.42<br>[2.04, 2.91] | 6.97<br>[6.11, 7.97] | 0.4<br>[0.35, 0.47] | 1.61<br>[1.38, 1.92] | 0.63<br>[0.52, 0.79] | 0.91<br>[0.8, 1.07] | 0.06<br>[0.05, 0.07] |
| Irion | 7.2<br>[6.24, 8.24] | 2.21<br>[1.83, 2.59] | 4.59<br>[4.05, 5.24] | 0.36<br>[0.31, 0.41] | 1.31<br>[1.12, 1.51] | 0.59<br>[0.49, 0.71] | 0.65<br>[0.57, 0.74] | 0.06<br>[0.05, 0.06] |
| Jack | 8.74<br>[7.39, 10.1] | 2.16<br>[1.81, 2.58] | 6.16<br>[5.25, 7.16] | 0.43<br>[0.36, 0.5] | 1.45<br>[1.25, 1.7] | 0.57<br>[0.46, 0.7] | 0.82<br>[0.72, 0.96] | 0.06<br>[0.05, 0.08] |
| Jackson | 9.62<br>[8.47, 11.28] | 3.01<br>[2.54, 3.62] | 6.24<br>[5.51, 7.26] | 0.38<br>[0.34, 0.45] | 1.68<br>[1.48, 1.97] | 0.79<br>[0.66, 0.98] | 0.83<br>[0.74, 0.95] | 0.06<br>[0.05, 0.07] |
| Jasper | 10.02<br>[8.39, 11.67] | 2.65<br>[2.19, 3.22] | 6.91<br>[5.82, 7.93] | 0.45<br>[0.37, 0.52] | 1.68<br>[1.41, 1.98] | 0.7<br>[0.57, 0.86] | 0.91<br>[0.78, 1.05] | 0.07<br>[0.06, 0.08] |
| Jeff Davis | 8.5<br>[7.47, 9.65] | 2.71<br>[2.31, 3.15] | 3.75<br>[3.39, 4.19] | 2.07<br>[1.81, 2.39] | 1.61<br>[1.41, 1.88] | 0.73<br>[0.62, 0.88] | 0.56<br>[0.5, 0.63] | 0.33<br>[0.28, 0.38] |
| Jefferson | 8.35<br>[7.18, 9.67] | 2.48<br>[2.06, 2.95] | 5.5<br>[4.79, 6.3] | 0.36<br>[0.31, 0.42] | 1.46<br>[1.26, 1.74] | 0.65<br>[0.54, 0.8] | 0.75<br>[0.64, 0.86] | 0.06<br>[0.05, 0.07] |
| Jim Hogg | 11.85<br>[10.53, 13.7] | 2.65<br>[2.32, 3.18] | 8.8<br>[7.85, 10.06] | 0.37<br>[0.33, 0.43] | 1.86<br>[1.61, 2.2] | 0.69<br>[0.59, 0.84] | 1.11<br>[0.97, 1.3] | 0.06<br>[0.05, 0.07] |
| Jim Wells | 8.62<br>[7.47, 10.28] | 2.3<br>[1.94, 2.82] | 5.98<br>[5.25, 7.08] | 0.32<br>[0.28, 0.39] | 1.46<br>[1.25, 1.76] | 0.61<br>[0.5, 0.75] | 0.81<br>[0.7, 0.96] | 0.05<br>[0.04, 0.06] |
| Johnson | 8.96<br>[7.88, 10.4] | 2.41<br>[2.06, 2.86] | 6.16<br>[5.51, 7.13] | 0.36<br>[0.32, 0.42] | 1.53<br>[1.33, 1.8] | 0.65<br>[0.53, 0.78] | 0.83<br>[0.73, 0.96] | 0.05<br>[0.05, 0.07] |
| Jones | 11.53<br>[9.79, 13.71] | 2.6<br>[2.12, 3.19] | 8.2<br>[7.1, 9.66] | 0.71<br>[0.6, 0.85] | 1.84<br>[1.57, 2.2] | 0.68<br>[0.55, 0.83] | 1.06<br>[0.91, 1.24] | 0.11<br>[0.09, 0.13] |
| Karnes | 11.84<br>[10.32, 13.48] | 2.9<br>[2.47, 3.36] | 8.29<br>[7.34, 9.51] | 0.6<br>[0.53, 0.69] | 1.93<br>[1.61, 2.21] | 0.77<br>[0.63, 0.9] | 1.06<br>[0.91, 1.22] | 0.09<br>[0.08, 0.11] |
| Kaufman | 7.85<br>[6.71, 8.78] | 2.32<br>[1.91, 2.68] | 5.22<br>[4.55, 5.89] | 0.28<br>[0.24, 0.32] | 1.38<br>[1.19, 1.58] | 0.61<br>[0.51, 0.72] | 0.72<br>[0.62, 0.82] | 0.04<br>[0.04, 0.05] |
| Kendall | 10.4<br>[8.84, 12.12] | 2.34<br>[1.94, 2.87] | 7.52<br>[6.36, 8.68] | 0.52<br>[0.44, 0.61] | 1.67<br>[1.42, 1.98] | 0.62<br>[0.51, 0.77] | 0.97<br>[0.82, 1.14] | 0.08<br>[0.07, 0.09] |
| Kenedy | 9.04<br>[8.23, 9.76] | 1.63<br>[1.39, 1.87] | 6.75<br>[6.25, 7.22] | 0.67<br>[0.6, 0.74] | 1.46<br>[1.31, 1.62] | 0.44<br>[0.37, 0.5] | 0.92<br>[0.83, 1.01] | 0.1<br>[0.09, 0.12] |
| Kent | 29.75<br>[26.71, 33.61] | 6.76<br>[5.85, 7.94] | 21.31<br>[19.34, 23.66] | 1.64<br>[1.42, 1.9] | 4.55<br>[3.97, 5.23] | 1.76<br>[1.48, 2.13] | 2.55<br>[2.28, 2.96] | 0.24<br>[0.21, 0.29] |
| Kerr | 13.63<br>[11.82, 15.63] | 3.17<br>[2.67, 3.76] | 9.77<br>[8.48, 11.15] | 0.73<br>[0.63, 0.84] | 2.15<br>[1.83, 2.51] | 0.82<br>[0.69, 1.01] | 1.23<br>[1.07, 1.41] | 0.11<br>[0.09, 0.13] |
| Kimble | 9.71<br>[8.58, 11.37] | 2.84<br>[2.43, 3.37] | 6.27<br>[5.58, 7.24] | 0.59<br>[0.52, 0.69] | 1.69<br>[1.47, 1.99] | 0.75<br>[0.62, 0.91] | 0.84<br>[0.74, 0.98] | 0.09<br>[0.08, 0.11] |
| King | 4.84<br>[4.4, 5.37] | 1.27<br>[1.11, 1.47] | 3.01<br>[2.75, 3.29] | 0.58<br>[0.52, 0.66] | 0.91<br>[0.8, 1.04] | 0.34<br>[0.29, 0.41] | 0.48<br>[0.43, 0.53] | 0.09<br>[0.08, 0.11] |
| Kinney | 18.19<br>[15.58, 21.55] | 3.73<br>[3.09, 4.52] | 13.49<br>[11.59, 15.96] | 0.95<br>[0.8, 1.14] | 2.73<br>[2.32, 3.33] | 0.96<br>[0.8, 1.19] | 1.63<br>[1.38, 1.95] | 0.14<br>[0.12, 0.17] |
| Kleberg | 29.86<br>[24.74, 35.79] | 5.7<br>[4.68, 7.01] | 23.2<br>[19.3, 27.71] | 0.88<br>[0.73, 1.06] | 4.19<br>[3.54, 5.1] | 1.42<br>[1.18, 1.8] | 2.64<br>[2.24, 3.15] | 0.13<br>[0.1, 0.16] |
| Knox | 6.42<br>[5.55, 7.41] | 1.81<br>[1.54, 2.14] | 4.35<br>[3.77, 4.98] | 0.27<br>[0.23, 0.31] | 1.15<br>[0.98, 1.34] | 0.48<br>[0.4, 0.57] | 0.62<br>[0.53, 0.72] | 0.04<br>[0.03, 0.05] |
| La Salle | 33.27<br>[28.81, 38.7] | 6.51<br>[5.53, 7.93] | 25.23<br>[22.02, 29.54] | 1.45<br>[1.24, 1.7] | 4.71<br>[4.03, 5.56] | 1.63<br>[1.36, 2] | 2.86<br>[2.44, 3.36] | 0.21<br>[0.18, 0.25] |
| Lamar | 8.6<br>[7.43, 9.98] | 2.61<br>[2.2, 3.14] | 5.62<br>[4.87, 6.53] | 0.37<br>[0.32, 0.43] | 1.52<br>[1.3, 1.79] | 0.69<br>[0.58, 0.82] | 0.77<br>[0.66, 0.89] | 0.06<br>[0.05, 0.07] |
| Lamb | 9.04<br>[7.8, 10.72] | 2.33<br>[1.95, 2.75] | 6.39<br>[5.58, 7.53] | 0.33<br>[0.29, 0.4] | 1.51<br>[1.29, 1.79] | 0.61<br>[0.51, 0.75] | 0.85<br>[0.73, 0.98] | 0.05<br>[0.04, 0.06] |
| Lampasas | 9.45<br>[8.24, 10.88] | 2.31<br>[1.97, 2.84] | 6.66<br>[5.81, 7.54] | 0.48<br>[0.41, 0.55] | 1.57<br>[1.34, 1.83] | 0.61<br>[0.51, 0.76] | 0.89<br>[0.77, 1.01] | 0.07<br>[0.06, 0.08] |
| Lavaca | 10.13<br>[8.74, 11.67] | 2.8<br>[2.36, 3.39] | 6.9<br>[5.93, 7.95] | 0.45<br>[0.38, 0.51] | 1.7<br>[1.46, 2.02] | 0.74<br>[0.61, 0.91] | 0.9<br>[0.78, 1.04] | 0.07<br>[0.06, 0.08] |
| Lee | 13.52<br>[11.43, 15.77] | 3.45<br>[2.85, 4.09] | 9.44<br>[8.09, 10.94] | 0.63<br>[0.53, 0.73] | 2.18<br>[1.82, 2.53] | 0.9<br>[0.73, 1.09] | 1.18<br>[1, 1.38] | 0.09<br>[0.08, 0.11] |
| Leon | 8.11<br>[7.18, 9.28] | 2.36<br>[2.02, 2.84] | 5.35<br>[4.74, 6.2] | 0.39<br>[0.34, 0.45] | 1.43<br>[1.24, 1.68] | 0.63<br>[0.53, 0.76] | 0.73<br>[0.65, 0.86] | 0.06<br>[0.05, 0.07] |
| Liberty | 7.21<br>[6.4, 8.19] | 2.14<br>[1.81, 2.5] | 4.85<br>[4.31, 5.5] | 0.24<br>[0.21, 0.28] | 1.28<br>[1.12, 1.48] | 0.57<br>[0.47, 0.67] | 0.68<br>[0.6, 0.78] | 0.04<br>[0.03, 0.04] |
| Limestone | 8.86<br>[7.69, 10.65] | 2.44<br>[2.06, 2.97] | 6.01<br>[5.23, 7.13] | 0.42<br>[0.37, 0.51] | 1.52<br>[1.29, 1.83] | 0.64<br>[0.54, 0.8] | 0.81<br>[0.71, 0.96] | 0.06<br>[0.05, 0.08] |
| Lipscomb | 11.03<br>[9.5, 13.06] | 2.61<br>[2.22, 3.14] | 7.98<br>[6.94, 9.41] | 0.43<br>[0.37, 0.51] | 1.77<br>[1.51, 2.14] | 0.69<br>[0.57, 0.84] | 1.03<br>[0.89, 1.22] | 0.06<br>[0.05, 0.08] |
| Live Oak | 8.08<br>[6.93, 9.22] | 2.24<br>[1.85, 2.63] | 5.32<br>[4.58, 6.03] | 0.5<br>[0.42, 0.57] | 1.41<br>[1.2, 1.6] | 0.59<br>[0.49, 0.71] | 0.73<br>[0.63, 0.84] | 0.08<br>[0.06, 0.09] |
| Llano | 10.3<br>[8.9, 12.24] | 2.98<br>[2.52, 3.57] | 6.58<br>[5.6, 7.66] | 0.75<br>[0.63, 0.88] | 1.78<br>[1.52, 2.09] | 0.79<br>[0.65, 0.98] | 0.87<br>[0.75, 1.02] | 0.11<br>[0.1, 0.13] |
| Lubbock | 22.51<br>[18.61, 26.76] | 4.71<br>[3.87, 5.64] | 17.08<br>[14.21, 20.28] | 0.76<br>[0.62, 0.9] | 3.3<br>[2.77, 3.94] | 1.2<br>[0.97, 1.45] | 1.99<br>[1.67, 2.37] | 0.11<br>[0.09, 0.13] |
| Lynn | 7.06<br>[6.11, 8] | 1.97<br>[1.64, 2.33] | 4.77<br>[4.22, 5.42] | 0.27<br>[0.24, 0.31] | 1.25<br>[1.07, 1.43] | 0.53<br>[0.44, 0.64] | 0.67<br>[0.59, 0.77] | 0.04<br>[0.04, 0.05] |
| Madison | 12.5<br>[10.69, 14.32] | 3.75<br>[3.1, 4.47] | 8.18<br>[7.03, 9.41] | 0.55<br>[0.47, 0.64] | 2.11<br>[1.8, 2.47] | 0.98<br>[0.8, 1.19] | 1.04<br>[0.9, 1.21] | 0.08<br>[0.07, 0.1] |
| Marion | 9.22<br>[7.76, 10.65] | 2.31<br>[1.82, 2.8] | 6.33<br>[5.39, 7.28] | 0.57<br>[0.48, 0.66] | 1.54<br>[1.26, 1.86] | 0.61<br>[0.47, 0.76] | 0.85<br>[0.71, 1] | 0.09<br>[0.07, 0.11] |
| Martin | 6.72<br>[5.83, 7.98] | 1.72<br>[1.44, 2.05] | 4.78<br>[4.18, 5.63] | 0.22<br>[0.19, 0.26] | 1.17<br>[1, 1.37] | 0.46<br>[0.38, 0.55] | 0.67<br>[0.59, 0.78] | 0.03<br>[0.03, 0.04] |
| Mason | 12.63<br>[10.78, 14.55] | 2.68<br>[2.22, 3.16] | 9.28<br>[8, 10.62] | 0.69<br>[0.59, 0.8] | 1.97<br>[1.66, 2.3] | 0.7<br>[0.56, 0.84] | 1.17<br>[1, 1.34] | 0.1<br>[0.09, 0.12] |
| Matagorda | 8.46<br>[7.28, 9.91] | 2.67<br>[2.2, 3.26] | 5.43<br>[4.72, 6.34] | 0.34<br>[0.29, 0.4] | 1.5<br>[1.29, 1.8] | 0.71<br>[0.59, 0.87] | 0.75<br>[0.64, 0.86] | 0.05<br>[0.04, 0.06] |
| Maverick | 9.6<br>[8.48, 11.33] | 2.87<br>[2.45, 3.44] | 6.48<br>[5.73, 7.53] | 0.29<br>[0.25, 0.34] | 1.66<br>[1.43, 1.97] | 0.76<br>[0.63, 0.93] | 0.87<br>[0.75, 1] | 0.04<br>[0.04, 0.05] |
| McCulloch | 8.14<br>[7.01, 9.57] | 2.08<br>[1.75, 2.53] | 5.61<br>[4.85, 6.69] | 0.44<br>[0.37, 0.52] | 1.38<br>[1.17, 1.64] | 0.55<br>[0.46, 0.68] | 0.76<br>[0.66, 0.91] | 0.07<br>[0.05, 0.08] |
| McLennan | 20.19<br>[17.52, 24.4] | 4.41<br>[3.77, 5.46] | 15.15<br>[13.18, 18.06] | 0.68<br>[0.59, 0.82] | 3.01<br>[2.55, 3.67] | 1.12<br>[0.93, 1.4] | 1.8<br>[1.51, 2.16] | 0.1<br>[0.08, 0.12] |
| McMullen | 21.31<br>[19.31, 24.01] | 3.81<br>[3.3, 4.52] | 16.57<br>[15.11, 18.48] | 0.91<br>[0.8, 1.06] | 3.2<br>[2.79, 3.63] | 1<br>[0.84, 1.19] | 2.06<br>[1.82, 2.32] | 0.14<br>[0.12, 0.16] |
| Medina | 11.66<br>[10.13, 13.43] | 2.86<br>[2.41, 3.4] | 8.29<br>[7.26, 9.43] | 0.5<br>[0.44, 0.58] | 1.88<br>[1.62, 2.19] | 0.74<br>[0.64, 0.9] | 1.06<br>[0.92, 1.23] | 0.08<br>[0.06, 0.09] |
| Menard | 9.08<br>[7.86, 10.44] | 2.19<br>[1.82, 2.65] | 6.33<br>[5.57, 7.18] | 0.55<br>[0.47, 0.63] | 1.52<br>[1.29, 1.77] | 0.58<br>[0.48, 0.69] | 0.85<br>[0.73, 0.97] | 0.08<br>[0.07, 0.1] |
| Midland | 7.09<br>[6.15, 8.28] | 2.44<br>[2.01, 2.93] | 4.41<br>[3.88, 5.04] | 0.26<br>[0.22, 0.3] | 1.33<br>[1.13, 1.56] | 0.65<br>[0.54, 0.79] | 0.63<br>[0.55, 0.73] | 0.04<br>[0.03, 0.05] |
| Milam | 9.94<br>[8.77, 11.31] | 2.62<br>[2.25, 3.09] | 6.8<br>[6.06, 7.81] | 0.45<br>[0.4, 0.52] | 1.67<br>[1.44, 1.92] | 0.69<br>[0.58, 0.83] | 0.91<br>[0.78, 1.03] | 0.07<br>[0.06, 0.08] |
| Mills | 8.59<br>[7.47, 9.95] | 2.04<br>[1.75, 2.44] | 6.1<br>[5.3, 7] | 0.48<br>[0.41, 0.55] | 1.44<br>[1.24, 1.67] | 0.54<br>[0.46, 0.66] | 0.82<br>[0.71, 0.95] | 0.07<br>[0.06, 0.09] |
| Mitchell | 10.16<br>[8.86, 11.8] | 2.32<br>[1.94, 2.75] | 7.32<br>[6.35, 8.48] | 0.53<br>[0.46, 0.62] | 1.65<br>[1.43, 1.93] | 0.61<br>[0.51, 0.73] | 0.95<br>[0.83, 1.1] | 0.08<br>[0.07, 0.1] |
| Montague | 11.31<br>[9.62, 13.22] | 3.35<br>[2.78, 4.18] | 7.18<br>[6.17, 8.45] | 0.75<br>[0.64, 0.89] | 1.93<br>[1.62, 2.35] | 0.88<br>[0.72, 1.12] | 0.94<br>[0.8, 1.12] | 0.11<br>[0.09, 0.14] |
| Montgomery | 8.41<br>[7.23, 9.61] | 2.35<br>[1.94, 2.75] | 5.73<br>[4.99, 6.51] | 0.34<br>[0.29, 0.39] | 1.45<br>[1.23, 1.69] | 0.62<br>[0.5, 0.74] | 0.78<br>[0.68, 0.9] | 0.05<br>[0.04, 0.06] |
| Moore | 8.75<br>[7.68, 10.01] | 2.89<br>[2.46, 3.48] | 5.58<br>[4.89, 6.35] | 0.27<br>[0.23, 0.3] | 1.57<br>[1.35, 1.82] | 0.78<br>[0.63, 0.91] | 0.76<br>[0.66, 0.87] | 0.04<br>[0.03, 0.05] |
| Morris | 8.28<br>[7.19, 9.66] | 2.14<br>[1.77, 2.65] | 5.78<br>[5.04, 6.66] | 0.4<br>[0.34, 0.47] | 1.41<br>[1.2, 1.68] | 0.56<br>[0.46, 0.7] | 0.79<br>[0.67, 0.91] | 0.06<br>[0.05, 0.07] |
| Motley | 18.68<br>[16.46, 21.03] | 4.41<br>[3.72, 5.1] | 13.38<br>[11.8, 14.75] | 0.94<br>[0.81, 1.07] | 2.92<br>[2.51, 3.41] | 1.15<br>[0.95, 1.36] | 1.64<br>[1.42, 1.89] | 0.14<br>[0.12, 0.16] |
| Nacogdoches | 36.74<br>[31.19, 44.3] | 6.63<br>[5.4, 8.17] | 29.12<br>[24.78, 34.67] | 1.05<br>[0.88, 1.26] | 5.08<br>[4.16, 6.17] | 1.65<br>[1.36, 2] | 3.27<br>[2.68, 3.88] | 0.15<br>[0.12, 0.18] |
| Navarro | 10.8<br>[8.95, 12.49] | 2.77<br>[2.27, 3.25] | 7.63<br>[6.37, 8.77] | 0.38<br>[0.32, 0.45] | 1.78<br>[1.47, 2.07] | 0.73<br>[0.58, 0.85] | 0.99<br>[0.83, 1.15] | 0.06<br>[0.05, 0.07] |
| Newton | 9.6<br>[8.21, 10.91] | 2.51<br>[2.11, 3] | 6.51<br>[5.64, 7.39] | 0.54<br>[0.46, 0.61] | 1.62<br>[1.37, 1.88] | 0.67<br>[0.55, 0.8] | 0.87<br>[0.75, 1] | 0.08<br>[0.07, 0.1] |
| Nolan | 11.26<br>[9.72, 13.14] | 2.8<br>[2.36, 3.37] | 8.04<br>[6.92, 9.26] | 0.44<br>[0.37, 0.51] | 1.83<br>[1.56, 2.16] | 0.73<br>[0.62, 0.89] | 1.03<br>[0.87, 1.2] | 0.07<br>[0.06, 0.08] |
| Nueces | 11.35<br>[9.78, 13.35] | 2.98<br>[2.51, 3.61] | 7.94<br>[6.83, 9.16] | 0.47<br>[0.4, 0.55] | 1.87<br>[1.6, 2.18] | 0.78<br>[0.65, 0.94] | 1.02<br>[0.88, 1.16] | 0.07<br>[0.06, 0.09] |
| Ochiltree | 11<br>[9.6, 12.86] | 2.85<br>[2.41, 3.43] | 7.88<br>[6.88, 9.01] | 0.33<br>[0.29, 0.39] | 1.81<br>[1.6, 2.14] | 0.76<br>[0.64, 0.93] | 1.01<br>[0.89, 1.18] | 0.05<br>[0.04, 0.06] |
| Oldham | 8.83<br>[7.51, 9.96] | 2.39<br>[1.98, 2.8] | 5.83<br>[5.03, 6.54] | 0.59<br>[0.5, 0.67] | 1.52<br>[1.27, 1.75] | 0.64<br>[0.51, 0.77] | 0.79<br>[0.67, 0.91] | 0.09<br>[0.07, 0.11] |
| Orange | 7.96<br>[6.83, 9.07] | 2.31<br>[1.95, 2.75] | 5.28<br>[4.56, 5.99] | 0.34<br>[0.29, 0.38] | 1.4<br>[1.18, 1.6] | 0.61<br>[0.51, 0.74] | 0.73<br>[0.63, 0.83] | 0.05<br>[0.04, 0.06] |
| Palo Pinto | 8.71<br>[7.46, 10] | 2.52<br>[2.07, 2.96] | 5.82<br>[4.95, 6.64] | 0.41<br>[0.34, 0.47] | 1.51<br>[1.28, 1.77] | 0.67<br>[0.53, 0.79] | 0.79<br>[0.67, 0.91] | 0.06<br>[0.05, 0.07] |
| Panola | 11.09<br>[9.43, 12.67] | 2.84<br>[2.33, 3.41] | 7.74<br>[6.63, 8.79] | 0.48<br>[0.4, 0.54] | 1.83<br>[1.53, 2.11] | 0.75<br>[0.61, 0.89] | 1<br>[0.85, 1.12] | 0.07<br>[0.06, 0.08] |
| Parker | 8.15<br>[6.81, 9.18] | 2.22<br>[1.8, 2.59] | 5.58<br>[4.68, 6.25] | 0.36<br>[0.3, 0.41] | 1.4<br>[1.16, 1.6] | 0.59<br>[0.47, 0.69] | 0.76<br>[0.64, 0.86] | 0.06<br>[0.04, 0.06] |
| Parmer | 9.49<br>[8.38, 10.78] | 2.65<br>[2.22, 3.1] | 6.53<br>[5.72, 7.42] | 0.33<br>[0.3, 0.38] | 1.61<br>[1.38, 1.89] | 0.7<br>[0.58, 0.84] | 0.87<br>[0.75, 1.02] | 0.05<br>[0.04, 0.06] |
| Pecos | 10.41<br>[9.02, 12.05] | 2.69<br>[2.25, 3.19] | 7.28<br>[6.37, 8.42] | 0.42<br>[0.37, 0.49] | 1.73<br>[1.47, 2.02] | 0.71<br>[0.59, 0.85] | 0.95<br>[0.82, 1.1] | 0.06<br>[0.05, 0.08] |
| Polk | 7.82<br>[6.9, 8.97] | 2.19<br>[1.88, 2.6] | 5.19<br>[4.55, 5.89] | 0.44<br>[0.39, 0.5] | 1.37<br>[1.19, 1.58] | 0.58<br>[0.49, 0.69] | 0.72<br>[0.63, 0.82] | 0.07<br>[0.06, 0.08] |
| Potter | 8.6<br>[7.47, 9.89] | 2.48<br>[2.07, 2.93] | 5.76<br>[4.99, 6.58] | 0.33<br>[0.29, 0.38] | 1.49<br>[1.28, 1.74] | 0.66<br>[0.55, 0.78] | 0.78<br>[0.67, 0.89] | 0.05<br>[0.04, 0.06] |
| Presidio | 9.51<br>[8.17, 10.81] | 2.6<br>[2.18, 3.01] | 6.53<br>[5.64, 7.46] | 0.35<br>[0.3, 0.4] | 1.61<br>[1.37, 1.85] | 0.69<br>[0.56, 0.81] | 0.86<br>[0.76, 1] | 0.05<br>[0.04, 0.06] |
| Rains | 19.97<br>[16.91, 23.44] | 4.19<br>[3.42, 5.04] | 14.84<br>[12.76, 17.4] | 0.95<br>[0.81, 1.11] | 2.97<br>[2.51, 3.51] | 1.07<br>[0.88, 1.31] | 1.76<br>[1.5, 2.05] | 0.14<br>[0.12, 0.17] |
| Randall | 12.64<br>[10.91, 14.81] | 3.1<br>[2.59, 3.77] | 9.06<br>[7.82, 10.57] | 0.5<br>[0.43, 0.59] | 2.02<br>[1.69, 2.43] | 0.8<br>[0.66, 1] | 1.14<br>[0.98, 1.34] | 0.07<br>[0.06, 0.09] |
| Reagan | 8.84<br>[7.83, 10.22] | 2.55<br>[2.2, 3.03] | 5.97<br>[5.35, 6.88] | 0.31<br>[0.28, 0.36] | 1.53<br>[1.34, 1.78] | 0.68<br>[0.58, 0.81] | 0.81<br>[0.71, 0.93] | 0.05<br>[0.04, 0.06] |
| Red River | 7.21<br>[6.28, 8.38] | 2.21<br>[1.82, 2.64] | 4.59<br>[4.04, 5.25] | 0.41<br>[0.36, 0.48] | 1.31<br>[1.12, 1.54] | 0.59<br>[0.48, 0.7] | 0.65<br>[0.57, 0.75] | 0.06<br>[0.05, 0.07] |
| Reeves | 8.38<br>[7.33, 9.67] | 2.5<br>[2.12, 2.96] | 5.54<br>[4.87, 6.35] | 0.31<br>[0.27, 0.35] | 1.46<br>[1.26, 1.73] | 0.66<br>[0.55, 0.8] | 0.75<br>[0.67, 0.87] | 0.05<br>[0.04, 0.05] |
| Refugio | 9.58<br>[8.24, 11.09] | 1.94<br>[1.63, 2.4] | 7.15<br>[6.18, 8.23] | 0.45<br>[0.39, 0.53] | 1.51<br>[1.3, 1.77] | 0.51<br>[0.42, 0.63] | 0.94<br>[0.8, 1.08] | 0.07<br>[0.06, 0.08] |
| Roberts | 7.08<br>[6.25, 8.11] | 2.16<br>[1.82, 2.67] | 4.58<br>[4.09, 5.14] | 0.33<br>[0.29, 0.38] | 1.28<br>[1.09, 1.51] | 0.58<br>[0.49, 0.72] | 0.65<br>[0.58, 0.73] | 0.05<br>[0.04, 0.06] |
| Robertson | 8.73<br>[7.68, 10.02] | 2.59<br>[2.23, 3.05] | 5.7<br>[5.04, 6.56] | 0.39<br>[0.34, 0.45] | 1.53<br>[1.31, 1.8] | 0.69<br>[0.58, 0.81] | 0.77<br>[0.68, 0.89] | 0.06<br>[0.05, 0.07] |
| Rockwall | 8.36<br>[7.35, 9.87] | 2.15<br>[1.79, 2.59] | 5.91<br>[5.17, 6.93] | 0.33<br>[0.29, 0.39] | 1.41<br>[1.21, 1.7] | 0.57<br>[0.46, 0.69] | 0.8<br>[0.7, 0.95] | 0.05<br>[0.04, 0.06] |
| Runnels | 8.79<br>[7.65, 10.12] | 2.48<br>[2.13, 2.93] | 5.92<br>[5.17, 6.77] | 0.4<br>[0.35, 0.46] | 1.52<br>[1.32, 1.76] | 0.66<br>[0.56, 0.79] | 0.79<br>[0.7, 0.93] | 0.06<br>[0.05, 0.07] |
| Rusk | 8.95<br>[7.6, 10.22] | 2.29<br>[1.93, 2.73] | 6.19<br>[5.35, 7.09] | 0.42<br>[0.36, 0.49] | 1.51<br>[1.28, 1.76] | 0.61<br>[0.5, 0.73] | 0.84<br>[0.7, 0.97] | 0.06<br>[0.05, 0.08] |
| Sabine | 10.28<br>[8.9, 11.69] | 3.12<br>[2.61, 3.64] | 6.51<br>[5.7, 7.36] | 0.63<br>[0.55, 0.73] | 1.79<br>[1.52, 2.07] | 0.82<br>[0.68, 0.98] | 0.87<br>[0.76, 0.99] | 0.1<br>[0.08, 0.11] |
| San Augustine | 7.56<br>[6.65, 8.87] | 2.26<br>[1.92, 2.74] | 4.87<br>[4.3, 5.75] | 0.43<br>[0.38, 0.52] | 1.35<br>[1.17, 1.63] | 0.6<br>[0.5, 0.73] | 0.68<br>[0.6, 0.8] | 0.07<br>[0.06, 0.08] |
| San Jacinto | 8.9<br>[7.58, 10.11] | 2.34<br>[1.91, 2.74] | 6.12<br>[5.27, 6.91] | 0.46<br>[0.39, 0.52] | 1.51<br>[1.27, 1.73] | 0.62<br>[0.51, 0.73] | 0.82<br>[0.71, 0.94] | 0.07<br>[0.06, 0.08] |
| San Patricio | 9.45<br>[8.13, 11.18] | 2.61<br>[2.18, 3.26] | 6.46<br>[5.61, 7.6] | 0.37<br>[0.32, 0.44] | 1.6<br>[1.36, 1.91] | 0.69<br>[0.57, 0.85] | 0.86<br>[0.74, 1] | 0.06<br>[0.05, 0.07] |
| San Saba | 9.46<br>[8.18, 10.77] | 2.32<br>[1.96, 2.75] | 6.61<br>[5.75, 7.45] | 0.54<br>[0.47, 0.62] | 1.57<br>[1.33, 1.81] | 0.61<br>[0.5, 0.73] | 0.88<br>[0.75, 0.99] | 0.08<br>[0.07, 0.1] |
| Schleicher | 21.93<br>[18.85, 25.65] | 4.86<br>[3.93, 5.82] | 16.25<br>[14.04, 18.96] | 0.79<br>[0.67, 0.94] | 3.3<br>[2.78, 3.93] | 1.25<br>[1, 1.51] | 1.94<br>[1.64, 2.28] | 0.12<br>[0.09, 0.14] |
| Scurry | 11.28<br>[9.62, 13.03] | 2.82<br>[2.38, 3.35] | 8<br>[6.94, 9.23] | 0.41<br>[0.35, 0.47] | 1.82<br>[1.55, 2.13] | 0.74<br>[0.62, 0.88] | 1.03<br>[0.88, 1.18] | 0.06<br>[0.05, 0.07] |
| Shackelford | 16.17<br>[13.79, 19.1] | 4.23<br>[3.44, 5.13] | 10.93<br>[9.37, 12.85] | 0.96<br>[0.83, 1.15] | 2.61<br>[2.18, 3.08] | 1.09<br>[0.89, 1.35] | 1.35<br>[1.16, 1.56] | 0.14<br>[0.12, 0.17] |
| Shelby | 8.96<br>[7.82, 10.14] | 2.65<br>[2.27, 3.09] | 5.94<br>[5.15, 6.74] | 0.35<br>[0.3, 0.4] | 1.57<br>[1.34, 1.8] | 0.71<br>[0.59, 0.82] | 0.8<br>[0.7, 0.92] | 0.05<br>[0.05, 0.06] |
| Sherman | 11.59<br>[10.22, 13.02] | 3.11<br>[2.6, 3.6] | 8.08<br>[7.13, 9.04] | 0.4<br>[0.35, 0.46] | 1.91<br>[1.66, 2.18] | 0.82<br>[0.68, 0.96] | 1.04<br>[0.9, 1.19] | 0.06<br>[0.05, 0.07] |
| Smith | 11.57<br>[10.11, 13.35] | 2.97<br>[2.51, 3.53] | 8.12<br>[7.13, 9.27] | 0.46<br>[0.4, 0.54] | 1.9<br>[1.63, 2.21] | 0.78<br>[0.64, 0.94] | 1.04<br>[0.91, 1.21] | 0.07<br>[0.06, 0.08] |
| Somervell | 8.87<br>[7.8, 10.3] | 2.28<br>[1.92, 2.71] | 6.15<br>[5.45, 7.03] | 0.45<br>[0.39, 0.52] | 1.49<br>[1.29, 1.75] | 0.6<br>[0.5, 0.73] | 0.82<br>[0.72, 0.95] | 0.07<br>[0.06, 0.08] |
| Starr | 8.64<br>[7.44, 10.16] | 2.51<br>[2.07, 3.05] | 5.86<br>[5.06, 6.79] | 0.25<br>[0.22, 0.29] | 1.49<br>[1.27, 1.8] | 0.67<br>[0.54, 0.83] | 0.79<br>[0.68, 0.92] | 0.04<br>[0.03, 0.05] |
| Stephens | 17.14<br>[14.51, 20.28] | 3.62<br>[3.01, 4.39] | 12.82<br>[10.88, 14.89] | 0.76<br>[0.63, 0.9] | 2.58<br>[2.17, 3.11] | 0.93<br>[0.78, 1.15] | 1.54<br>[1.3, 1.83] | 0.11<br>[0.09, 0.13] |
| Sterling | 15.78<br>[13.42, 18.26] | 5.26<br>[4.34, 6.38] | 9.97<br>[8.74, 11.34] | 0.53<br>[0.45, 0.62] | 2.71<br>[2.26, 3.18] | 1.39<br>[1.12, 1.66] | 1.25<br>[1.08, 1.47] | 0.08<br>[0.07, 0.1] |
| Sutton | 8.37<br>[7.43, 9.66] | 2.11<br>[1.77, 2.48] | 5.88<br>[5.24, 6.78] | 0.37<br>[0.33, 0.44] | 1.42<br>[1.22, 1.64] | 0.56<br>[0.46, 0.67] | 0.8<br>[0.7, 0.93] | 0.06<br>[0.05, 0.07] |
| Swisher | 11.14<br>[9.71, 12.95] | 2.67<br>[2.26, 3.15] | 8.01<br>[6.98, 9.29] | 0.46<br>[0.4, 0.54] | 1.8<br>[1.55, 2.13] | 0.7<br>[0.59, 0.84] | 1.03<br>[0.9, 1.22] | 0.07<br>[0.06, 0.08] |
| Tarrant | 10.48<br>[8.85, 12.29] | 2.74<br>[2.24, 3.34] | 7.31<br>[6.15, 8.48] | 0.41<br>[0.34, 0.48] | 1.73<br>[1.45, 2.04] | 0.72<br>[0.59, 0.89] | 0.95<br>[0.8, 1.11] | 0.06<br>[0.05, 0.07] |
| Taylor | 17.01<br>[14.15, 20.13] | 4.04<br>[3.3, 4.92] | 12.45<br>[10.29, 14.56] | 0.59<br>[0.48, 0.7] | 2.62<br>[2.14, 3.13] | 1.04<br>[0.85, 1.27] | 1.5<br>[1.23, 1.78] | 0.09<br>[0.07, 0.1] |
| Terrell | 7.72<br>[6.89, 8.59] | 1.67<br>[1.41, 1.91] | 5.58<br>[5.01, 6.21] | 0.47<br>[0.41, 0.53] | 1.29<br>[1.13, 1.45] | 0.45<br>[0.38, 0.53] | 0.77<br>[0.68, 0.86] | 0.07<br>[0.06, 0.08] |
| Terry | 9.1<br>[7.79, 10.65] | 2.75<br>[2.29, 3.29] | 6.04<br>[5.28, 6.91] | 0.31<br>[0.27, 0.37] | 1.59<br>[1.33, 1.84] | 0.73<br>[0.59, 0.88] | 0.81<br>[0.69, 0.93] | 0.05<br>[0.04, 0.06] |
| Throckmorton | 15.54<br>[13.48, 18.03] | 4.92<br>[4.24, 5.85] | 9.64<br>[8.34, 11.05] | 1.01<br>[0.87, 1.19] | 2.65<br>[2.3, 3.14] | 1.29<br>[1.1, 1.56] | 1.22<br>[1.05, 1.42] | 0.15<br>[0.13, 0.18] |
| Titus | 9.17<br>[7.87, 10.62] | 2.58<br>[2.09, 3.05] | 6.29<br>[5.41, 7.21] | 0.33<br>[0.28, 0.37] | 1.58<br>[1.31, 1.84] | 0.68<br>[0.54, 0.82] | 0.84<br>[0.73, 0.97] | 0.05<br>[0.04, 0.06] |
| Tom Green | 12.57<br>[10.94, 14.97] | 3.2<br>[2.68, 3.84] | 8.85<br>[7.76, 10.49] | 0.5<br>[0.43, 0.59] | 2.04<br>[1.73, 2.42] | 0.84<br>[0.69, 1.03] | 1.13<br>[0.96, 1.34] | 0.07<br>[0.06, 0.09] |
| Travis | 14.19<br>[12, 17.2] | 3.7<br>[3.07, 4.52] | 9.76<br>[8.25, 11.92] | 0.69<br>[0.58, 0.85] | 2.28<br>[1.91, 2.84] | 0.96<br>[0.76, 1.2] | 1.22<br>[1.03, 1.48] | 0.1<br>[0.09, 0.13] |
| Trinity | 9.71<br>[8.59, 11.29] | 2.51<br>[2.12, 3.01] | 6.66<br>[5.95, 7.67] | 0.53<br>[0.47, 0.62] | 1.64<br>[1.42, 1.93] | 0.66<br>[0.55, 0.8] | 0.89<br>[0.78, 1.03] | 0.08<br>[0.07, 0.1] |
| Tyler | 10.5<br>[8.95, 12.33] | 2.71<br>[2.25, 3.23] | 7.21<br>[6.17, 8.48] | 0.58<br>[0.49, 0.69] | 1.74<br>[1.46, 2.08] | 0.71<br>[0.59, 0.88] | 0.94<br>[0.8, 1.1] | 0.09<br>[0.07, 0.11] |
| Upshur | 8.79<br>[7.75, 10.34] | 2.38<br>[2.04, 2.9] | 6.02<br>[5.35, 7.01] | 0.4<br>[0.35, 0.46] | 1.5<br>[1.3, 1.76] | 0.63<br>[0.53, 0.78] | 0.81<br>[0.7, 0.94] | 0.06<br>[0.05, 0.07] |
| Upton | 10.32<br>[8.69, 11.95] | 2.06<br>[1.71, 2.5] | 7.89<br>[6.65, 9] | 0.38<br>[0.31, 0.44] | 1.61<br>[1.36, 1.89] | 0.54<br>[0.45, 0.66] | 1.01<br>[0.86, 1.18] | 0.06<br>[0.05, 0.07] |
| Uvalde | 13<br>[10.86, 15.32] | 3.68<br>[3, 4.33] | 8.86<br>[7.45, 10.42] | 0.44<br>[0.37, 0.52] | 2.15<br>[1.77, 2.52] | 0.96<br>[0.78, 1.15] | 1.12<br>[0.94, 1.33] | 0.07<br>[0.05, 0.08] |
| Val Verde | 10.24<br>[9.05, 12.23] | 3.02<br>[2.66, 3.79] | 6.87<br>[6.02, 8.06] | 0.35<br>[0.3, 0.42] | 1.76<br>[1.55, 2.11] | 0.8<br>[0.69, 1.01] | 0.91<br>[0.79, 1.05] | 0.05<br>[0.05, 0.06] |
| Van Zandt | 9.18<br>[8.01, 10.45] | 2.41<br>[2.06, 2.87] | 6.3<br>[5.53, 7.17] | 0.43<br>[0.37, 0.49] | 1.55<br>[1.34, 1.82] | 0.64<br>[0.54, 0.77] | 0.84<br>[0.73, 0.97] | 0.07<br>[0.05, 0.08] |
| Victoria | 9.52<br>[8.32, 11.18] | 2.72<br>[2.3, 3.3] | 6.42<br>[5.65, 7.43] | 0.39<br>[0.34, 0.45] | 1.64<br>[1.4, 1.96] | 0.72<br>[0.6, 0.89] | 0.86<br>[0.75, 1.02] | 0.06<br>[0.05, 0.07] |
| Walker | 71.95<br>[57.36, 87.16] | 10.37<br>[7.98, 13.08] | 58.46<br>[46.95, 70.66] | 3.03<br>[2.41, 3.7] | 9.12<br>[7.23, 11.17] | 2.53<br>[1.94, 3.26] | 6.17<br>[4.94, 7.65] | 0.41<br>[0.33, 0.51] |
| Waller | 68.07<br>[55.11, 83.94] | 9.63<br>[7.43, 11.73] | 56.92<br>[46.32, 70.14] | 1.53<br>[1.24, 1.89] | 8.53<br>[6.77, 10.81] | 2.32<br>[1.83, 2.9] | 5.98<br>[4.84, 7.49] | 0.21<br>[0.17, 0.26] |
| Ward | 8.53<br>[7.39, 9.88] | 2.56<br>[2.13, 3.07] | 5.66<br>[4.9, 6.5] | 0.3<br>[0.26, 0.35] | 1.5<br>[1.3, 1.77] | 0.68<br>[0.56, 0.83] | 0.77<br>[0.67, 0.89] | 0.05<br>[0.04, 0.05] |
| Washington | 37.1<br>[30.86, 43.83] | 5.93<br>[4.82, 7.3] | 29.92<br>[24.79, 35.51] | 1.19<br>[0.99, 1.42] | 4.97<br>[4.08, 6.02] | 1.48<br>[1.18, 1.76] | 3.31<br>[2.72, 3.96] | 0.17<br>[0.14, 0.21] |
| Webb | 9.96<br>[8.63, 11.52] | 2.74<br>[2.34, 3.21] | 6.93<br>[6.05, 8] | 0.29<br>[0.26, 0.34] | 1.68<br>[1.43, 1.97] | 0.72<br>[0.6, 0.87] | 0.91<br>[0.79, 1.06] | 0.04<br>[0.04, 0.05] |
| Wharton | 10.58<br>[9.26, 12.25] | 2.99<br>[2.51, 3.49] | 7.18<br>[6.33, 8.3] | 0.41<br>[0.36, 0.48] | 1.78<br>[1.54, 2.1] | 0.78<br>[0.64, 0.93] | 0.94<br>[0.83, 1.09] | 0.06<br>[0.05, 0.07] |
| Wheeler | 8.17<br>[7, 9.41] | 2.05<br>[1.72, 2.4] | 5.74<br>[5.04, 6.58] | 0.35<br>[0.3, 0.41] | 1.38<br>[1.18, 1.61] | 0.55<br>[0.45, 0.64] | 0.78<br>[0.68, 0.9] | 0.05<br>[0.04, 0.06] |
| Wichita | 20.68<br>[17.14, 24.26] | 4.62<br>[3.73, 5.56] | 15.37<br>[12.8, 17.95] | 0.73<br>[0.6, 0.86] | 3.1<br>[2.57, 3.69] | 1.18<br>[0.94, 1.44] | 1.82<br>[1.55, 2.14] | 0.11<br>[0.09, 0.13] |
| Wilbarger | 16.7<br>[14.21, 19.53] | 4.27<br>[3.43, 5.05] | 11.73<br>[10.02, 13.67] | 0.73<br>[0.62, 0.86] | 2.63<br>[2.2, 3.13] | 1.09<br>[0.86, 1.33] | 1.42<br>[1.23, 1.66] | 0.11<br>[0.09, 0.13] |
| Willacy | 14.43<br>[12.55, 16.51] | 3.29<br>[2.7, 3.89] | 10.59<br>[9.25, 12.07] | 0.55<br>[0.48, 0.63] | 2.25<br>[1.91, 2.61] | 0.85<br>[0.69, 1] | 1.31<br>[1.14, 1.49] | 0.08<br>[0.07, 0.09] |
| Williamson | 8.24<br>[7.19, 9.63] | 2.35<br>[2, 2.84] | 5.52<br>[4.83, 6.4] | 0.38<br>[0.33, 0.45] | 1.43<br>[1.24, 1.7] | 0.62<br>[0.53, 0.77] | 0.75<br>[0.66, 0.88] | 0.06<br>[0.05, 0.07] |
| Wilson | 8.21<br>[7.22, 9.5] | 2.17<br>[1.86, 2.59] | 5.65<br>[5.02, 6.42] | 0.38<br>[0.33, 0.44] | 1.4<br>[1.22, 1.64] | 0.58<br>[0.49, 0.7] | 0.77<br>[0.68, 0.89] | 0.06<br>[0.05, 0.07] |
| Winkler | 9.72<br>[8.31, 11.49] | 2.82<br>[2.35, 3.46] | 6.58<br>[5.62, 7.75] | 0.33<br>[0.28, 0.39] | 1.67<br>[1.43, 2] | 0.74<br>[0.62, 0.91] | 0.87<br>[0.75, 1.03] | 0.05<br>[0.04, 0.06] |
| Wise | 9.26<br>[7.98, 10.8] | 2.52<br>[2.12, 3.04] | 6.34<br>[5.51, 7.35] | 0.39<br>[0.34, 0.46] | 1.57<br>[1.34, 1.85] | 0.67<br>[0.54, 0.81] | 0.85<br>[0.73, 1] | 0.06<br>[0.05, 0.07] |
| Wood | 12.14<br>[10.5, 14.33] | 2.76<br>[2.24, 3.37] | 8.72<br>[7.58, 10.17] | 0.66<br>[0.57, 0.78] | 1.92<br>[1.64, 2.32] | 0.72<br>[0.59, 0.89] | 1.1<br>[0.94, 1.31] | 0.1<br>[0.08, 0.12] |
| Yoakum | 10.29<br>[8.71, 11.99] | 2.94<br>[2.39, 3.5] | 7.1<br>[6.05, 8.18] | 0.28<br>[0.23, 0.33] | 1.75<br>[1.45, 2.07] | 0.78<br>[0.61, 0.94] | 0.93<br>[0.8, 1.09] | 0.04<br>[0.04, 0.05] |
| Young | 8.88<br>[7.67, 10.31] | 2.43<br>[1.98, 2.92] | 6.05<br>[5.28, 7.01] | 0.4<br>[0.34, 0.46] | 1.52<br>[1.29, 1.76] | 0.64<br>[0.53, 0.77] | 0.82<br>[0.7, 0.95] | 0.06<br>[0.05, 0.07] |
| Zapata | 8.73<br>[7.51, 10.36] | 2.2<br>[1.83, 2.71] | 6.27<br>[5.43, 7.37] | 0.26<br>[0.22, 0.31] | 1.46<br>[1.21, 1.73] | 0.58<br>[0.48, 0.72] | 0.84<br>[0.71, 0.99] | 0.04<br>[0.03, 0.05] |
| Zavala | 10.09<br>[8.64, 11.78] | 2.78<br>[2.34, 3.41] | 6.99<br>[5.95, 8.16] | 0.32<br>[0.27, 0.38] | 1.7<br>[1.43, 2.03] | 0.73<br>[0.61, 0.91] | 0.92<br>[0.78, 1.08] | 0.05<br>[0.04, 0.06] |

## Notes

### Competing Interest Statement

The authors have declared no competing interest.

### Summary of Updates

Add the full author list

